# An intermittent energy restriction diet ameliorates comorbid MASLD and T2DM through the *Klebsiella pneumoniae*/LPS/Hepatic HADHA-K353 acetylation axis

**DOI:** 10.64898/2026.07.10.26357698

**Authors:** Wu Luo, Ruiyu Wu, Zhihao Peng, Kunyao Tan, Dan Zhu, Xuanping Ouyang, Zhiyong Xiao, Zihao Liu, Hui Liu, Xiaoya Chang, Zhuoming Yin, Jian Li, Xinyu Zhang, Xuanming Liu, Dongbo Liu

## Abstract

The intermittent energy restriction (iER) represents an effective dietary strategy for improving metabolic diseases including metabolic dysfunction-associated steatotic liver disease (MASLD) and type 2 diabetes mellitus (T2DM), yet the underlying mechanisms remain elusive. In this study, we integrated human clinical data, mouse models, and in vitro experiments to investigate the role of iER in modulating the gut-liver axis in comorbid MASLD and T2DM. We demonstrate that an iER diet improves hyperglycemia, hepatic steatosis and decreases the abundance of gut pathogen Klebsiella pneumoniae, which is strongly associated with reductions in blood endotoxin, lipopolysaccharide (LPS) levels, suggesting a potential role of K. pneumoniae-derived LPS in mediating effects of the iER on hepatometabolic improvements. We confirm that K. pneumoniae-derived LPS exacerbates lipid accumulation and inflammation using an in vitro model. Mechanistically, we reveal a core target of protein lysine acetylation (Kac), hydroxyacyl-CoA dehydrogenase α-subunit (HADHA) Lys353 in the liver of db/db mice through a multi-omics analysis. The iER decreases HADHA-K353 acetylation and enhances its enzyme activity. A Kac-mimicking mutation (K353R) increases its enzyme activity and stability, blocks its binding to the inflammasome adaptor ASC, and alleviates lipid accumulation and inflammation in K. pneumoniae-derived LPS induced in vitro model. This study provides novel insights into the potential benefits of the iER in comorbid MASLD and T2DM.

## Introduction

Metabolic dysfunction-associated steatotic liver disease (MASLD), formerly named non-alcoholic fatty liver disease (NAFLD), and type 2 diabetes mellitus (T2DM) have become a global public health concern with an increasing prevalence worldwide. It is estimated that the global prevalence of MASLD among individuals with T2DM is 55.48%, and this prevalence increases to 64.36% among T2DM patients who are also obese[1]. The coexistence of MASLD and T2DM increases the risk of multiple intrahepatic and extrahepatic adverse events, leading to worse metabolic status and elevated risk of all-cause mortality[2]. Therefore, the co-management of this comorbidity is urgently needed. MASLD is characterized by excessive hepatic fat accumulation, which subsequently leads to inflammation and liver injury, known as metabolic dysfunction-associated steatohepatitis (MASH), ultimately resulting in liver fibrosis, cirrhosis, and even hepatocellular carcinoma[3]. The bidirectional relationship between MASLD and T2DM exacerbates disease progression, indicating that targeting MASLD therapies can improve the glycemic control of this comorbidity[4]. Nevertheless, to date, only one MASH pharmaceutical product, Resmetirom, a thyroid hormone receptor β (TRβ) agonist, has received FDA approval for commercial distribution. The therapeutic response rate among patients is limited to 25-30%, and the high cost of the drug indicates that there is a continued necessity to identify novel interventional targets and strategies for MASLD[5].

The gut microbiota and/or microbiota-derived metabolites play a crucial role in the development of MASLD and T2DM[6, 7, 8], yet the exact mechanisms remain to be elucidated. MASLD and T2DM share common pathophysiological characteristics, including gut microbiota dysbiosis, aberrant hepatic lipid metabolism, insulin resistance and metabolic inflammation[9, 10]. Diet has been tightly linked to these metabolic processes through gut microbiota and its derived metabolites. Poor dietary habits, characterized by high intake of processed meats, low-quality carbohydrates, and sugary beverages, coupled with low consumption of plant-based diet, plays a significant physiological role in the growth, proliferation, and invasive pathogenic mechanisms of gut opportunistic pathogens such as *Enterobacteriaceae*, including *Escherichia coli* (*E. coli*) and *Klebsiella pneumoniae* (*K. pneumoniae*)[11, 12, 13]. These pathogens specifically sense nutritional changes in the intestinal microenvironment and undergo adaptive modifications, producing various microbial metabolites in response. These microbial metabolites may act as signaling molecules, penetrating the intestinal barrier and entering the bloodstream to interact with host cells, leading to disturbance of host metabolism[11, 14]. Furthermore, some specific pathogenic bacteria can translocate into the bloodstream through the intestinal barrier, and these extra-intestinal bacteria may further disrupt host metabolism via local immune regulation[15]. The interaction between nutrients and pathogens in the development of comorbid MASLD and T2DM provides valuable insights for dietary intervention targeting pathogens in the co-management of MASLD and T2DM.

Intermittent energy restriction (iER) has emerged as a promising dietary intervention for MASLD or T2DM in recent years[16, 17, 18, 19]. However, there is a paucity studies investigating populations with concurrent MASLD and T2DM, particularly regarding the molecular mechanisms underlying the benefits of iER in this comorbid condition. Although the role of gut microbiota in mediating the effects of dietary interventions on metabolic disorders has been well established, the mechanisms involving gut microbiota-mediated post-translational modifications (PTMs) of host proteins remain poorly understood and are now gaining increasing attention[20, 21, 22]. Diet not only modulates the gut microbiome but also influences host metabolic responses through PTMs[23]. Pathogens and commensals can alter host PTMs profiles through protein secretion, microbial derived metabolites and diet-regulated metabolic shifts, all of which contribute to the disease development[24, 25, 26]. Lysine acetylation modification, a ubiquitous PTM found on thousands of non-histone proteins, is associated with metabolic reprogramming, cell signaling, and other major cellular functions. By altering the charge and size of modified lysine residues, acetylation may affect protein stability, subcellular localization, and interactions with other signaling molecules[27]. Nevertheless, the biological functions of most lysine acetylation sites in metabolic enzymes remain largely unexplored, particularly their mysterious non-metabolic effects.

In our previous study, we reported an iER diet known as the Chinese Medical Nutrition Therapy (CMNT) regimen, which employs intermittent energy restriction through the replacement of conventional food with a specially designed CMNT diet during energy-restriction periods[28]. The CMNT diet is primarily composed of whole grain rice flour and is supplemented with a blend of medicinal-edible plants such as *Ganoderma lucidum*, *Folium Mori*, *Dioscorea opposita Thunb*., *Radix puerariae*, etc[29]. In this study, we evaluated the effects of the iER diet on hepatometabolic function and glycemic control in patients with MASLD and T2DM, as well as in db/db mice, followed by mechanistic investigations targeting gut microbiota, microbiota-derived metabolites, and hepatic protein acetylation. Using a multi-omics analysis, we identified the enterobacteria *K. pneumoniae*-LPS-hepatic HADHA-K353 acetylation axis as a key mediator linking iER to comorbid MASLD and T2DM. We isolated *K. pneumoniae* strain and extracted it derived LPS from patient with MASLD and T2DM. The *in vitro* model of comorbid MASLD and T2DM was constructed with MIHA cells using *K. pneumoniae-*derived LPS combined with high glucose and high free fatty acids (HGHF) conditions. Subsequently, we explored the functional role of HADHA-K353 acetylation in enzyme activity, protein stability and inflammation signal transduction using an acetylation-null mutant at the cellular level. Our findings provide novel theoretical insights into the effectiveness of iER for the comorbid MASLD and T2DM.

## Results

### The iER improves glycemic control, ameliorates intrahepatic fat and reduces systemic inflammation in patients with MASLD and T2DM

To evaluate the effects of iER on glycemic control, intrahepatic fat reduction, insulin resistance and systemic inflammation in humans, we performed an exploratory pilot clinical study at a single center, which comprised a part of an ongoing multicenter randomized clinical trial (identifier at ClinicalTrials.gov is NCT05439226). The iER group was instructed to consume the provided specific Chinese Medical Nutrition Therapy (CMNT) diet consisting of 6 cycles of 5 consecutive days followed by 10 days of regular food intake and the control group received standard dietary advice (**Figure 1A**). Diet ingredients and calorie information of CMNT diet are provided in **Table S7-S8**. A total of 20 participants (83.3%) in the iER group and 19 participants (79.17%) in the control group completed the trial (**Figure S1**). Baseline characteristics for age, gender, and anthropometric and metabolic variables are presented in **Table S2**. In addition, there was no significant difference in dietary intake of energy and macronutrients between the two groups, as assessed by Food Frequency Questionnaire (FFQ) during the regular food/*ad libitum* diet phase. (**Table S5**). At the end of intervention, participants in the iER group were significantly reduced the use of glucose lowering medications compared with the control group (**Table S6**). No serious adverse events were reported during the course of the study.

**Figure 1.**
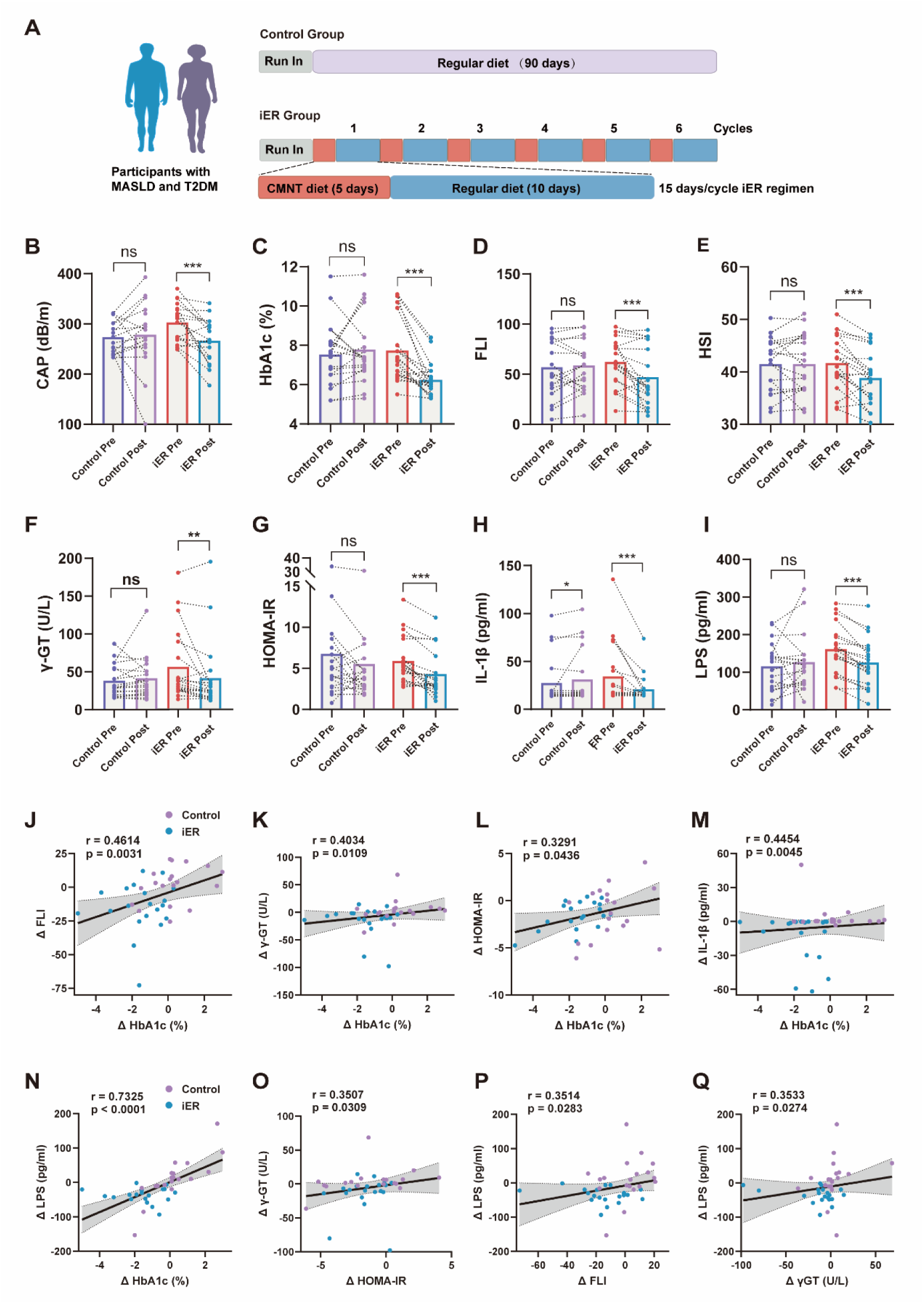
The iER improves glycemic control, ameliorates intrahepatic fat and reduces systemic inflammation in patients with MASLD and T2DM. (A) Schematic overview of clinical experimental design. Participants were randomized to either iER group (n=20) or control group (n=19). Participants in the iER group consumed a low-calorie CMNT diet for 5 consecutive days, followed by a return to their regular diet for the next 10 days. The iER regime lasted for 90 days (6 cycles). Participants in control group continued their regular diet throughout the intervention period. (B-G) Changes in CAP score (B), HbA1c (C), FLI (D), HSI (E), γ-GT(F), HOMA-IR (G), IL-1β (H) and LPS (I) levels at baseline (Pre) and at the end of the study (Post) in both the iER group and the control group. The number of samples for each clinical characteristic is shown in Table S3. Bars show mean, thin lines show paired data. p-values were calculated using two-tailed Wilcoxon signed-rank tests for non-normally distributed paired data and paired t tests for normally distributed data. *p < 0.05, **p < 0.01, ***p < 0.001, ns, no significance. (J-N) Scatter plots of the correlations between changes of HbA1c and changes of FLI (H), serum γ-GT (I), HOMA-IR (L), serum IL1β (M) and serum LPS (N), respectively. (O) Correlations between the changes of HOMA-IR and changes of γ-GT. (P-Q) Correlations between the changes of serum LPS and changes of FLI (P) and serum γ-GT (Q). The correlation analysis were performed in all completed participants (n = 39). Spearman’s correlations were used for non-normally distributed data and Pearson’s correlations were used for normally distributed data. r, correlation coefficient, and p < 0.05 was considered statistically significant. See also Figure S1-S2 and Table S2-S5. Source Data are provided as a Source Data file.

To investigate the effects of iER on intrahepatic fat and glycemic control in patients with MASLD and T2DM, we determined the changes in controlled attenuation parameter (CAP) values by transient elastography (TE) and HbA1c levels during the intervention, which was regard as coprimary outcomes[28]. As shown in **Figures 1B-1C and Table S3**, after 3-month intervention, both CAP values and HbA1c levels in iER group were significantly reduced (p < 0.001 and p < 0.001, respectively), but no significant changes were observed in the control group (p = 0.8622 and p = 0.3978, respectively). The net change of CAP and HbA1c values in the iER group relative to the control group was –38.35 (95% CI: –72.03 to –4.68) and –1.75% (95% CI: –2.62% to –0.88%), respectively. Fatty liver index (FLI) and hepatic steatosis index (HSI), as noninvasive markers of hepatic steatosis, were also applied to quantify the degree of steatosis. After the iER intervention, patients with MASLD and T2DM had significantly lower FLI and HIS indices, as well as fasting plasma glucose (FPG) levels, whereas these values were not significantly altered in the control group (**Figures 1D-E and Figure S2B**). Plasma gamma-glutamyltransferase (γ-GT), alanine aminotransferase (ALT) and aspartate aminotransferase (AST), reflecting the degree of liver damage, were obviously reduced after 3 months of iER regimen, while the control group showed no significant changes (**Figure 1F and Figures S2C-2D**). The DiabetesLiver score and fibrotic non-alcoholic steatohepatitis index (FNI), both established noninvasive liver fibrosis markers, were employed to evaluate fibrosis progression. Following iER intervention, patients with MASLD and T2DM exhibited markedly decreased DiabetesLiver score and FNI value[30], in contrast to the control group, which showed no significant differences (**Figures S2G-S2H**). Together with the improvements in liver function, we observed significant reduction of body mass index (BMI) and significant increase in high density lipoprotein (HDL) in the iER group compared with the control group (**Figures S2A and S2E**). Considering that hyperglycemia, hepatic lipid accumulation, insulin resistance and circulating inflammation show clear biological links to the progression of MASLD and T2DM[31, 32], representative indicators of these biological functions were detected. The homeostasis model assessment-insulin resistance (HOMA-IR), an indicator of insulin resistance, along with serum IL-1β, IL-18 and LPS, indicators of active inflammation, were significantly reduced. In contrast, the control group showed a significant increase in serum IL-1β, while no significant changes were observed in HOMA-IR, IL-18, or LPS (all p > 0.05). – (**Figures 1G-I and Figure S2F**). The unadjusted changes within each group and the adjusted between-group differences for all anthropometric and metabolic variables are detailed in **Table S3** and **Table S4**, respectively. Taken together, these results suggest that iER exerts beneficial metabolic effects on comorbid MASLD and T2DM.

Analyses performed in all completed participants (n = 39) showed that changes in HbA1c from baseline to 3 months were significantly positive associated with changes in FLI (r=0.4616, p=0.0031), γ-GT (r=0.4034, p=0.0109), HOMA-IR (r=0.3291, p=0.0436), IL-1β (r=0.4454, p=0.0045), LPS (r=0.7325, p< 0.0001), TG (r=0.3623, p=0.0234) and DiabetesLiver score (r=0.4230, p=0.0091), and negative associated with changes in HDL (r=-0.4410, p=0.0049) (**Figures 1J-N and Figures S2I-K**). The changes in HOMA-IR was significantly positive correlation with changes in γ-GT (r=0.3507, p=0.0305) (**Figures 1O**). The changes in FLI were significantly positive associated with changes in LPS (r=-0.3514, p=0.0283), DiabetesLiver score (r=0.5469, p=0.0005) and TG (r=0.3812, p=0.0166) and negative associated with changes in HDL (r=-0.3601, p=0.0243) (**Figure 1P and Figures S2L-N**). The changes in LPS were significantly positive correlation with γ-GT (r=0.3533, p=0.0274) (**Figure 1Q**). The correlation analysis of all clinical indices among these participants were shown by the heatmap (**Figure S2O**). These results suggest iER improves comorbid MASLD and T2DM, which is associated with reductions in blood endotoxin, LPS levels. The iER-induced decrease in hepatic steatosis, insulin resistance, and systemic inflammation potentially contributes to the improvement of glucose homeostasis.

### The iER decreases the abundance of the enterobacteria *K. pneumoniae*, which correlates with a reduced blood LPS levels in patients with MASLD and T2DM

To further elucidate how iER might modulate the gut microbiota to influence host liver metabolism, we performed shotgun metagenomic sequencing of fecal samples collected from the participants before and after the iER intervention, aiming to identify specific microbial taxa potentially involved in this regulatory process. Although the Shannon and Chao1 diversity index (α-diversity) and principal component analysis (PCA) analysis revealed no significant changes in structure of gut microbiota following iER intervention (**Figures S3A-S3B**), linear discriminant analysis (LDA) combined with effect size (LEfSe) showed a reduction in the abundance of the phylum Pseudomonadota (Proteobacteria), the class Gammaproteobacteria, the order Enterobacterales, the family Enterobacteriaceae, the genus *Klebsiella*, and the species *K. pneumoniae* after iER intervention compared with baseline (**Figures 2A-2C**). These taxa were among the top 20 dominant bacteria in terms of relative abundance. In contrast, no significant changes were observed in the other top 20 dominant bacteria at various taxonomic levels before and after the intervention (**Figures S3C–S3F**). Among the genus *Klebsiella*, the abundances of multiple species other than *K. pneumoniae*, which had the highest relative abundance, were markedly decreased after iER intervention (**Figures 2D and S3G**). These findings indicate that the beneficial effects of iER on MASLD with comorbid T2DM might be attributed to a decrease in the abundance of *K. pneumoniae*, an opportunistic gut pathogen.

**Figure 2.**
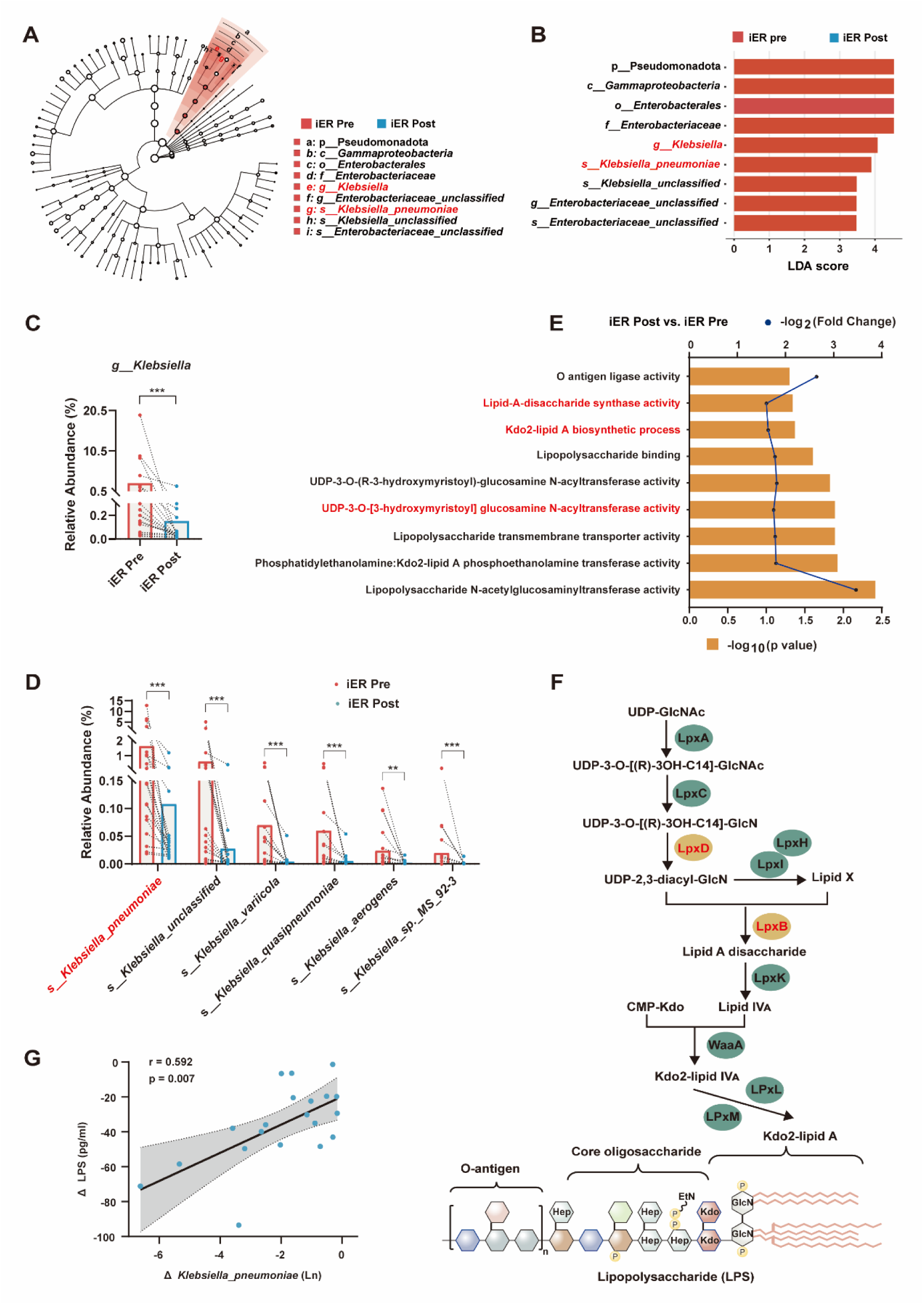
The iER reduces the abundance of Enterobacteria *K. pneumoniae* and circulating LPS level in patients with MASLD and T2DM. (A-B) The linear discriminant analysis (LDA) effect size (LEfSe) analysis of shotgun metagenomic sequencing was conducted to identify bacterial taxa with significant differences in abundance in fecal samples before and after iER intervention (LDA score threshold >3.0, p < 0.05). (A) Taxonomic cladograms generated using LEfSe analysis. (B) LDA value distribution histogram based on LEfSe analysis. **(C)** Change in relative abundance of the genus *Klebsiella* at baseline and the end of iER intervention. **(D)** Changes in the relative abundance of *Klebsiella* at the species level (top 15) before and after the iER intervention. **(E)** LPS-related microbiota functions involved in LPS biosynthesis, binding, transport and modification by GO enrichment analysis based on metagenomic data. **(F)** LPS biosynthesis pathway. Red indicates that coding gene involved in the LPS biosynthesis is changed by iER intervention based on metagenomic sequencing. **(G)** Spearman’s correlations between *K. pneumoniae* abundance and serum LPS. r, Spearman’s correlation coefficient, and p < 0.05 was considered statistically significant. Data are shown as mean ± SEM. p-values were calculated using two-tailed Wilcoxon signed-rank tests for paired comparisons. *p < 0.05, **p < 0.01, ***p < 0.001, ns, no significance. See also **Figure S3**. Source Data are provided as a Source Data file.

Subsequently, the functional potential of the gut microbiota was assessed and functional changes were investigated before and after the iER intervention. Notably, Gene Ontology (GO) enrichment analysis of metagenomic data revealed significant changes in microbiota functions associated with lipopolysaccharide (LPS) biosynthesis, binding, transport, and modification following iER intervention **(Figure 2E)**. Specifically, the enzymatic activities of LpxD (UDP-3-O-[3-hydroxymyristoyl] glucosamine N-acyltransferase) and LpxB (lipid-A disaccharide synthase), which are involved in the Kdo2-lipid A biosynthetic pathway (p < 0.05, Wilcoxon signed-rank tests), were significantly modulated post-iER intervention compared with baseline levels **(Figure 2F)**. Virulence-associated genes involved in LPS modification, including the gene encoding the LPS-modifying enzyme (VF0684), were identified by screening genome sequences against the VFDB 2023 (**Figures S3H**). LPS is a structural component of the outer membrane of gram-negative bacteria and induces a pro-inflammatory innate immune response, which accelerates the development of MASLD[33]. Moreover, LPS from different bacteria exhibit different bioactivities in a species-specific manner, which can be attributed to LPS structural diversity[34]. We found that diminished richness of *K. pneumoniae* exhibited a significantly positive correlation with the lowered serum LPS levels following iER intervention in patients with comorbid MASLD and T2DM (r=0.592, p=0.007) **(Figure 2G)**. Therefore, it is reasonable to consider that changes in *K. pneumoniae* and its derived LPS after the iER intervention can be linked to the improvement of comorbid MASLD and T2DM.

### *K. pneumoniae*-derived LPS exacerbates lipid droplet accumulation, insulin resistance and inflammation in HGHF-induced hepatocytes

Based on the above findings, we further explored through *in vitro* experiments whether *K. pneumoniae*-derived LPS exacerbates hepatic lipid accumulation, insulin resistance, and inflammation in high-glucose-cultured steatotic human normal liver (MIHA) cells, which is used as an *in vitro* model for comorbid MASLD and T2DM. We first isolated a *K. pneumoniae* strain (ZH2.1) from patient with comorbid MASLD and T2DM, which was confirmed by 16S rDNA sequencing and whole-genome sequencing **(Figure 3A)**. The morphology of ZH2.1 was characterized by transmission electron microscopy (TEM) and exhibited Gram-negative character following Gram staining **(Figures 3B-3D)**. The genome of ZH2.1 was sequenced, revealing a genome size of 5,440,791 base pairs and 5,060 predicted open reading frames. Notably, 99.96% of the coding genes were annotated in the NR database **(Figure S4A)**. Using whole-genome analysis, constructing a phylogenetic tree based on single-copy genes from the 49 *Klebsiella* genomes in the GenBank database reveals that ZH2.1 is a member of *K. pneumoniae*, with the closest relative to *K. pneumoniae* KCTC2242 **(Figure 3E)**. A comparative analysis was conducted among six bacterial strains (*K. pneumoniae* ZH2.1, ATCC BAA-2146, HS11286, TH1 and W14, and *E. coli* O157:H7) focusing on genes associated with LPS synthesis and modification. Compared with ATCC-2146, ZH2.1 possesses the rfbB and RfbK1 genes but lacks the wbdB gene **(Figure S4B)**. LPS was extracted from *K. pneumoniae* using an LPS isolation kit. A laboratory *E. coli* isolates served as a control. Silver staining demonstrated distinct compositional differences in LPS derived from different bacterial species, with *E. coli* derived LPS (L2880, sigma) as the control **(Figure S4C)**.

**Figure 3.**
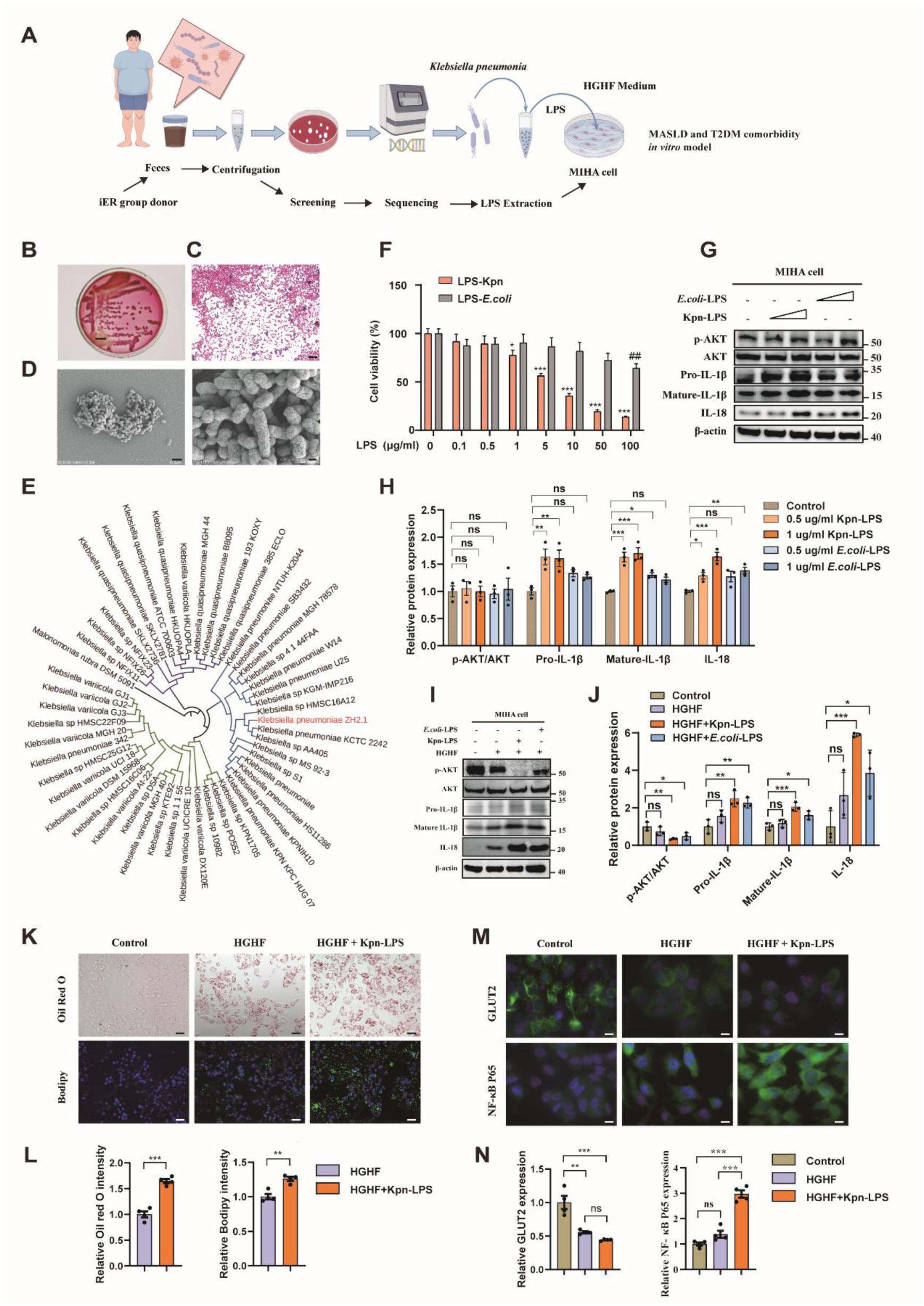
*K. pneumoniae*-derived LPS exacerbates lipid droplet accumulation, insulin resistance and inflammation in HGHF induced MIHA cells. (A) A diagram illustrating *K. pneumoniae* was isolated and cultured from iER group donor and LPS was extracted from *K. pneumoniae* and, in combination with HGHF, was used to establish an *in vitro* model for comorbid T2DM and MASLD. (B) *K. pneumoniae* on a MacConkey Inositol Adonitol Carbenicillin agar plate. Scale bar, 1 cm. **(C)** Gram-stained *K. pneumoniae* display Gram-negative characteristics. Scale bar, 10 μm. **(D)** TEM image of *K. pneumoniae*. Scale bar, 2 μm (Left) and 0.4 μm (Right). **(E)** Evolutionary relationships between the isolated clinical strain ZH2.1 and other *K. pneumoniae* strains based on the genomic BLAST in NCBI. **(F)** The CCK8 assay was used to determine the cytotoxicity of LPS from strains *K. pneumoniae* and *E. coli* on MIHA cells. *p < 0.05 and ***p < 0.001 versus the 0 μg/mL LPS-Kpn group and ##p < 0.01 versus the 0 μg/mL LPS-E.coli group. **(G-H)** Western blotting analysis of the expression of p-AKT, AKT, pro-IL-1β and IL-18 in *K. pneumoniae* derived LPS or *E. coli* derived LPS treated MIHA cells. The relative intensities of proteins were normalized against β-actin. **(I-J)** Western blotting analysis of the expression of p-AKT, AKT, pro-IL-1β and IL-18 in HGHF combined with *K. pneumoniae* or *E. coli* derived LPS. The relative intensities of proteins were normalized against β-actin. **(K-L)** Representative Oil Red O staining, Bodipy 493/503 (green) staining of lipid droplets accumulation. Nucleus marker DAPI (blue). Scale bar, 50 μm. **(M-N)** GLUT2 translocated from cytoplasm to cell membrane and cytoplasmic NF-κB p65 translocation into the nucleus under immunofluorescence after HGHF combined with LPS treatment in MIHA cells. Nucleus marker DAPI (blue), GLUT2 (green) and NF-κB p65 (green) were visualized. Scale bar, 10 μm. Data represent the mean ± SEM. *p < 0.05, **p < 0.01, ***p < 0.001. Significant differences between mean values were determined by Student’s unpaired t-test (L) and one-way ANOVA with Tukey’s multiple comparison test (F, H, J and N). See also **Figure S4**. Source Data are provided as a Source Data file.

LPS from different bacterial strains exhibited differential cytotoxicity in MIHA cells as determined by CCK8 assay **(Figure 3F)**. Insulin signaling and inflammatory factors expression in the hepatocytes were assessed to determine whether *K. pneumoniae*-derived LPS could exacerbates insulin resistance and inflammation. Akt activation, primarily through Akt-Ser473 phosphorylation, plays a critical role in insulin sensitivity[35]. When treated with either *K. pneumoniae*-derived LPS or *E. coli*-derived LPS, MIHA cells showed increased IL-1β and IL-18 protein expression to varying degrees, but did not alter the proteins expression of insulin-stimulated AKT phosphorylation at Ser473 **(Figures 3G-3H)**. However, LPS derived from both *K. pneumoniae* and *E. coli* significantly reduced the protein expression of p-AKT at Ser473 while increasing the protein expression levels of IL-1β and IL-18 in HGHF-induced MIHA cells **(Figures 3I-3J)**. In addition, Oil Red O, Bodipy 493/503 and immunofluorescence staining showed that *K. pneumoniae*-derived LPS increased lipid droplet accumulation, reduced glucose transporter 2 (GLUT2) plasma membrane translocation and induced cytoplasmic NF-κB p65 translocation into the nucleus in HGHF-induced MIHA cells **(Figures 3K-N)**. These results suggest that *K. pneumoniae*-derived LPS exacerbates lipid droplet accumulation, insulin resistance and inflammation in HGHF-indued MIHA cells.

### The iER improves glycemic control and exhibits hepatometabolic protective effects in db/db mice

The db/db BKS mice are a widely used model for studying comorbid MASLD and T2DM[36]. To verify the effects of iER on glucose homeostasis and hepatic metabolic protective effects in db/db mice, 12-week-old male db/db mice were fed 1-day low-calorie CMNT diet (50% of normal daily intake), 3-day very-low-calorie CMNT diet (10% of normal daily intake) and 7-day *ad libitum* diet per cycle for 7 cycles. The db/db mice with *ad libitum* access to regular chow were used as the model group, while age-matched wild-type (WT) mice served as healthy controls **(Figure 4A)**. Specifically, iER substantially reduced the bodyweight of db/db mice at the end of the intervention, and average food intake and daily water intake also were decreased, and less urine was observed during the intervention **(Figures S5A-S5C)**. The elevated fasting blood glucose levels in db/db mice were significantly reduced, while blood insulin levels were increased by the iER intervention **(Figures 4B-4C)**. HOMA-IR values, which reflected insulin resistance, were significantly reduced after iER intervention in db/db mice **(Figure 4D)**. Furthermore, a marked increase in insulin sensitivity was observed by iER intervention, as evidenced by a significantly acute hypoglycemic response to insulin and smaller areas under the insulin tolerance test (ITT) curves compared with the *ad libitum* fed db/db mice **(Figures 4E-4F)**. Transmission electron microscopy (TEM) analysis of the liver revealed a considerable amount of large lipid droplets (LDs) and severely shriveled mitochondria in db/db mice. After the iER intervention, the number and size of LDs markedly decreased, mitochondrial injury was significantly improved and mitochondrial number in was dramatically increased, indicating enhanced mitochondrial function **(Figure 4G)**. HE and Oil Red O staining demonstrated that iER reduced hepatic steatosis, and Masson staining showed diminished hepatic fibrosis deposition in db/db mice **(Figures 4H-K)**. Liver triglycerides (TG), serum TG and total cholesterol (TC) levels were significantly decreased, whereas liver TC level was not significantly changed by iER intervention **(Figures S5D-S5G)**. Serum ALT and AST levels, indicators of liver damage, were significantly reduced in db/db mice following iER intervention **(Figures 4L-M)**. The iER increased AKT phosphorylation at Ser473, indicating that it enhanced the hepatic insulin signaling in db/db mice **(Figures 4O-4P)**. Consistent with clinical findings, iER reduced serum levels of the pro-inflammatory factor LPS in db/db mice **(Figures 4N)**. Moreover, iER significantly reduced the expressions of IL-1β and IL-18 in the liver of db/db mice by Western blotting detection, thereby improving hepatic inflammation **(Figures 4O-4P)**. Altogether, these results suggest that iER alleviates hyperglycemia, insulin resistance, hepatic steatosis and hepatic inflammation in db/db mice, exerting its robust hepatometabolic protective effects.

**Figure 4.**
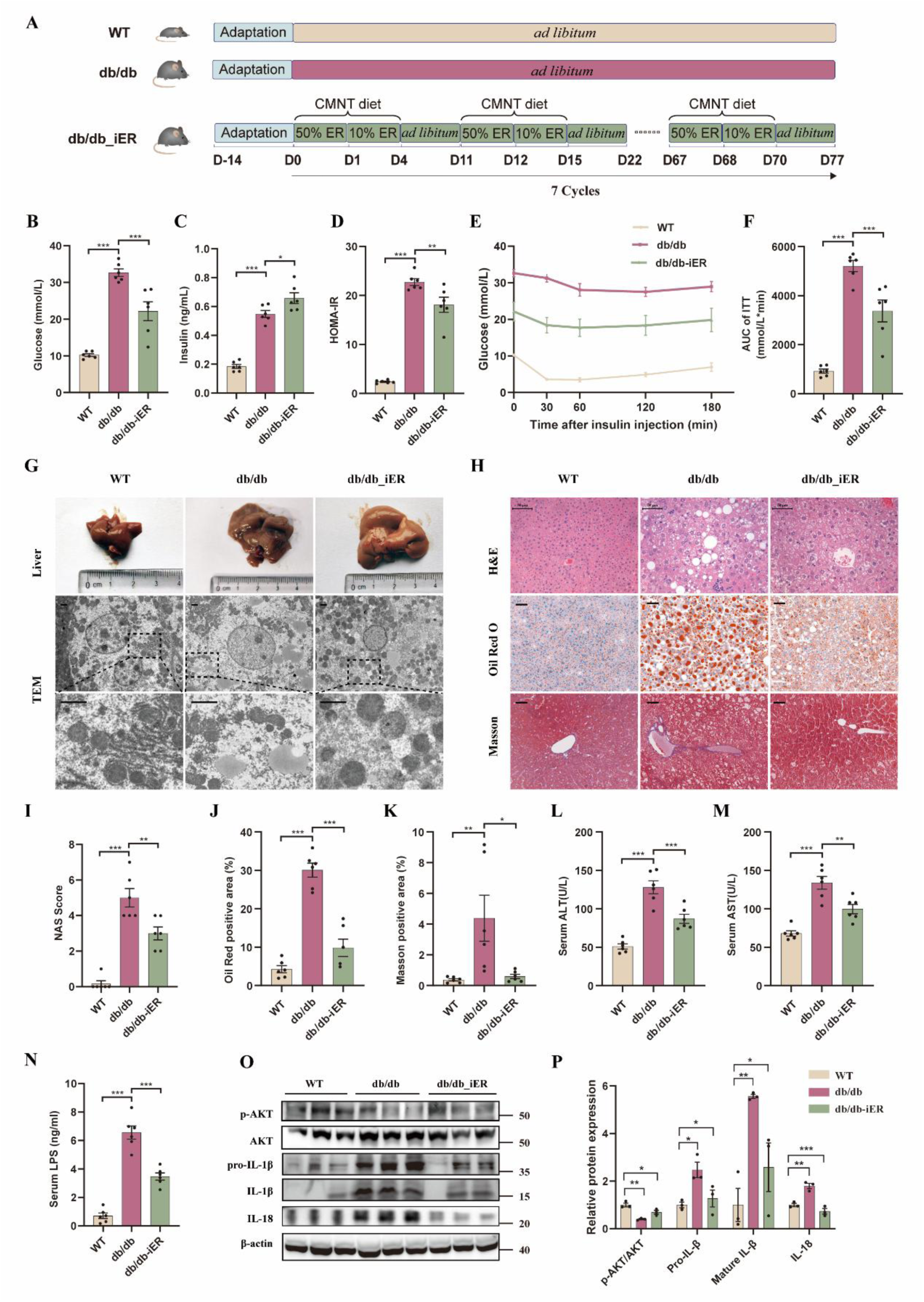
The iER reduces hyperglycinemia, insulin resistance, hepatic steatosis and liver inflammation in db/db mice. **(A)** Experimental scheme for assessing metabolic protective effects of iER in db/db mice. Twelve-week-old male db/db mice were randomly divided into two groups (n = 6). The db/db_ iER group mice adhered to an intermittent energy restriction (iER) regimen. Each iER cycle entails low-calorie Chinese Medical Nutrition Therapy (CMNT) diet, a medicinal plant-based diet, for 4 days (provided at 50% and 10% of normal daily intake, respectively), followed by 7 days of *ad libitum* feeding with standard mouse chow. This cycle was repeated 7 times. The db/db group received a regular chow diet *ad libitum*. Age-matched BKs wild-type male mice were used as a normal control group (n = 6). **(B)** Fasting glucose. **(C)** Fasting insulin. **(D)** HOMA-IR. **(E)** Insulin-tolerance test (ITT). **(F)** AUC for blood glucose in ITT. **(G)** Representative liver gross morphology and TEM images from the liver of mice (n = 6). Scale bar, 1.25 μm. **(H)** Representative images of H&E-stained, ORO-stained and Masson-stained liver sections of mice (n = 6). Scale bar, 50 μm. **(I-K)** Quantification of liver sections. **(L-M)** Serum ALT and AST. (N) Serum LPS. **(O-P)** Western blots analysis on the expression of p-AKT/AKT, pro-IL-1β, mature-IL-1β and IL-18 proteins in the liver of db/db mice (n = 3 mice per group). The relative intensities of proteins were normalized against β-actin. Data are shown as mean ± SEM. Statistical analyses were performed using one-way ANOVA with Tukey’s multiple comparison test. *p < 0.05, **p < 0.01, ***p < 0.001. See also **Figure S5**. Source Data are provided as a Source Data file.

### The iER regulates gut microbiota composition in db/db mice

To assess the protective effects of iER on intestinal injury, histopathological analyses were performed in colon and ileum tissues. HE staining showed that broken colon and ileum villi were observed in db/db mice, along with massive inflammatory infiltration in the lamina propria and crypts, and some crypts were noted to be missing. However, such disruption was reversed by the iER intervention and closer to the normal state **(Figures 5A-5B)**.

**Figure 5.**
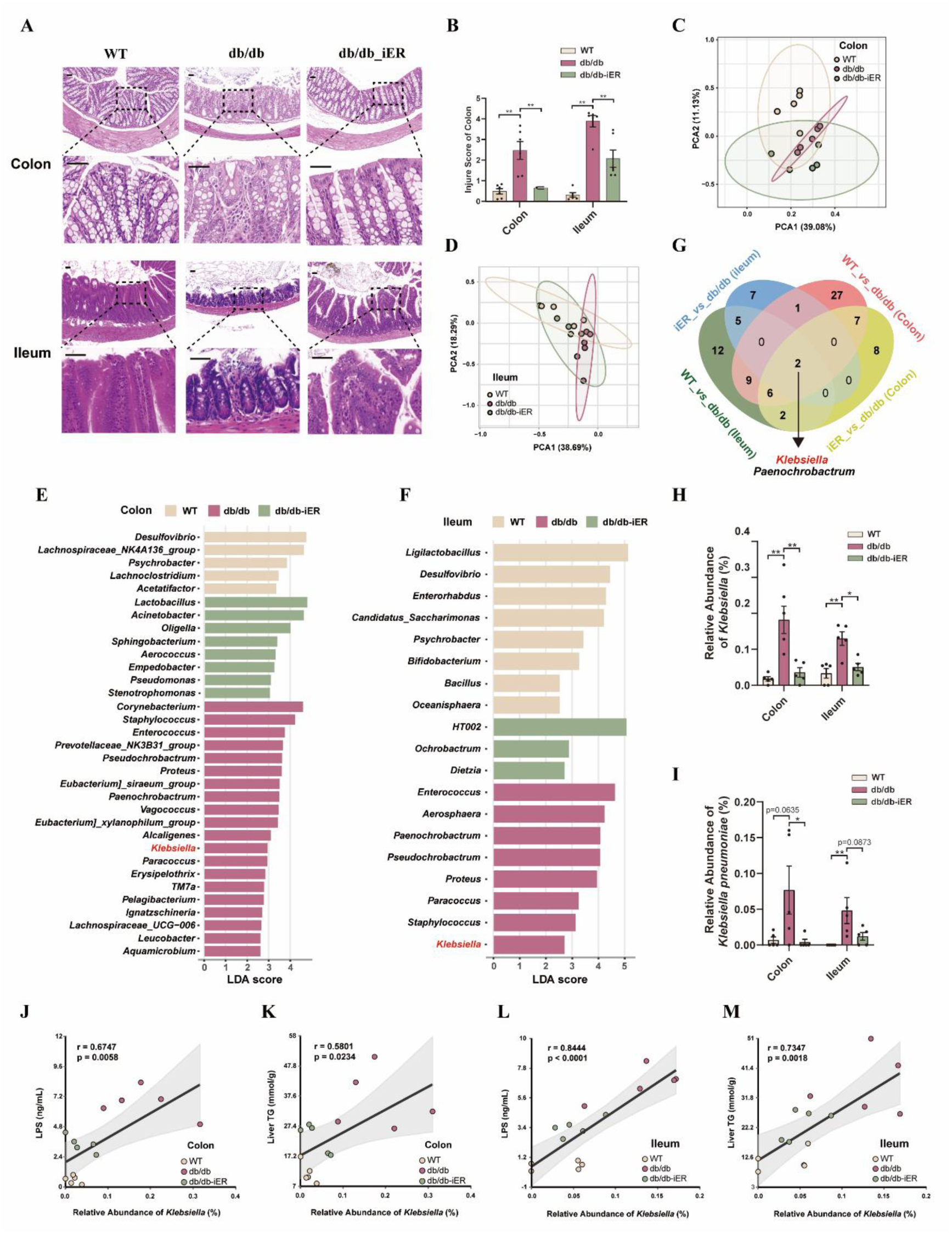
The iER attenuates gut dysbiosis in db/db mice. (A-B) Representative images of H&E staining (A), and quantification (B) for the colon and ileum sections from mice (n = 6 per group). Scale bar: 25 µm. **(C-D)** PCA plot of microbiota composition in the colon and ileum contents using Bray–Curtis distance metrics. **(E-F)** The difference of genus-level relative abundance by LEfSe in the colon and ileum contents of mice (p < 0.05 and LDA > 2.5). **(G)** Venn diagram showing the overlap of differential genera from comparisons of different groups in colon and ileum contents. **(H-I)** Changes in the relative abundance of the genus *Klebsiella* and the Enterobacteriaceae species *K. pneumoniae* in the colon and ileum contents of mice in WT, db/db and db/db_iER groups (n=5 per group). **(J-M)** Spearman’s correlations between the relative abundance of the genus *Klebsiella* in the colon contents and serum LPS or liver TG, and between that in the ileum contents and serum LPS or liver TG, in mice. r, Spearman’s correlation coefficient, and p < 0.05 was considered statistically significant. Data are shown as mean ± SEM. The p-values were calculated using one-way ANOVA with Tukey’s multiple comparison test (B) and Kruskal–Wallis test with post hoc Dunn’s multiple comparison test (H and I). *p < 0.05, **p < 0.01, ***p < 0.001. See also **Figure S6.** Source Data are provided as a Source Data file.

To determine whether iER-induced gut microbiota changes in mice mirror those in human samples, we further profiled the compositional alterations of gut microbiota in the mice colon and ileum contents in response to iER intervention through 16S rRNA gene sequencing **(Figure S6A)**. Compared with the db/db group, iER exhibited no significant differences in the α-diversity index, as reflected by the Chao1 index **(Figures S6B-6C)**. PCA revealed significant structural changes of gut microbiota in colon and ileum contents of mice after iER intervention **(Figures 5C-5D)**. Taxonomic profiling demonstrated significant differences at phylum and genus levels **(Figures S6D-6G)**. Through Linear Discriminant Analysis (LDA) in colon and ileum contents, we identified several genera with significant differences in abundance among the WT, db/db, and db/db_iER groups. Notably, *Klebsiella*, as a signature genus identified in human samples in this study, emerged as one of the most significant taxonomic markers distinguishing between db/db_iER group and db/db group **(Figures 5E-5F)**. The Venn diagram analysis of differential genera in colon and ileum contents revealed two significant genera, *Klebsiella* and *Paenochrobactrum*, when comparing the WT, db/db, and db/db_iER groups **(Figure 5G)**. Consistent with outcomes of human clinical trials, iER intervention significantly decreased the abundance of genus *Klebsiella* in colon and ileum contents compared with the db/db group **(Figure 5H)**. The species *K. pneumoniae* was significantly reduced in colon contents (p < 0.05) and showed a decreasing trend in ileum contents (p = 0.2021) following iER intervention **(Figure 5I)**. Spearman’s correlation analysis revealed relative abundance *Klebsiella* in colon and ileum contents was positively correlated with glucose, liver TG, HOMA-IR and serum LPS, respectively **(Figures 5J-5M and S6H-6K)**. These results suggest that iER modulates the gut microbiota composition, particularly through significant reduction of *Klebsiella* abundance, which can be related to improvements in glycemic control and hepatometabolic function.

### Multi-omics analysis identifies HADHA as the core target of iER in liver of db/db mice

To systematically identify the metabolic pathways and molecular targets mediating iER’s hepatoprotective effects in db/db mice, we performed transcriptomic, proteomic and acetylomic analyses of liver tissues with or without iER intervention **(Figure 6A)**. Compared with the db/db group, db/db_iER group exhibited extensive alterations in lipid metabolism at both the mRNA and protein levels, implicating this pathway as a primary mediator of iER’s benefits. Specifically, PCA analysis highlighted strikingly distinctions between the two groups **(Figures S7A and S8A)**. Transcriptome analysis identified 1507 differentially expressed genes (DEGs) (absolute fold-change ≥1.5, p < 0.05) in the db/db_iER group compared with the db/db group, including 742 upregulated genes and 765 downregulated genes **(Figure S7B)**. Proteomic analysis identified 30 differentially expressed proteins (DEPs) (absolute fold-change ≥1.2, p < 0.05), including 19 upregulated proteins and 11 downregulated proteins **(Figures S8B-S8C)**. Kyoto Encyclopedia of Genes and Genomes (KEGG) analysis of the transcriptomic data showed that upregulated genes were enriched in pathways related to glycerolipid metabolism, fatty acid degradation, PI3K-Akt signaling pathway, and PPAR signaling pathway, whereas downregulated genes were enriched in pathway related to Non-alcoholic fatty liver disease **(Figures S7C-S7D)**. Lipid metabolism-related differentially expressed genes (DEGs) transcripts were manually enriched, and presented in the heatmap **(Figure S7E)**. The nine modulated lipid metabolism functions associated with 77 DEGs, which were screened by KEGG pathway enrichment analysis. We then profiled the lipid metabolism network of the functions and DEGs using KEGG pathway enrichment analysis **(Figure S7F)**. KEGG analysis of the proteomic data demonstrated that DEPs enriched in pathways related to fatty acid metabolism, PPAR signaling pathway and Non-alcoholic fatty liver disease **(Figure S8D)**. The KEGG and GO gene sets also showed that the differential proteins were mostly enriched in these lipid metabolism pathways **(Figures S8E-8I)**. The PPI results showed that the major interacting differentially expressed proteins were Hadha, Acsl1, Acsf2 and Crat.

**Figure 6.**
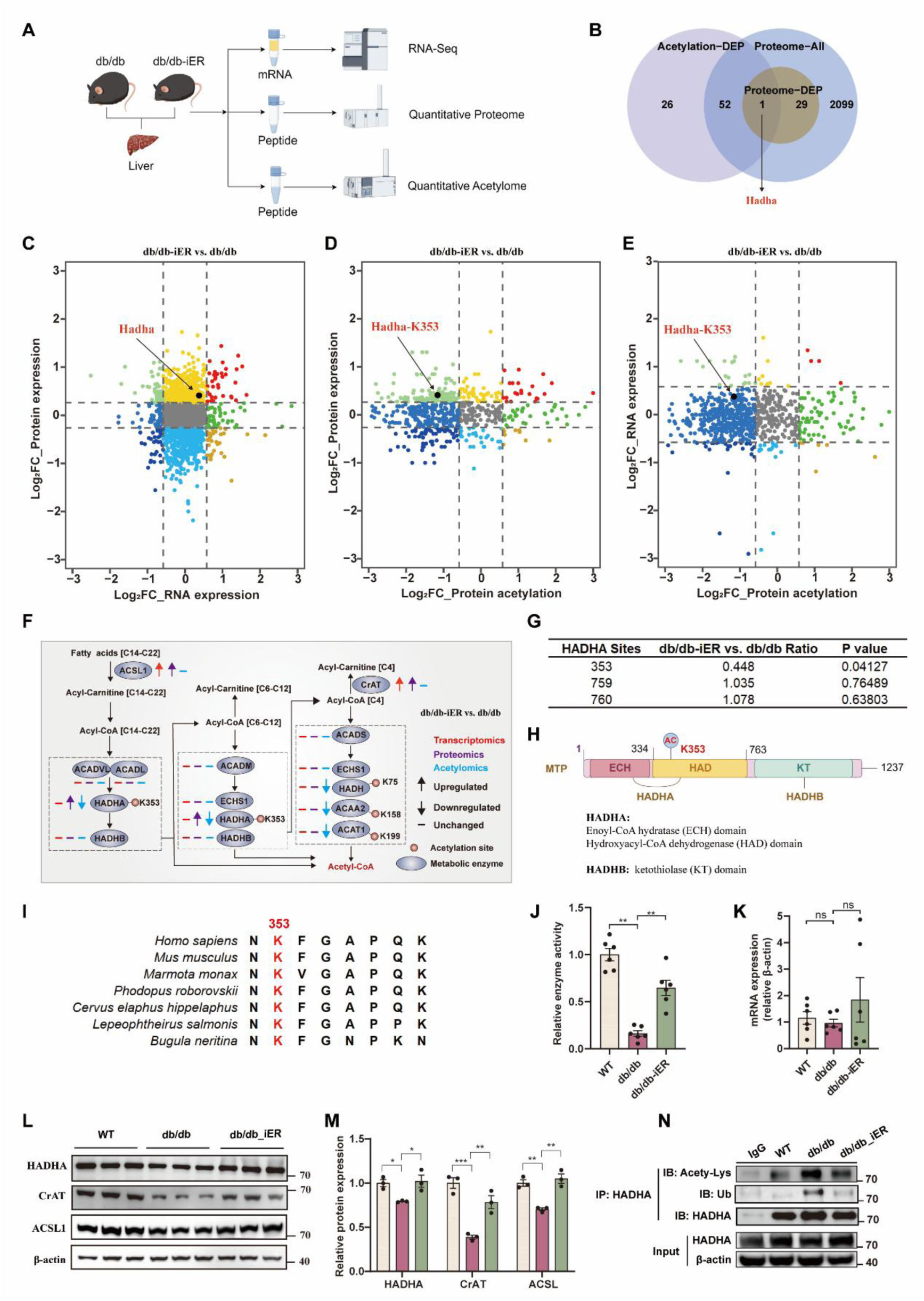
Multi-omics analysis identifies HADHA is identified as a core target by iER intervention in db/db mice. **(A)** A Schematic diagram showing the experimental workflow of the core target identification by multi-omics in the liver of db/db mice. **(B)** Venn diagram showing the overlap of differential expression acetylated proteins and all identified protein and DEPs (protein), respectively. n = 3 mice per group for acetylome. n = 4 mice per group for proteome. **(C-E)** Intersection analysis of transcriptome, proteome and acetylome showing genes or proteins with significant change in mRNA expression, protein expression and acetylation level in the liver of db/db mice after iER intervention. **(F)** Metabolic enzymes in fatty acid β-oxidation pathways. The upregulated, downregulated and unchanged states of genes or proteins in db/db_iER versus db/db mice liver are indicated at mRNA, protein and acetylation levels. **(G)** The ratio and p value of acetylated HADHA on different sites under db/db_iER versus db/db from acetylome. **(H)** Domain pattern of HADHA, the linear structure location of the K353 site and the physiological function of the domain. **(I)** Conservation of HADHA-K353 acetylated sites in multiple organisms. **(J)** Relative enzyme activity of HADHA in the liver of db/db mice. **(K-M)** The expression of HADHA at the mRNA and protein expression level in the liver of db/db mice. **(N)** Immunoprecipitation was performed to detect acetylated and ubiquitylated HADHA in the liver of db/db mice. Abbreviations: DEP, differentially expressed proteins; FC, fold change. Data are shown as mean ± SEM. The p-values were calculated using one-way ANOVA with Tukey’s multiple comparison test. *p < 0.05, **p < 0.01, ***p < 0.001. See also **Figure S7-S9**. Source Data are provided as a Source Data file.

Notably, iER increased protein expression of carnitine acetyltransferase (CrAT, acetyl group transfer) **(Figure 6L)**, an enzyme critical for acetyl-CoA/acetylcarnitine interconversion and metabolic flexibility[37]. Since acetyl-CoA serves as the direct donor for protein acetylation, CrAT-mediated control of acetyl-CoA pools may modulate acetylation-dependent pathways. Thus, we hypothesized that iER could regulate hepatic metabolism through protein acetylation. Protein lysine acetylation is a reversible and dynamic post-translational modification that regulates cellular metabolism, which has been linked to metabolic dysfunction[38]. The iER markedly decreased global acetylation levels, but did not alter the levels of ubiquitination **(Figure S9A)**. To gain further insight into the effects of iER on hepatic metabolism, we performed acetylomics analysis of total liver proteins from db/db mice. PCA analysis showed a clear separation between the db/db_iER and db/db groups **(Figure S9B)**. Lysine acetylomics analysis identified 101 differentially regulated acetylated sites in 79 proteins (absolute fold-change ≥1.2, p < 0.05) including 99 upregulated acetylated sites in 77 proteins and 2 downregulated acetylated sites in 2 proteins **(Figures S9C and S9E)**.KEGG analysis revealed pathways enriched in fatty acid degradation **(Figure S9F)**.

The overlaps among all identified genes, proteins, and acetylated proteins were presented in **Figure S9D**. Multi-omics data indicate that the fatty acid β-oxidation pathway is a key metabolic process that is strongly altered after iER intervention. By comparing proteomic and acetylomic data, we found that only one metabolic enzyme, HADHA, was markedly changed at both levels, indicating that the proteome and lysine acetylome might be differentially shaped by the iER intervention, with HADHA emerging as a core molecular target of iER **(Figure 6B)**. Protein-protein interaction (PPI) analysis of the proteome and acetylome revealed that HADHA formed the central node of an interconnected protein module, highlighting its potential importance in metabolic regulation **(Figures S8J and S9G-9H)**. Intersection analysis of transcriptomics, proteomics and acetylomics revealed that iER increased HADHA protein expression and reduced HADHA-K353 acetylation, lysine but did not affect the transcriptional expression level **(Figures 6C-6E)**. Given that the ubiquitin-proteasome system is the most common pathway for degradation of cellular proteins,[39] we speculated that iER induced HADHA deacetylation modification increases HADHA protein stability by inhibiting its ubiquitination-degradation.

We curated 12 metabolic enzymes related to fatty acid β-oxidation pathways based on our multi-omics data **(Figure 6F)**. Compared with the db/db group, iER intervention did not significantly alter the mRNA or protein expression levels of most enzymes, except for HADHA, ACSL (fatty acid activation), and CrAT, which were upregulated only at the protein level. However, the acetylation modifications of several fatty acid β-oxidation enzymes (including HADHA, ACAA2, HADH and ACAT1) were significantly downregulated. Since the abundance of most enzymes remained unchanged, these acetylation changes might functionally regulate their catalytic activity and/or substrate binding. To investigate the functional acetylation sites in HADHA, MS/MS spectra of HADHA was extracted from the acetylome data set. The results showed that lysine (K) 353 is the main acetylation site in HADHA **(Figure S9I)**. The acetylation data indicated a reduction in HADHA K353ac by 0.448-fold through iER intervention, while other identified modification sites including K759ac and K760ac did not exhibit a significant change **(Figure 6G)**. Existing PTM annotations in PhosphoSitePlus revealed dual acetylation and ubiquitination at HADHA K353[40], implicating its potential regulatory role in metabolic control **(Figure S9J)**. HADHA, the α-subunit of mitochondrial trifunctional protein (MTP), forms a functional complex with HADHB (β-subunit) to catalyze long-chain fatty acyl-CoA β-oxidation. Structurally, HADHA contains two catalytic domains: an enoyl-CoA hydratase (ECH) domain and a hydroxyacyl-CoA dehydrogenase (HAD) domain[41]. K353, located within the HAD domain (residues 334-763), mediates the dehydrogenation of long-chain 3-hydroxyacyl-CoA to 3-ketoacyl-CoA, coupled with NADH production **(Figure 6H)**. We analyzed the conservation of HADHA-K353 amino acid residues across different species and observed that all three sites were highly conserved through evolution **(Figure 6I)**. To validate the iER-reduced changes of HADHA at different levels in mouse liver, we conducted qPCR, Western blot, Co-immunoprecipitation (Co-IP) and enzyme activity analyses. Consistent with results of multi-omics, iER downregulated HADHA protein levels, without affecting HADHA mRNA expression. Importantly, iER dramatically lowered the lysine deacetylation of HADHA and enhanced HADHA relative enzyme activity **(Figures 6J-6N)**. These results suggest that iER-induced HADHA deacetylation at K353 site is likely linked to hepatometabolic improvements.

### HADHA protects against LPS combined with HGHF-induced fat accumulation, insulin resistance and inflammation in hepatocytes

HADHA protein expression in MIHA cells was not significantly altered by *K. pneumonia-derived* or *E. coli* derived LPS treatment, but *K. pneumonia-derived* LPS further decreased HADHA protein expression in HGHF-induced MIHA cells **(Figures S10A-10D)**. Mirroring db/db mouse liver data, *K. pneumonia-derived* LPS and HGHF co-treatments downregulated HADHA protein expression (without significant changes in HADHA mRNA levels) and enzyme activity, decreased CrAT, ACSL1 and p-AKT protein expression, and increased pro-IL-1β, mature IL-1β, and IL-18 protein expression **(Figures 7A-7B and S10E-10F)**. *K. pneumonia-derived* LPS combined with HGHF increased global acetylation levels, but did not alter ubiquitination levels in MIHA cells **(Figure S10G)**. To determine the effect of acetylation on HADHA activity in hepatocytes, we treated MIHA cells with the lysine deacetylase inhibitors (KDACi) nicotinamide (NAM, an inhibitor of nicotinamide adenine dinucleotide (NAD^+^)-dependent sirtuins) and suberoylanilide hydroxamic acid (SAHA, an inhibitor of Zn^2+^-dependent deacetylases). HADHA activity in cell lysates was not significantly changed by treatment with SAHA, but treatment with NAM or NAM and SAHA was significantly more potent relative to SAHA alone, suggesting that HADHA activity was modulated by lysine acetylation in MIHA cells **(Figure 7C)**. Co-IP assay showed *K. pneumonia-derived* LPS and HGHF co-treatments increased HADHA acetylation **(Figure 7D)**. These results suggest that *K. pneumonia-derived* LPS combined with HGHF induces fat accumulation, insulin resistance and inflammation, and the *in vitro* comorbid MASLD and T2DM model is successfully established in MIHA cells.

**Figure 7.**
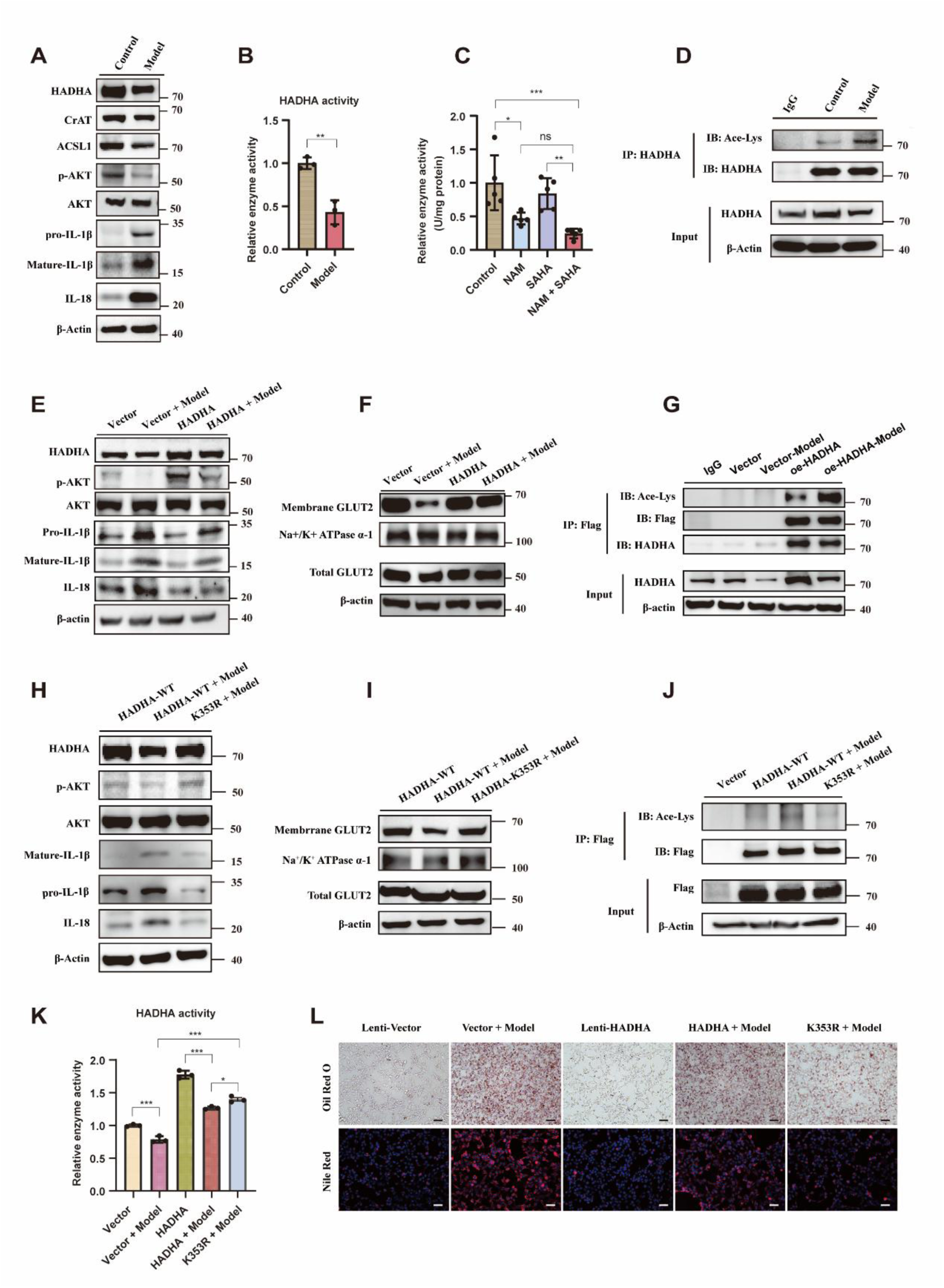
The effect of HADHA and its mutant HADHA-K353R on lipid deposition, insulin resistance and inflammation in HGHF combine with LPS induced MIHA cells. **(A)** Western blotting analysis of the expression of p-AKT, AKT, pro-IL-1β, IL-18 and HADHA in LPS combined with HGHF-induced MIHA cells. **(B)** Relative HADHA enzyme activity in MIHA cells upon HGHF + LPS treatment. **(C)** Relative HADHA enzyme activity in MIHA cells cultured in the presence or absence of the lysine deacetylase inhibitors (KDACi). **(D)** Co-IP assay was performed to detect acetylated HADHA in MIHA cells upon HGHF + LPS treatment. **(E-F)** Western blotting analysis of the expression of p-AKT, AKT, pro-IL-1β, IL-18, HADHA and GLUT2 in HGHF and LPS induced MIHA cells transfected with overexpressed HADHA (oe-HADHA). **(G)** Co-IP assay was performed to detect the expression of exogenous acetylated HADHA in oe-HADHA MIHA cells with or without HGHF + LPS treated treatment. **(H-I)** Western blotting analysis of the expression of p-AKT, AKT, pro-IL-1β, IL-18, HADHA and GLUT2 in oe-HADHA-WT and oe-HADHA-K353R MIHA cells with or without LPS+ HGHF treatment. **(J)** Co-IP assay was performed to detect the expression of acetylated HADHA in oe-HADHA-WT and oe-HADHA-K353R MIHA cells with or without LPS+ HGHF treatment. **(K)** Relative HADHA enzyme activity was determined in oe-HADHA-K353R MIHA cells with or without HGHF + LPS treatment. **(L)** Representative images of Oil Red O staining and Nile Red staining of lipid droplets accumulation were detected in oe-HADHA-WT and oe-HADHA-K353R MIHA cells with or without HGHF + LPS treatment. The empty lentiviral vector (Lenti-Vector) was generated as a negative control. Scale bar, 50 μm. Data are shown as mean ± SEM. Statistical analysis was performed using two-tailed unpaired Student’s t test for two group comparisons or one-way ANOVA followed by Tukey’s multiple comparison test for multiple comparisons between groups. *p < 0.05, **p < 0.01, ***p < 0.001, ns, no significance. See also **Figures S10-S12**. Source Data are provided as a Source Data file.

To further investigate the role of HADHA on hepatic steatosis, insulin resistance and inflammation, we examined these effects of HADHA on *K. pneumonia-derived* LPS combined with HGHF-induced MIHA cells using genetic approaches. HADHA was cloned into a lentiviral vector and stably transduced with MIHA cells **(Figures S11A-S11B)**. The ectopic protein expression of HADHA was significantly increased **(Figure S11C)**. HADHA overexpression remarkedly enhanced insulin-stimulated phosphorylation of AKT at Ser473 and GLUT2 plasma membrane translocation, while suppressing the protein expression of pro-inflammatory cytokines IL-1β and IL-18 in LPS combined with HGHF-induced MIHA cells **(Figures 7E-7F and S11D-S11E)**. *K. pneumonia-derived* LPS and HGHF co-treatments increased HADHA acetylation, reduced its enzyme activity and attenuated lipid accumulation in HADHA overexpression-transfected MIHA cells **(Figures 7G and 7K-L)**. In addition, the endogenous HADHA protein expression was selectively reduced by HADHA-specific siRNA transfection. Compared with scrambled siRNA, knockdown of HADHA in MIHA cells significantly attenuated insulin-stimulated AKT phosphorylation at Ser473 and GLUT2 plasma membrane translocation, and elevated the expression of inflammatory factors IL-1β and IL-18 under LPS combined with HGHF treatment **(Figures S11F-S11I)**. Taken together, these results indicate that HADHA overexpression reduces *K. pneumonia-derived* LPS combined with HGHF-induced hepatocellular steatosis, insulin resistance and inflammation in hepatocytes, whereas HADHA knockdown reverses these effects.

### HADHA-K353 deacetylation enhances HADHA enzyme activity and stability, which alleviates hepatic fat accumulation and insulin resistance in LPS combined with HGHF-induced MIHA cells

To understand the metabolic role of deacetylated HADHA-K353, we mutated lysine (K) 353 to arginine, which bears the same charge as lysine but cannot be acetylated **(Figures S12A-12B)**. The deacetylation of HADHA-K353R was confirmed by Co-IP assay **(Figure S12C)**. MIHA cells transfected with FLAG-tagged HADHA mutant expression plasmids (HADHA-K353R) significantly increased HADHA enzyme activity **(Figure S12D)**. HADHA-K353R overexpression, which mimicked iER intervention in db/db mice, displayed significantly decreased HADHA acetylation, elevated HADHA enzyme activity, dramatically reduced cellular lipid accumulation, and enhanced insulin-stimulated AKT phosphorylation at Ser473 as well as GLUT2 plasma membrane translocation in *K. pneumoniae*-derived LPS combined with HGHF-induced MIHA cells compared with the HADHA-WT **(Figures 7I-7L and Figure S12E-H)**. Taken together, these results indicate that HADHA-K353 deacetylation enhances enzymatic activity, stability, inhibit lipid deposition and mitigate insulin resistance in *K. pneumonia-derived* LPS combined with HGHF-induced hepatocytes.

### HADHA-K353 deacetylation blocks its binding to inflammasome adaptor ASC

Considering that iER reduced *K. pneumonia* and blood LPS levels, inhibited inflammatory cytokine expression and secretion, and decreased HADHA-K353 acetylation. A previous study has reported that HADHA acetylation in cardiomyocytes can recruit NLRP3 inflammasome adaptor apoptosis-associated speck-like protein containing a CARD (ASC), which translocates to the mitochondria, triggering the activation of the NLRP3 inflammasome[42]. Therefore, we hypothesized that HADHA-K353 deacetylation blocks its binding to ASC, thereby inhibiting hepatic inflammatory signaling activation in comorbid MASLD and T2DM. We found that HADHA at K353 site can bind to ASC (docking score = −199.74) by molecular docking analysis (**Figure 8A**). An increased protein expression of mitochondrial ASC was observed in both *K. pneumonia-derived* LPS combined with HGHF-induced MIHA cells and db/db mice. iER intervention reduced mitochondrial ASC protein expression in db/db mice (**Figures 8B-E**). Co-IP assay confirmed that HADHA physically interacted with ASC in LPS combined with HGHF induced WT-MIHA cells and overexpression-HADHA MIHA cells, and the liver of db/db mice, whereas iER intervention reduced interaction between HADHA and ASC in the liver of db/db mice (**Figures 8F-8H**). To interrogate whether HADHA deacetylation at K353 site block its binding to ASC, HADHA-K353R structure was predicted by AlphaFold2. HADHA-K353R mutant displayed a lower binding energy for ASC (docking score = –167.94) and induced conformational change in the ASC structure compared with the wild-type (WT) counterpart **(Figures 8I and S12I**). Consistent with the results from the molecular docking analysis, Co-IP results showed that mutation of HADHA-K353R led to reduced interaction with ASC compared with the WT counterpart in LPS combined with HGHF induced MIHA cells **(Figure 8J)**. Collectively, these data suggest that HADHA-K353 deacetylation blocks its binding to ASC, which can contribute to inhibit inflammatory cytokine expression.

**Figure 8.**
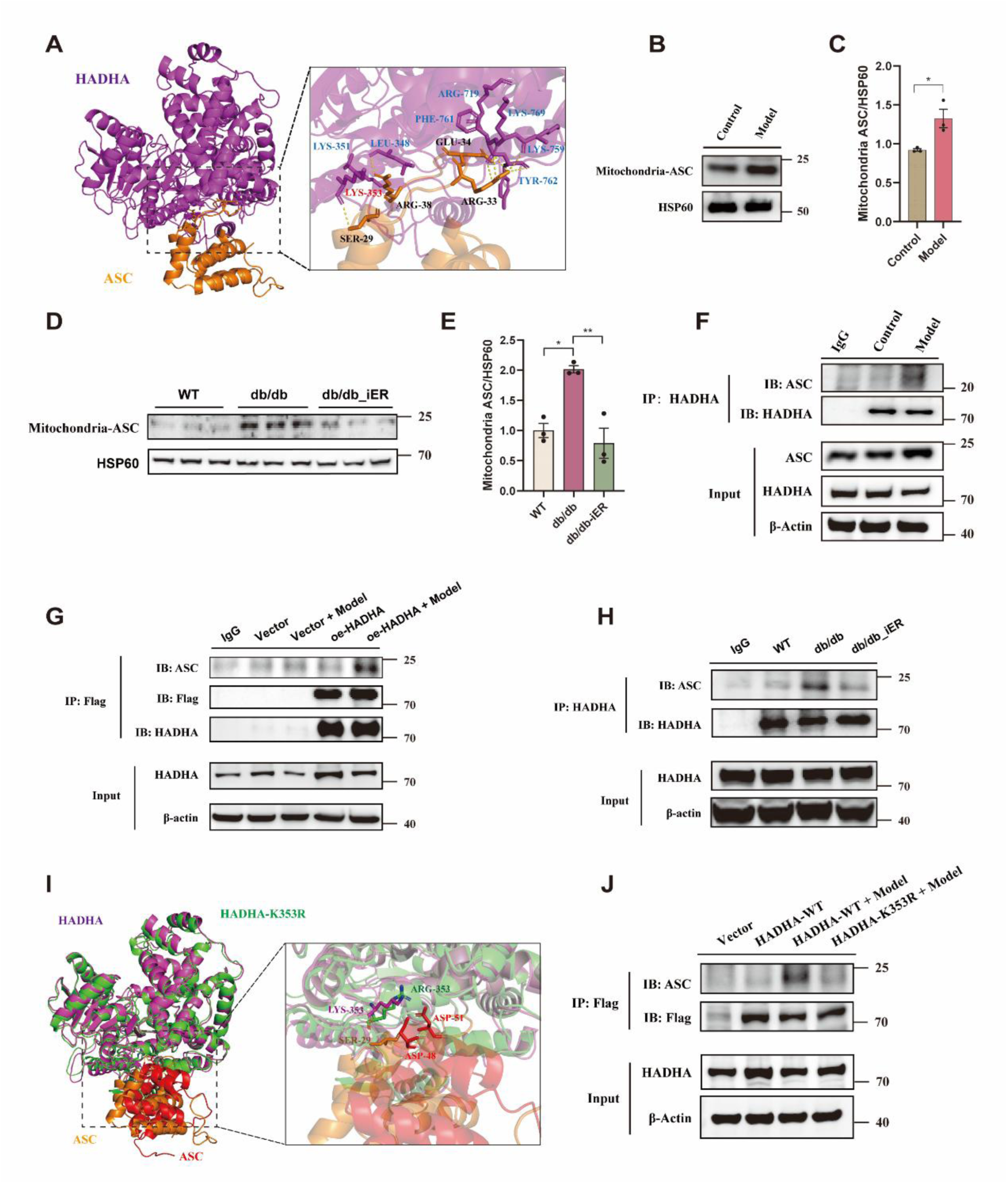
HADHA-K353 deacetylation blocks its binding to ASC, inhibiting the activation of inflammatory signaling in HGHF combined with LPS induced hepatocytes. **(A)** Structure diagram showing the docking model of HADHA (purple)-ASC (brown) complex. **(B-E)** Western blotting analysis of the differential expression of isolated mitochondria ASC from the liver of mice and MIHA cells. Mitochondrial Hsp60 served as loading control. **(F-H)** Co-IP analysis to detect interaction between HADHA and ASC in HGHF combined with LPS induced MIHA cells, oe-HADHA MIHA cells with or without HGHF + LPS treatment and mouse liver with or without iER intervention. **(I)** Conformational change in the ASC structure induced by HADHA-K353R mutant. The predicted HADHA-K353R structure was represented as green ribbons by AlphaFold2 and native HADHA-WT structure was represented as purple ribbons. **(J)** Co-IP analysis to detect interaction between HADHA or HADHA-K353R and ASC in HGHF combined with LPS induced MIHA cells. See also **Figure S12**. Source Data are provided as a Source Data file.

## Discussion

In this study, we demonstrate that an iER regimen improves glycemic control, reduces hepatic steatosis, insulin resistance and inflammation in comorbid MASLD and T2DM. To our knowledge, this is the first study to specifically evaluate the impacts of iER on glucolipid metabolism in MASLD patient with comorbid T2DM, while also exploring the gut-liver axis mechanisms involved. To date, only a limited number of studies have investigated the impact of iER on liver steatosis and glycemic control in populations with MASLD or T2DM. A 5:2 diet regimen, a form of iER, has also been found efficacy on reduction in liver fat or glycemic control in patients with T2DM[16, 43, 44]. An alternate-day energy restriction diet, another form of iER, reduces liver steatosis by ultrasonography in patients with MASLD[45]. A fasting-mimicking diet, a generalized form of iER, reduces liver fat and liver inflammation measured by magnetic resonance imaging in patients with T2DM[46]. Although these findings suggest the clinical effectiveness of iER, the mechanisms involved remained largely unknown. Our study revealed that the *K. pneumoniae*/LPS/hepatic HADHA-K353 acetylation axis mediates the beneficial effects of iER on comorbid MASLD and T2DM, representing a novel mechanism.

We found that iER reduced levels of the phylum Pseudomonadota, the class Gammaproteobacteria, the order Enterobacterales, the family Enterobacteriaceae, the genus *Klebsiella*, and the species *K. pneumoniae* in patients with MASLD and T2DM by metagenomic analyses. Notably, these taxa represented the top 20 most abundant bacteria and were the only differentially abundant taxa maintained across all taxonomic levels within this dominant group. *K. pneumoniae*, a conditional pathogenic bacterium that can colonize the intestines, is enriched in multiple metabolic diseases, including inflammatory bowel diseases, hypertension and chronic spontaneous urticaria[47, 48, 49]. Two studies recently found that *K. pneumoniae* enrichment was observed in a small Chinese cohort of patients with T2DM or hypertension patients with T2DM[50, 51]. Another previous study reported that high alcohol-producing *K. pneumoniae* strain was associated with up to 60% of MASLD cases in a small Chinese cohort and contributed to fatty liver disease progression, while non-alcohol-producing and low-alcohol-producing *K. pneumoniae* strains were also detected in this population, indicating strain-specific pathogenicity[52]. Consistent with these findings, other clinical evidence also demonstrated that *K. pneumoniae* exacerbated disease progression in a strain-specific manner across diverse pathological conditions[53, 54, 55]. Therefore, we speculated that the specific strain from comorbid MASLD and T2DM has unique characteristics that make it particularly pathogenic compared with other *K. pneumoniae* strains. We isolated a *K. pneumoniae strain* (ZH2.1) from fecal sample of a patient with MASLD and T2DM. A comparative genomic analysis revealed that the strain ZH2.1 exhibited distinct variations in genes associated with LPS synthesis and modification compared with other *K. pneumoniae* or *E. coli* strains. Furthermore, the strain ZH2.1 showed unique characteristics that render it particularly differential cytotoxic and inflammatory compared with other *K. pneumoniae* strains.

A subsequent question is how iER-induced a decrease in gut pathogen *K. pneumoniae* contributed to the hepatometabolic improvements. It is well established that gut microbiota influences liver disease progression by regulating changes in intestinal metabolites or directly translocating the host organs[26, 53]. In this study, functional analysis of metagenomic shotgun sequencing revealed changes in LPS biosynthetic pathways after iER intervention. Reduced serum LPS levels are closely associated with the decrease of *K. pneumoniae*, which is linked to significant reductions in hyperglycemia, hepatic steatosis, insulin resistance, and inflammation in patients with MASLD and T2DM following iER intervention. Our findings are similar to previous report that *K. pneumoniae* can exacerbate MASLD progression by increasing LPS production[33]. Except for *K. pneumoniae*, other Gram-negative bacteria producing LPS with different endotoxin activity levels showed different capacities to induce MASLD[33, 56]. It is interesting to note that LPS from different *E. coli* strains (e.g., O55:B5, O111:B4, O127:B8, and O128:B12) exhibits differential proinflammatory activities and other biologic activities[57, 58]. These functional variations likely arise from strain-specific LPS structures among bacterial strains. LPS is a complex biomacromolecule consisting of three structural components: O-antigen, a core oligosaccharide, and lipid A. The O-antigen is recognized by the host immune system, while lipid A is responsible for the endotoxic activity and inflammatory properties of LPS[59]. The role of LPS in the regulation of inflammation and immunity is complex. For example, the *Proteobacteria*-derived LPS exhibits a heightened capacity to activate the TLR4 receptor in comparison to that from *Bacteroides*, but it can also cause LPS tolerance. Thus, it cannot be conclusively determined that greater proportion of *Proteobacteria* is necessarily a driver of inflammation[34]. Our study found that increased subclinical inflammation occurs in comorbid MASLD and T2DM, unlike acute bacterial infections which trigger substantial LPS release into the bloodstream[31]. LPS is regarded as a differential bacterial metabolite in a variety of chronic metabolic diseases, including MASLD and T2DM[7, 60, 61]. Given its structural diversity, it will be possible for LPS to develop diseases biomarkers in the future. In this study, the elevated blood LPS levels were observed in db/db mice, which may result from increased intestinal permeability, representing a chronic and progressive pathological process. iER decreased the abundance of *K. pneumoniae* and serum LPS levels, and increased the expression of intestinal tight junction proteins (ZO-1, Occludin and Claudin-1), thereby protecting the intestinal barrier integrity from intestinal macrophage infiltration and decreasing intestinal inflammation in db/db mice. Previous work found that LPS can modulate intestinal epithelial permeability and inflammation in a species-specific manner[62]. Therefore, a more nuanced understanding for the biological functions of LPS is required in the context of specific diseases. Therefore, we extracted *K. pneumoniae* derived LPS from clinical isolate (ZH2.1), which exhibited distinct cytotoxicity and compositional profiles compared with commercial *E. coli* derived LPS (L2880, Sigma), as demonstrated by CCK-8 assays and silver staining. These differences indicated that LPS bioactivity may be strains-specific, highlighting the importance of strain– and origin-dependent considerations when evaluating LPS-mediated inflammation responses.

The iER decreased the abundance of gut pathogen *K. pneumoniae*, which is strongly positively correlated with reductions in serum LPS levels. This suggests that *K. pneumoniae*-derived LPS may be a key metabolite exacerbating comorbid MASLD and T2DM, and that iER alleviates this comorbidity by reducing *K. pneumoniae* and its derived LPS. The current study also corroborates that *K. pneumoniae*-derived LPS exacerbates lipid accumulation, insulin resistance and inflammation in HGHF-indued hepatocytes. Another key question is how changes occurring in the gut affect hepatic metabolism by iER intervention in comorbid MASLD and T2DM. The potential relationship between bacterial metabolites and host PTMs represents an exciting avenue of research for deciphering the gut microbiota-host interactions. However, currently, there are limited reports available[20, 23]. Through a multi-omics approach, we identified HADHA, a core target of lysine acetylation that controls the rate-limiting step of hepatic fatty acid β-oxidation, as the mediator of iER-induced hepatometabolic improvements in db/db mice. HADHA, the α subunit of the mitochondrial trifunctional protein (MTPα), contains an enoyl-CoA hydratase (ECH) domain and a 3-hydroxyl-CoA dehydrogenase (HAD) domain. These domains catalyze the long-chain enoyl-CoA hydratase activity of the second step and the 3-hydroxyacyl-CoA dehydrogenase activity of the third step in fatty acid β-oxidation, respectively[41]. Heterozygous mice lacking HADHA had significantly reduced fatty acid β-oxidation in liver tissue and developed liver lipid deposition and inflammation[63]. In this study, we demonstrated that iER remarkably decreased hepatic HADHA acetylation, increased its protein level and enzyme activity, but had no significant effects on the transcription level. This suggests that iER-induced upregulation of HADHA expression occurs in a transcription-independent manner, and protein ubiquitination may be involved in this degradation process[39]. Co-IP assay confirmed that iER decreased hepatic HADHA ubiquitination, thereby stabilizing the protein in db/db mice. Moreover, our *in vitro* studies show that *K. pneumoniae*-derived LPS reduced HADHA protein level and enzyme activity, and increased its acetylation in HGHF-induced MIHA cells. HADHA overexpression alleviates accumulation of lipid droplets, insulin resistance and inflammation while knockdown HADHA results in the exact opposite changes in *K. pneumoniae*-derived LPS combined with HGHF-induced MIHA cells. These results suggest that HADHA regulates hepatometabolic protective effects. Mass spectrometry-based proteomics technology has uncovered that HADHA is acetylated at multiple sites. However, the effect and significance of individual acetylation sites are mostly unknown[64]. Our comparative acetylome study found that the acetylation of HADHA at the K353 site (This specific site had never been previously studied), a highly conserved lysine residue on HADHA across species, was decreased by iER intervention in the liver of db/db mice. The K353 site resides in the HAD domain (amino acids 334-763) of HADHA, which catalyzes the oxidation of long-chain 3-hydroxyacyl-CoA to 3-ketoacyl-CoA with concomitant NADH production. Notably, a highly debated issue is whether the enzyme activity of MTP is regulated by protein PTMs[65]. One important reason for the differential outcomes is the selectivity of the substrate, considering that MTP is a multifunctional enzyme system. To accurately reflect the changes in enzyme activity regulated at the K353 site, we have customized long-chain 3-hydroxypalmitoyl-CoA (C16) as the substrate, since it is not commercially available, to detect HADHA enzyme activity. Our results showed the acetylation of HADHA at the K353 site downregulated its enzyme activity. Furthermore, treatment of MIHA cells with deacetylase inhibitors (NAM and SAHA) found that HADHA activity was significantly altered by NAM (alone or combined with SAHA) but not by SAHA alone, suggesting lysine acetylation regulates HADHA enzyme activity. Site-directed mutagenesis on HADHA K353 is necessary in order to further validate its function. We found that an acetylation-null mutant (K353R) enhanced the HADHA enzyme activity and reduced lipid accumulation and insulin resistance in *K. pneumoniae*-derived LPS combined with HGHF-induced hepatocytes.

HADHA acetylation is involved in the regulation of multiple metabolic processes. it reduces fatty acid β-oxidation of islets and promotes palmitate-potentiated insulin secretion[66]. Conversely, HADHA deacetylation decreases insulin resistance in adipocytes[67]. It is worth noting that the acetylation modification at different lysine sites in HADHA may regulate different biological functions, and even lead to opposite regulatory outcomes[67, 68]. This has also been reported in other functional proteins[69]. Metabolizing enzymes, in addition to their canonical catalytic functions, also have a variety of non-metabolic roles. These functions include participation in signal regulation and acting as signaling molecules, playing a key role in diverse physiological and pathological processes[70, 71]. Accumulating evidence has indicated that metabolism and cellular signaling are inextricably connected, enabling the regulation of cellular processes, such as proliferation, differentiation, and inflammation, in response to the cellular metabolic status. Protein acetylation is associated with the acetyl-CoA/CoA ratio, whereas deacetylation relies on the NAD^+^/NADH ratio, indicating that acetylation could act as a metabolic regulator.

Our studies demonstrate that iER-induced HADHA deacetylation modulates its enzymatic activity, attenuates hepatic steatosis and regulates insulin resistance. An interesting question is whether direct HADHA acetylation could regulate hepatic inflammation signaling. Consistent with this speculation is that the acetylation of K255 on HADHA could recruit ASC, a component of the NLRP3 inflammasome, translocated to the mitochondria, which promotes the assembly of the NLRP3 inflammasome, thereby induces the release of inflammatory factors and leads to the occurrence of myocardial fibrosis in the heart tissue of obese mice[42]. Therefore, we further investigated HADHA-ASC interaction in both *in vitro* and *in vivo* models of comorbid MASLD and T2DM. Our studies confirmed that iER decreases HADHA interaction with ASC in the liver of db/db mice. *K. pneumoniae*-derived LPS combined with HGHF induces HADHA acetylation *in vitro* cell models. Mutation of K353 to arginine (K353R), which mimicked inhibition of HADHA-K353 acetylation modification, results in reduced HADHA interaction with ASC and suppresses the IL-1β and IL-18 expression, which in turn further decreases liver inflammation.

In summary, we investigated beneficial effects of iER on comorbid MASLD and T2DM from strain-specific Enterobacteriaceae *K. pneumoniae*, structural-specific LPS and site-specific hepatic HADHA-K353 acetylation and revealed that a critical role of *K. pneumoniae*/LPS/HADHA-K353 acetylation signal axis in regulating hepatic metabolism and glucose homeostasis **(Figure 9)**. This study provides a new perspective for understanding the pathogenesis of comorbid MASLD and T2DM, as well as a theoretical basis for developing nutritional intervention strategies that target pathogenic bacteria, their derived metabolites, and PTMs of metabolic enzymes.

**Figure 9.**
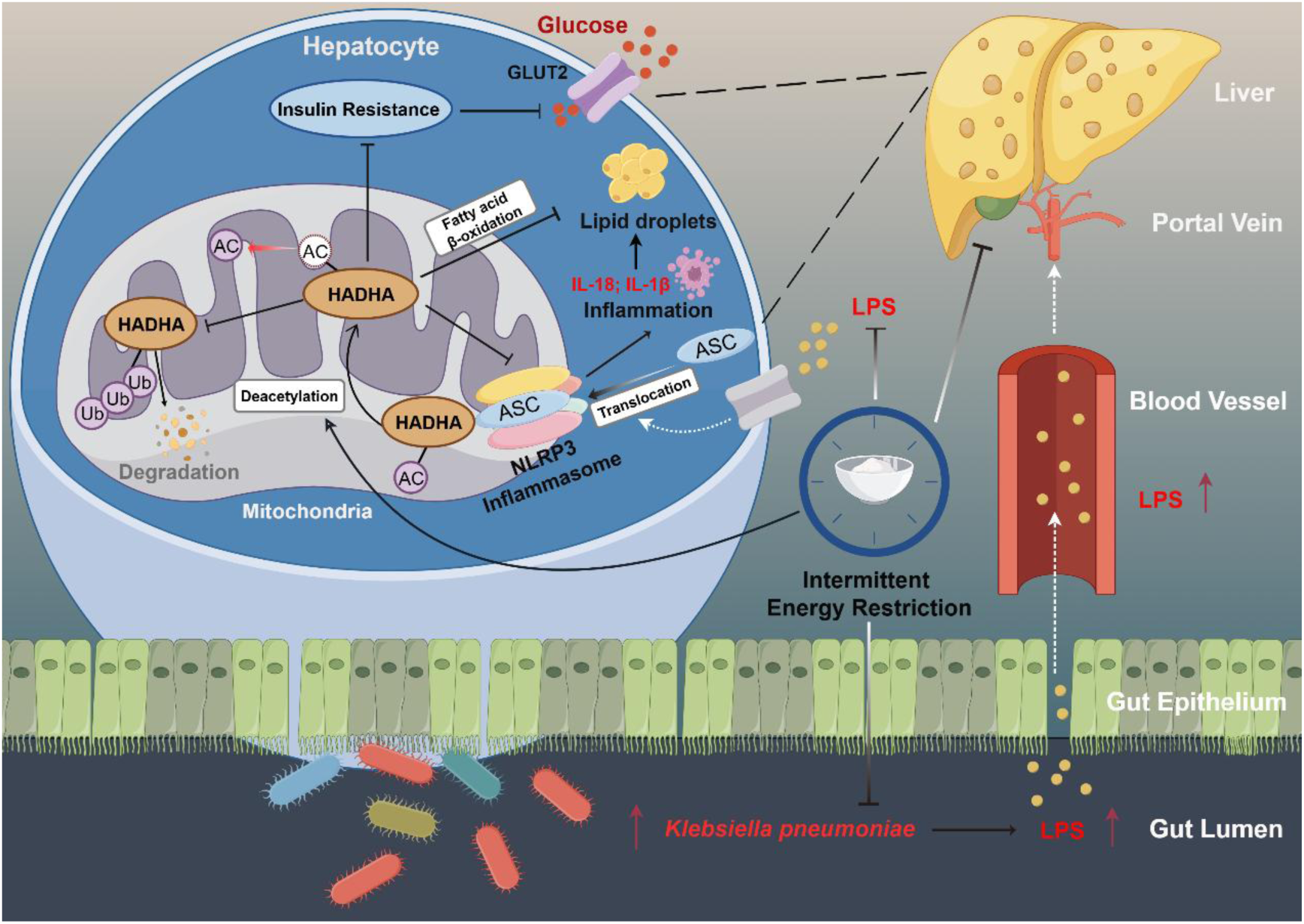
Schematic diagram illustrating the finding of this study. iER improves hyperglycemia, hepatic steatosis, insulin resistance, inflammation and decreases the abundance of gut pathogen *Klebsiella pneumoniae* in comorbid MASLD and T2DM, which is strongly associated with reductions in blood endotoxin, lipopolysaccharide (LPS) levels. *K. pneumoniae* derived LPS exacerbates fat accumulation, insulin resistance, glucose metabolism disorders and inflammation in hepatocytes. The iER promotes hepatic HADHA-K353 deacetylation, which enhances HADHA enzyme activity and protein stability, thereby alleviating hepatic lipid accumulation and insulin resistance. HADHA deacetylation inhibits the binding of HADHA to ASC, blocks the activation of the NLRP3 inflammasome and reduces hepatic inflammation.

### Limitations of the study

Several limitations of this study should be acknowledged. First, while our integrated multi-omics and *in vitro* analyses implicate gut *K. pneumoniae*, its derived LPS, and hepatic HADHA-K353 acetylation as key mechanistic components linking iER to the amelioration of comorbid MASLD and T2DM, a direct causal role for these factors requires further in *vivo* validation (e.g., via gavage of *K. pneumoniae* or its purified LPS). Second, the long-term clinical benefits of iER on disease progression should be assessed in larger, multicentric trials. Therefore, although our data delineate a novel gut-liver axis pathway, its exact causal contribution remains to be fully established.

## Materials and Methods

### Clinical trial design and participants

This randomized controlled clinical trial was conducted as an exploratory pilot study at a single center that comprised part of an ongoing multicenter randomized clinical trial (ClinicalTrials registration no. NCT05439226). Ethics approval was granted by the Biomedical Research Ethics Committee of Hunan Agricultural University (BRECHAU20200235). All participants provided written consent for publication of potentially identifiable clinical information. This study was carried out in accordance with the Declaration of Helsinki. The study protocol detailing participant recruitment, inclusion and exclusion criteria, study conduct, and planned data analyses is published elsewhere[28].

Seventy-six participants were recruited from Hunan Shanshui physical examination center (Hunan, China). Baseline information was collected and assessed, including anthropometrics, a self-reported history of current disease, medical history, dietary habits and exercise habits during the run-in period. Participants from the single center were randomly assigned to an intermittent energy restriction (iER) group or usual care (control) group in a 1:1 ratio determined by a computer-generated random assignment sequence. All participants were blinded. The iER group was instructed to consume the provided low-energy Chinese Medical Nutrition Therapy (CMNT) diet (917 kcal/day) as a specific meal replacement for 5 consecutive days, followed by 10 days of regular food intake for 6 cycles (90 days). Dietary composition of CMNT diet was based on wholegrains and homologous substances of medicine and food. The CMNT diet included four ready-to-consume prepared foods: composite nutritional rice, solid beverages, meal replacement biscuits, and fruit and vegetable gruel. These foods were assigned to three meals a day. As a main component of the CMNT diet, the raw materials of composite nutritional rice include whole grains such as rice, millet, corn, and buckwheat flour, supplemented with the homologous substances of medicine and food such as *Folium Mori* (Mulberry leaf), *Radix Puerariae* (Kudzu root), *Fructus lycii* (Goji berry), *Poria cocos* (Hoelen), and *Ganoderma lucidum* (Reishi mushroom), among others. These multi-component mixtures are manufactured into a low-glycemic index (low-GI) simulated rice grains, which possesses a high content of slowly digestible starch and complex carbohydrates compared with traditional rice. The detailed formulation, macronutrient composition, energy ratios, multivitamin and mineral supplement of composite nutritional rice and other components are provided in **Table S7 and Table S8**. The control group received dietary advice based on the guidelines for the Prevention and Treatment of Type 2 diabetes mellitus in China (2020 edition).

Body weight, medication usage and adverse events were self-reported. The semi-quantitative Food Frequency Questionnaire (FFQ) was used to assess dietary intake. Blood and fecal samples were collected. Anthropometric measures, biochemical measurements (glucose, insulin, CRP, and lipid profile, etc.), liver fat and liver fibrosis assessments were determined at baseline and end of iER intervention at Hunan Shanshui physical examination center (Changsha, China). Serum IL-1β and LPS levels were measured using ELISA kits at the Hunan Provincial Engineering Research Center of Medical Nutrition Intervention Technology for Metabolic Diseases. Detailed information and measurements of the study are provided in **Table S3-Table S6**. The gut microbiota of fecal samples was assessed in before and after iER intervention. Participants in the control group underwent the same tests as those in the iER intervention group, except for the gut microbiota assessment. In addition, glucose-lowering medications dose adjustment was carried out as described in our previous study[28].

Sample size estimation was based on the reduction in HbA1c. The estimated sample size of this trial was 24 per group, allowing a 20% dropout rate[19]. However, due to the COVID-19 pandemic in China, we were only able to include 19-20 subjects per group, which nevertheless proved sufficient to achieve > 80% statistical power. Subsequent analysis indicated the therapeutic potential of iER intervention in this study population.

### Clinical Trial design and Interim Analysis

The interim analysis was not pre-specified in the original study protocol (Protocol-v2.0). It was introduced in Protocol-v3.0 following approval by the Data and Safety Monitoring Board (DSMB) and the Biomedical Research Ethics Committee of Hunan Agricultural University. The supporting details are provided in the Statistical Analysis Plan and Protocol documents. The interim analysis results are presented in the attached Interim report. The DSMB reviewed the interim results and recommended continuation of the trial (see attached DSMB Interim Analysis Meeting Minutes). The purpose of adding an interim analysis is to evaluate early efficacy signals of the iER intervention in patients with comorbid MASLD and T2DM, given unknown risk-benefit profiles in this population. Assessing safety and compliance, particularly for participants with heterogeneous metabolic responses to fixed-energy restriction (917 kcal/day). Additional amendments included broadening BMI eligibility (>18), expanding to six centers to boost recruitment and generalizability, and adding metabolomics/gut microbiota analyses to secondary outcomes for mechanistic exploration. All changes were prospectively included in Statistical Analysis Plan & Protocol. Primary outcomes, study design, and follow-up duration remained unchanged from the original protocol.

### Mice experiments

Twelve-week-old male diabetic db/db mice (BKS.Cg-+ Lepr db /+ Lepr db /Jcl) and age-matched male C57BLKS/JGpt mice (WT) that were used as the healthy controls were purchased from GemPharmatech (Jiangsu, China). All experimental mice were housed in a pathogen-free environment, under a 12 h light/12 h dark cycle at 20-24 °C ambient temperature, with 50% ± 10% relative humidity. All the mice used in this experiment had free access to drinking water throughout the experiment. All animal experiments adhered to the Guidelines for the Care and Use of Laboratory Animals: Eighth Edition (ISBN-10: 0-309-15396-4) and received approval from the Animal Care Committee of Hunan Agricultural University (Permission No. 2023-050).

The db/db mice were randomly assigned to two groups with 6 mice per group. Mice in the db/db_iER group were subjected to the iER regimen, which involved a cyclical dietary pattern. Each cycle consisted of 4 days on a low-calorie CMNT diet that provided 50% of their normal daily energy intake on Day 1 and 10% on Days 2-4, followed by 7 days of *ad libitum* feeding with standard – mouse chow, repeated for a total of 7 cycles. Prior to administering the CMNT diet, mice were transferred to fresh, sterilized cages to avoid coprophagy or the consumption of leftover chow. The mouse CMNT diet was prepared using the same four human meal-replacement components (ingredient list provided in **Table S7**), which were blended together in proportions that precisely matched the human daily intake ratio. This mixture was then homogenized with hydrogel (ClearH_2_O) to create a consistent feed for the mice. The mice of db/db group were fed a regular chow diet without any restrictions. Additionally, age-matched male WT mice served as the normal control group (n=6). Calorie information of mouse CMNT diet and control diet was shown in **Table S9 and Table S10**. Bodyweight, food intake, and water consumption were recorded daily. At the end of the experiment, all mice were killed to collect serum, contents of colon and ileum, and tissues samples including liver, colon and ileum.

### Establishment of an *in vitro* comorbid MASLD and T2DM model

The human immortalized normal liver cell line MIHA was purchased from Hunan Fenghui Biotechnology Co., Ltd (Hunan, China). MIHA cells were cultured in RPMI-1640 medium (Invitrogen) containing 2 g/L glucose, supplemented with 10% fetal bovine serum and 1% penicillin-streptomycin in a humidified incubator at 37°C with 5% CO_2_.

To construct a comorbid MASLD and T2DM model *in vitro*, the MIHA cells were incubated with RPMI-1640 medium containing 40 mM glucose and 0.5 mM free fatty acids (oleic acid: palmitic acid, 2:1, molar ratio, Sigma-Aldrich) (dissolved in 0.5% fatty acid-free BSA) for 48 h, followed by 1 µg/mL LPS for 24 h to mimic HGHF conditions, accompanied by inflammation[72, 73, 74]. To induce NLRP3 inflammasome activation, the MIHA cells were incubated with 5 mM adenosine 5’-triphosphate disodium salt hydrate (ATP) for 30 min before collecting the cell samples[75]. For the control group, cells were stimulated with 0.5% fatty acid-free BSA.

### The isolation and culture of *K*. *pneumoniae*

A fecal sample from a patient with T2DM and MASLD in the iER group, whose relative abundance of *K. pneumoniae* was close to the average at the baseline of the iER group, was selected for the isolation of *K*. *pneumoniae*. Approximately 1 g of fresh stool was suspended in 9 mL of phosphate-buffered saline buffer, vortexed and kept to dissolve in an isothermal shaker maintained at 37 ℃ for 6 h to equilibrate. The mixture was homogenized and centrifuged, and the supernatant was collected. The 10-fold serial dilutions were made in PBS with vigorous shaking between dilutions. MacConkey Inositol Adonitol Carbenicillin agar plates (Huankai Microbial, China) were coated with the different concentrations of fecal bacteria suspension, inverted, and cultured in a 37 °C incubator for 24-48 h. Single colonies were picked from the plate and inoculated into liquid Luria-Bertani (LB) medium. The precipitated bacteria were collected for genomic DNA extraction using the TIANamp Bacteria DNA Kit (DP302; TIANGEN, China). 16S rRNA gene PCR sequencing was performed to confirm identification as a *K*. *pneumoniae* species.

### Whole-genome sequencing and assemblies of *K. pneumoniae*

Genomic DNA was extracted from the isolated *K. pneumoniae* strain (ZH2.1) in a patient with MASLD and T2DM using the Tris-EDTA-NaCl buffer extraction method. The collected DNA was detected using agarose gel electrophoresis and quantified using Qubit. After quality control, the high-quality genomic DNA was subjected to sequencing library building and the genome of *K. pneumoniae* was sequenced using single-molecule, real-time technology. Low-quality reads were filtered using SMRT Link v.8.0 and the filtered reads were assembled using the software Canu to generate one contig without gaps. PacBio long-read sequencing of *K. pneumoniae* (ZH2.1) was performed. The genome-wide phylogeny of the isolated strain ZH2.1 was analyzed using AutoMLST, an automated network tool[76].

### LPS extraction from *K. pneumoniae*

LPS extraction from *K. pneumoniae* was performed using the LPS extraction kit (iNtRON, South Korea) according to the manufacturer’s instructions. *E. coli*, isolated from a donor with T2DM and MASLD in the iER group-derived samples, was used as the control strain. Different types of LPS based on their sources were separated by electrophoresis on 12% polyacrylamide gels and subsequently visualized using silver staining with the Fast Silver Stain Kit (Beyotime, China), following the manufacturer’s instructions. *E.coli* derived LPS served as controls.

### Measurement of LPS and inflammatory factors

Human and mouse serum LPS levels were assayed using Human Lipopolysaccharides (LPS) ELISA Kit (CSB-E09945h, CUSABIO, China) and Mouse Lipopolysaccharides (LPS) ELISA Kit (CSB-E13066m, CUSABIO, China) respectively following manufacturer’s manuals. Human IL-1β ELISA Kit (E-EL-H0149, Elabscience, China) was used to quantify IL-1β concentrations in human serum and cultured MIHA cell supernatants. Human serum IL-18 levels were measured using Human IL-18 ELISA Kit (E-EL-H0149, Elabscience, China) according to manufacturer’s instructions.

### Blood glucose and insulin measurement

Blood glucose levels of mice were measured after 6 h of fasting using a blood glucose test kit (A154-1-1, Nanjing Jiancheng Bioengineering Institute, China) according to the manufacturer’s instructions. Blood samples were collected via tail vein or cardiac puncture. The fasting serum insulin levels were determined using the Mouse Insulin ELISA kit (E-EL-M2614, Elabscience, China). The homeostasis model assessment of insulin resistance (HOMA-IR) was calculated using the following equation: fasting glucose (mmol/L) × fasting insulin (mU/L) /22.5.

### Intraperitoneal insulin tolerance test (ITT)

Mice were single-caged and fasted for 5 h prior to the tests. For the ITT, mice were injected with insulin (0.75 unit/kg, Sigma) intraperitoneally, and blood glucose levels were measured at 0 (baseline), 30, 60, 90, 120,150 and 180 min after injection.

### Liver and serum biochemistry indexes measurements

Serum total triglyceride (TG), total cholesterol (TC), aspartate transaminase (AST) and alanine transaminase (ALT) levels were assayed using an automatic biochemical analyzer (Hitachi, LABOSPECT003, Japan). For liver TG and TC measurements, tissue samples were washed with cold PBS and homogenized at a ratio of 0.2 g tissue to 1.8 mL of 0.9% saline (1:9, w/v) and then were measured with commercial kits according to the manufacturer’s instructions (TG: A110-1-1; TC:A111-1-1, Nanjing Jiancheng Bioengineering Institute). Values were normalized to total protein concentration, as determined by BCA assay (Beyotime, China). All reagents and kits’ source and identifiers were listed in the key resources table.

### HADHA enzyme activity assay

A modified HADHA enzyme activity assay was performed according to the previously described methods[77, 78, 79]. Briefly, cells or liver samples were homogenized by ultrasonication for 10 min with ice cold assay buffer (containing 75 mM Tris (pH 10) with 75 mM KCl, NAD^+^ 1.2 mM) and centrifuged with 8,000 × g for 10 min at 4 °C. The supernatant was collected and the reaction was initiated by adding 3-Hydroxypalmitoyl-CoA (C16) (Chemsoon, China) as the substrate. The reaction mixture contained 100 μL supernatant, 20 μL NAD^+^ (1.2 mM), 10 μL substrate (125 μM), 66 μL assay buffer and 4 μL WST-8 kit (containing WST-8 and 1-mPMS, Beyotime) in each well of a 96-well plate. The initial absorbance at a wavelength 450 nm by microplate reader was recorded as the baseline. After 30-min incubation at 37 °C, the developing color reaction was recorded as the final reading. HADHA enzyme activity is expressed as the difference in absorbance values between the two readings and is corrected for protein concentration to give enzyme activity in U per microgram of protein. One unit of enzyme activity is defined as a decrease of 1 nmol NADH per min for 1 mg of protein. The results are expressed as a percentage of the control group.

### qRT-PCR

Total RNA was isolated from liver tissue or MIHA cells using Total RNA extraction kit (DP419, Tiangen, China) according to the manufacturer’s instructions. The concentration and purity of total RNA were determined by spectrophotometry, whereas the integrity of the RNA was confirmed by electrophoresis. 50 ng of total RNA was reverse-transcribed using cDNA Synthesis SuperMix (TSK302M,TSINGKE). Real-time PCR was carried out in a CFX96TM real-time system (Bio-Rad) with 2×T5 Fast qPCR Mix (TSE202, TSINGKE) and gene-specific primers. The following conditions were used for real-time PCR: 95 °C for 1 min, then 95 °C for 10 s, 58°C for 5 s and 72 °C for 15 s in 40 cycles. The 2^−ΔΔCT^ method was used to analyze the relative changes in gene expressions normalized against β-actin mRNA expression. The primers are described in **Table S11**.

### Western blotting and Immunoprecipitation

The protein of liver tissue or cultured MIHA cells was extracted, as whole-cell lysates, using RIPA lysis buffer (P0013B, Beyotime, China) with PMSF. For separate cell membrane protein extraction, we used membrane and cytosol protein extraction kit (P0033, Beyotime, China). For analyzing proteins in isolated mitochondria, mitochondria were extracted from fresh mouse liver or MIHA cells using Tissue Mitochondrial Isolation Kit (C3606, Beyotime, China) or Cell Mitochondrial Isolation Kit (C3601, Beyotime, China), respectively, as instructed. Total protein was measured with BCA kit (P0010S, Beyotime). Then, equal quantities of the indicated protein were separated by 10-15% SDS polyacrylamide gel electrophoresis (SDS-PAGE) and transferred to nitrocellulose membranes (66485, Pall, USA). The membranes with proteins were blocked with 5% nonfat milk in Tris-buffered saline Tween-20 (TBST), incubated with the indicated primary antibodies overnight at 4°C and incubated with HRP-conjugated secondary antibodies. The proteins were then detected using an Enhanced chemiluminescence (ECL) substrate solution (WBKLS0500, Millipore) and visualized in a iBright FL1500 Imaging System (Thermo Fisher Scientific, USA). β-Actin was used as loading control for the whole cell lysate. Na^+^/K^+^ ATPase was used as a loading control for membrane fraction. HSP60 served as loading control for mitochondrial fractions. Densitometry-based quantification of western blots was performed with ImageJ software. For Co-IP assays, liver tissue or cultured MIHA cells were lysed using IP lysis buffer (P0013, Beyotime, China). The supernatants were incubated with either anti-HADHA or anti-Flag antibodies at 4 °C overnight with gentle agitation. Mouse and rabbit control IgGs were used as negative control. Antibody-linked complexes were captured by incubation with Dynabeads™ Protein G beads (Thermo Fisher Sci., 10007D) for 3 h at 4 °C. Protein bound to magnetic beads were mixed with loading buffer and boiled at 100 °C for 5 min and subjected to Western blotting. The IPKine HRP, Goat anti-rabbit IgG HCS (A25222, Abbkine) was applied as appropriate to avoid the interference of the antibody light chain. Antibodies used are listed in **Table S1**.

### Cytotoxicity assay

The cytotoxicity of LPS on MIHA cells was measured using a Cell Counting Kit-8 (CCK-8) assay kit (Beyotime, China). MIHA cells were seeded into 96-well plates and treated with different concentrations of LPS for 24 h. 10 μL CCK-8 solution was added to each well and incubated for 2 h in the incubator at 37 °C with 5% CO_2_. Cell viability was determined by measuring the absorbance at 450 nm, using a microplate reader (Bio-TekELx800, USA).

### Transmission electron microscopy (TEM) and Gram staining

The mitochondrial and lipid droplets morphology of mouse liver was examined by TEM assay. Mouse liver tissues were fixed, embedded and ultrathin sectioned, dehydrated in a gradient ethanol series and embedded in epoxy resin. Samples were then stained with 4% uranyl acetate and counterstained with lead citrate. Images were photographed with a Hitachi HT7800 transmission electron microscope (Hitachi, Japan). For TEM analyses of bacteria, *K. pneumoniae* was grown at 37℃ in LB broth. Cells were harvested by centrifugation at 10, 000 rpm for 10 min at 4°C and then fixed with 2.5% glutaraldehyde solution in PBS, embedded with conventional methods, and slices were observed with the Hitachi HT7800 transmission electron microscope (Hitachi, Japan). Gram staining was performed using a Gram Stain Kit (G1060, Solarbio, China) following the manufacturer’s protocol. The *K. pneumoniae* was imaged using a bright field microscope (Leica, Germany).

### Histopathological analysis

The livers or intestines (colon and ileum) of mice were fixed in 10% neutralized formaldehyde, embedded in paraffin, and sectioned. Paraffin sections were stained with hematoxylin and eosin (H&E) for morphology. Liver fibrosis was evaluated by Masson’s trichrome staining. Oil Red O (Sigma, USA) staining was performed on liver cryosections embedded in OCT to evaluate lipid droplets accumulation. All images shown are representative results of at least three biological replicates. HE staining analyzes the pathological changes of the liver and intestines. The pathological grading of liver samples was evaluated using the NAFLD activity score (NAS) according to a previous report[80]. Intestinal injury scoring was evaluated using a Chiu’s score method[81]. For quantification of Oil Red O areas and Masson’s trichrome staining, the images were subsequently analyzed using ImageJ.

### Oil Red O, Bodipy 493/503 and Nile Red staining for Cells

For cell culture staining, cells were fixed with 4% paraformaldehyde for 20 min at room temperature, washed with PBS, and then subjected to Oil Red O (O0625, Sigma) and Bodipy 493/503 (D3922, Thermo Fisher) and Nile Red (C2051S, Beyotime, China) staining according to the standard protocols. The image was performed using Leica microscope (Leica, Germany).

### Immunofluorescence for cells

The translocation of GLUT2 to the plasma membrane in MIHA cells was observed by immunofluorescence staining. Briefly, MIHA cells were plated in cell culture dishes and fixed with 4% paraformaldehyde for 20 min, followed by permeabilization using 0.1% saponin. Subsequently, MIHA cells were blocked with 10% normal goat serum and were incubated with anti-GLUT2 antibody (1:200) at 37 °C for 1 h, and then incubated with Alexa Fluor 488-conjugated Donkey anti-Goat IgG (H+L) for 1 h at 37 °C. Nuclei were stained with DAPI. Fluorescence images were captured by using a Leica DM 4000 fluorescence microscope with a 100× oil immersion objective.

### HADHA siRNA knockdown in hepatocytes

For HADHA knockdown, MIHA cells were transfected with siRNA (GenePharma, Shanghai, China) using the Lipo8000 transfection reagent following the manufacturer’s instructions (Beyotime Biotechnology, China). Negative siRNA was used as a normal control. Western blot analysis was used to assess the knockdown efficiency of HADHA-targeting siRNA 72 h after transfection. The siRNA sequences are listed in the **Table S11**.

### Stable overexpression of HADHA in MIHA cell line

Blank pCDH-CMV-MCS-EF1-GFP+Puro vector was purchased from Fenghui Biotechnology (Changsha, China). For stable overexpression of HADHA, the full-length CDS of human HADHA was cloned into the pCDH-CMV-MCS-EF1-GFP+Puro vector to generate Flag-HADHA plasmid (**Figure S11A**). To establish an MIHA cell line stably overexpression of HADHA, lentivirus was produced by co-transfecting HKT293T cells with pCDH-HADHA plasmid and packaging plasmids psPAX2 and pMD2.G (Fenghui Biological, China). Viral supernatants were harvested after 48 h of transfection and then used to infect MIHA cells. Cell lines were screened in 1640 culture medium containing 2.5 μg/mL puromycin (Beyotime, China). Cells resistant to puromycin were isolated and cultured for subsequent treatments.

### Mutagenesis of HADHA

For site-directed mutagenesis, the HADHA^WT^ plasmid expressing human KR mutant (converting lysine site of HADHA to arginine) at Lysine 353 of human HADHA (K353R) was constructed using Mut Express II Fast Mutagenesis Kit V2 (Vazyme, China). The point mutation was generated by overlap extension PCR by segmented amplification with primers containing mutations shown in **Table S11**. CE Design V1.04 (https://crm.vazyme.com/cetool/multipoint.html) was used for primer design. All the mutations introduced were verified by DNA sequencing. For stable overexpression of HADHA-K353R in MIHA cell line, the plasmids were treated similarly to HADHA^WT^ plasmid.

### Molecular docking analysis

Molecular docking analysis was performed to determine HADHA and ASC interactions. The three-dimensional structures of wild-type human HADHA (PDB ID: 6DV2) and full-length human ASC (PDB ID: 2KN6) were downloaded from the Protein Data Bank (PDB). The three-dimensional structure of HADHA-K353R was predicted usingAlphaFold2. Docking between HADHA (wild-type and its mutant) and ASC was performed by using the online platform HDOCK server (http://hdock.phys.hust.edu.cn/). All 100 possible docking models were analyzed to identify potential interaction surfaces between HADHA (wild-type and its mutant) and ASC. The biomolecular visualization tool PyMOL was employed to highlight the interaction sites between wild-type HADHA, its mutant K353R, and ASC. Differences in binding between wild-type HADHA, its mutants, and ASC were compared, and structural images of wild-type HADHA and its mutant bound to ASC were generated.

### 16S rRNA Microbiome Sequencing

Fecal contents from the ileum and colon of mice were collected. The total DNA was extracted using TIANamp Stool DNA Kit (DP328, TIANGEN, China) according to the manufacturer’s instruction. The bacterial hypervariable V3-V4 region of 16S rRNA genes were amplified using the universal primer 341F/805R (**Table S11**), and the products were subjected to sequencing on Illumina Novaseq 6000 PE250 platform (LC-Bio Technology Co., Ltd., China). 16S rRNA gene raw sequence reads were processed, trimmed, and clustered into amplicon sequence variants (ASVs) using DADA2. The sequence alignment of species annotation was performed by QIIME2 plugin feature-classifier, and the alignment database was SILVA and NT-16S[82]. Alpha and beta diversities were calculated using QIIME2, Relative abundance was used ins bacterial taxonomy. The one-way ANOVA with Tukey’s multiple comparison test was used to identify the differentially abundant genus, and significances were declared at p < 0.05. LDA effect size (LEfSe, LDA ≥ 3.0, p < 0.05) was performed using nsegata-LEfSe.

### Metagenomics sequencing

Fecal samples were collected before and after the iER interventions and the total DNA was extracted using Fecal Genomic DNA Extraction Kit (DP712, TIANGEN, China). Shotgun metagenomic sequencing of fecal DNA was performed on an Illumina NovaSeq 6000 platform with PE150 (LC-Bio Technology Co., Ltd., China). Sequencing reads were quality-processed and aligned to the human genome hg19 as described in detail previously[83]. The Wilcox test was used to identify the differentially abundant species, and significances were declared at p < 0.05 and |log2-fold change| > 1. Microbial functions were assigned using the gene ontology (GO) term enrichment analysis. Spearman’s correlation analysis was performed using R package, and only correlations with p < 0.05 are displayed.

### Transcriptome analysis

Total RNA was extracted from mouse liver tissues following the instructions of RNeasy Micro Kit (74004, QIAGEN). Sequencing libraries were prepared using NEBNext Ultra RNA Library Prep Kit for Illumina (E7530, NEB). Illumina HiSeq 2500 systems were used for RNA sequencing. Fragments per kilobase of exon model per million mapped reads (FKPM) of every mRNA were used for further analysis. Genes with |log_2_FoldChange| > 0.5849 and p < 0.01 were considered to be significantly differentially expressed genes (DEGs). KEGG pathway enrichment analysis of DEGs related to hepatic lipid metabolism were performed using the OmicShare tools (https://www.omicshare.com/tools).

### Proteomics analysis

The proteins in liver samples from the db/db mice with or without iER intervention were extracted using Urea (8M Urea, 100mM Tris/HCl, pH 8.5) buffer. Protein digestion was conducted using trypsin at a ratio of 1:50 (trypsin/protein). The enzymatically digested peptides were labeled according to the Tandem mass tag (TMT) 10plex reagent kit instructions. The peptides were fractionated using an UltiMate™ 3000 Rapid Separation LC system (Thermo Scientific, USA) and were ionized by a NanoESI source before being subjected to a tandem mass spectrometer (Orbitrap Exploris™ 480, Thermo Fisher Scientific, USA) for data-dependent acquisition (DDA) mode detection. The resulting MS/MS data were processed using MaxQuant search engine (version 2.1.4.0). Tandem mass spectra were searched against the *Mus musculus* SwissProt database concatenated with reverse decoy database. Proteins with |log2FoldChange| > 0.263 and p < 0.05 were considered to be significantly differentially expressed proteins (DEPs).

### Acetyl-proteomics analysis

Liver samples were homogenized in tissue lysis buffer, then subjected to sonication and centrifugation. To enrich acetylated peptides, proteins were digested with trypsin and then dissolved in Immunoaffinity Purification (IAP) Buffer, added pretreated Anti-Ac-K antibody beads (PTMScan Acetyl-Lysine Motif (Ac-K) Kit, Cell Signaling Technology), then incubated for 1.5 h at 4℃ with rotation. The peptides bound to the beads were eluted with 0.1% trifluoroacetic acid. The eluted peptides were then desalted using C18 ZipTips (Millipore, Burlington, MA, USA) and analyzed by liquid chromatography-tandem mass spectrometry (LC-MS/MS). For protein identification and quantification, MS raw data were processed using MaxQuant software. Tandem mass spectra were searched against the *Mus musculus* SwissProt database concatenated with reverse decoy database. Protein quantitative normalization is employed to remove the effect of protein expression levels on modification abundance, enabling its application in subsequent bioinformatics analyses. Differentially Acetylated sites were identified with a fold-change cutoff of ≥2 and p < 0.05.

### Statistical analysis

GraphPad Prism version 10 (GraphPad Software) was used for statistical analysis. The experimental data were shown as the mean ± SEM values (normal distribution) or median (interquartile range (IQR): 25%-75%, non-normal distribution). The Shapiro-Wilk test was applied to assess whether the data followed a normal distribution. Based on the normality test results, parametric tests were used for data with normal distribution, nonparametric tests were applied otherwise. For paired testing, parametric paired t-tests were used for data with normal distribution, and the Wilcoxon signed-rank test was used for data with non-normal distribution. For unpaired testing, parametric t-tests (Student’s unpaired t-test) and nonparametric Mann-Whitney U tests were used for comparisons between two groups. For multiple group comparisons, one-way ANOVA with Tukey post-hoc test was used for data with normal distribution, and the Kruskal-Wallis test was used for data with non-normal distribution in similar scenarios. All tests were done two-tailed. Differences between proportions were tested using the χ^2^ test. Furthermore, to account for potential baseline imbalances, between-group differences in post-intervention outcomes were analyzed using Analysis of Covariance (ANCOVA). For correlation analyses, Spearman’s correlations were used for non-normally distributed data and Pearson’s correlations were used for normally distributed data. A p-value < 0.05 was considered statistically significant for all tests. ns, not significant. *p < 0.05, **p < 0.01, ***p < 0.001.

### Data availability

All raw sequence data generated in this study have been deposited in the National Genomics Data Center, China National Center for Bioinformation, under project accession code PRJCA041614. The metagenomic sequencing data generated have been deposited in the Genome Sequence Archive (GSA) section of the National Genomics Data Center under accession code CRA027030. The sequence data from 16S rRNA sequencing experiments have been deposited in GSA section under accession code CRA026967 for colon contents of db/db mice and CRA026968 for ileum contents of db/db mice. The genome sequencing of *Klebsiella pneumoniae*_ZH2.1 have been deposited in GSA section under accession code CRA027395. The transcriptome raw data generated have been deposited in GSA section with accession number CRA027232. The raw proteome and acetylated proteome data generated have been deposited in the OMIX section of the National Genomics Data Center under accession codes OMIX010782 and OMIX010776, respectively.

## Acknowledgements

This research was financially supported by the National Nature Sciences Foundation of China (82401036); the Changsha Science and Technology Project (kh1801123); the Natural Science Foundation of Hunan Province, China (2025JJ60788); Hunan Science and Technology Innovation Plan (2025ZYJ003).We would like to thank the staff members of Hunan Shanshui Physical Examination Center for performing participants health examinations in this study.

## Author Contributions

D.L., and W.L. contributed to the study design. W.L., Z.P., K.T., X.O., Z.L. and X.C. performed animal studies and cellular studies. R.W., W.L. and Z.Y. recruited participants and collected clinical samples. D.Z. supported the study as an endocrinologist. W.L., K.T. and H.L. performed bioinformatics and statistical analyses. W.L. wrote the manuscript. D.L., X.L. and Z.X. reviewed and edited the manuscript. J.L. and X.L. coordinated and supervised the study. All authors discussed the results, commented on the manuscript and approved the final manuscript.

