## Supplementary Materials for "An intermittent energy restriction diet ameliorates comorbid MASLD and T2DM through the *Klebsiella pneumoniae*/LPS/Hepatic HADHA-K353 acetylation axis"

**Figure S1.** A flow diagram of an exploratory intervention trial in patients with MASLD and T2DM, related to Figure 1.

**Figure S2.** iER exhibits robust metabolic protective effects on in patients with MASLD and T2DM in a pilot randomized controlled clinical trial, related to Figure 1.

**Figure S3.** Comparison of gut microbial composition before and after iER intervention in patients with MASLD and T2DM based on shotgun metagenomic sequencing, related to Figure 2.

**Figure S4.** Genome sequence characteristics and functional annotation of *K. pneumoniae* ZH2.1 strain, related to Figure 3.

**Figure S5.** Metabolic studies in db/db mice, related to Figure 4.

**Figure S6.** Alteration of intestinal barrier function of mouse ileum tissue and gut microbiome composition of mouse ileum contents in response to iER intervention, related to Figure 5.

**Figure S7.** Transcriptome sequencing analysis reveals alterations in hepatic lipid metabolism by iER intervention in db/db mice, related to Figure 6.

**Figure S8.** Liver proteome further reveals iER intervention increases lipid metabolism-related differential proteins in db/db mice, related to Figure 6.

**Figure S9.** Liver protein acetylome reflected alteration of fatty acid metabolism by iER intervention in db/db mice, related to Figure 6.

**Figure S10.** Effects of LPS combine with HGHF treatment on HADHA expression, pan-acetylated/ubiquitination modifications, insulin resistance and inflammation in MIHA cells, related to Figure 7.

**Figure S11.** The effect of HADHA overexpression and knockdown HADHA on lipid deposition, insulin resistance and inflammation in HGHF combine with LPS MIHA cells, related to Figure 7.

**Figure S12.** The effect of HADHA-K353R on lipid deposition, insulin resistance and inflammation in HGHF combine with LPS MIHA cells, related to Figure 7 and Figure 8.

**Table S1.** Key resources in current study.

**Table S2.** Baseline characteristics of the randomized participants, related to Figure 1.

**Table S3.** Changes in clinical characteristics from pre- to post-control or iER among completed participants, related to Figure 1.

**Table S4.** Comparison of post-intervention outcomes between Control and iER groups using ANCOVA, related to Figure 1.

**Table S5.** Dietary information between the iER group and the control group, related to Figure 1.

**Table S6.** Glucose-lowering medication changes, related to Figure 1.

**Table S7.** Diet ingredients and daily intake information of human Chinese medical nutrition therapy (CMNT) Diet, related to Figure 1.

**Table S8.** Calorie information of CMNT diet, related to Figure 1.

**Table S9.** Mouse CMNT diet calorie information, related to Figure 3.

**Table S10.** Mouse regular chow diet calorie information, related to Figure 3.

**Table S11.** Primer sequences for qRT-PCR or PCR, related to Figure 5-Figure 7.

#### Supplementary Figures

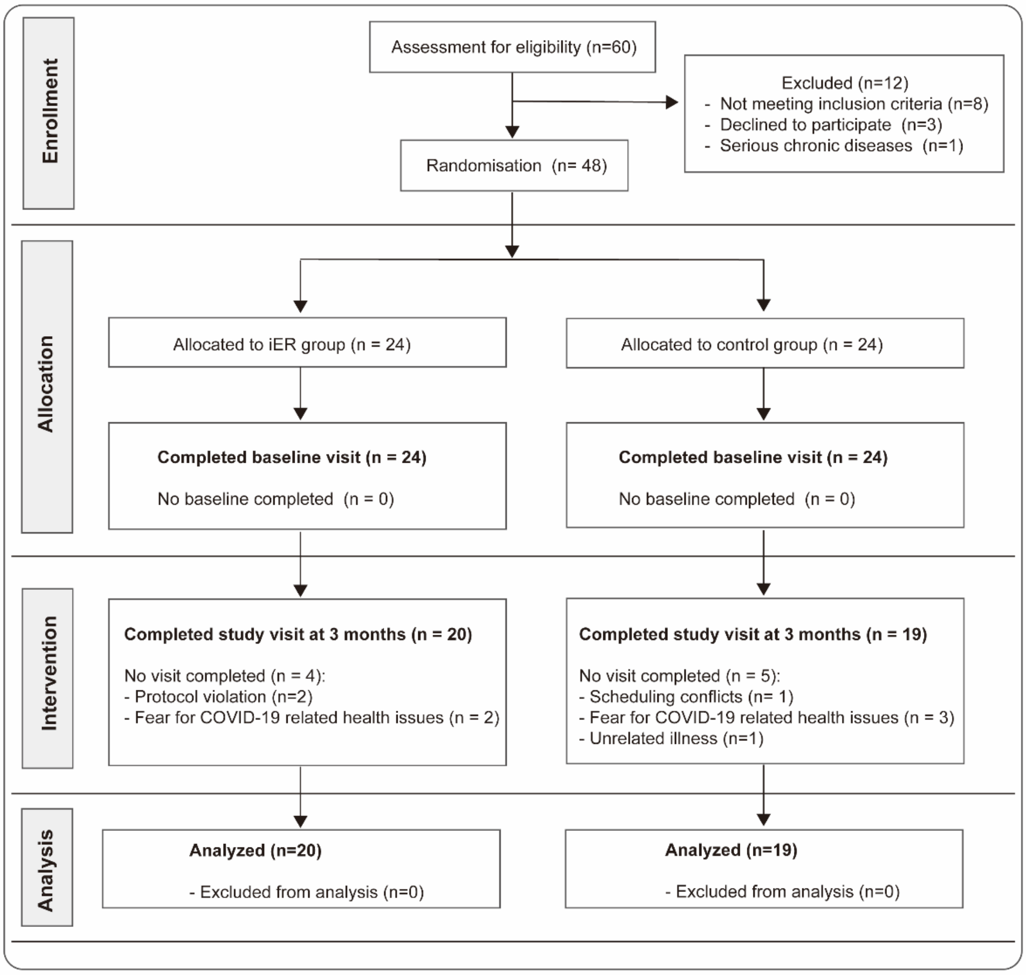


**Figure S1. A flow diagram of an exploratory intervention trial in patients with MASLD and T2DM, related to Figure 1.** A total of 60 individuals were screened and 12 individuals were excluded as they did not meet one or more inclusion criteria. 48 participants were randomized into iER group (n=24) and control group (usual care, n=24). Baseline measures were assessed after randomization. At the end of the intervention, there were 20 individuals in iER group and 19 individuals in the control group.

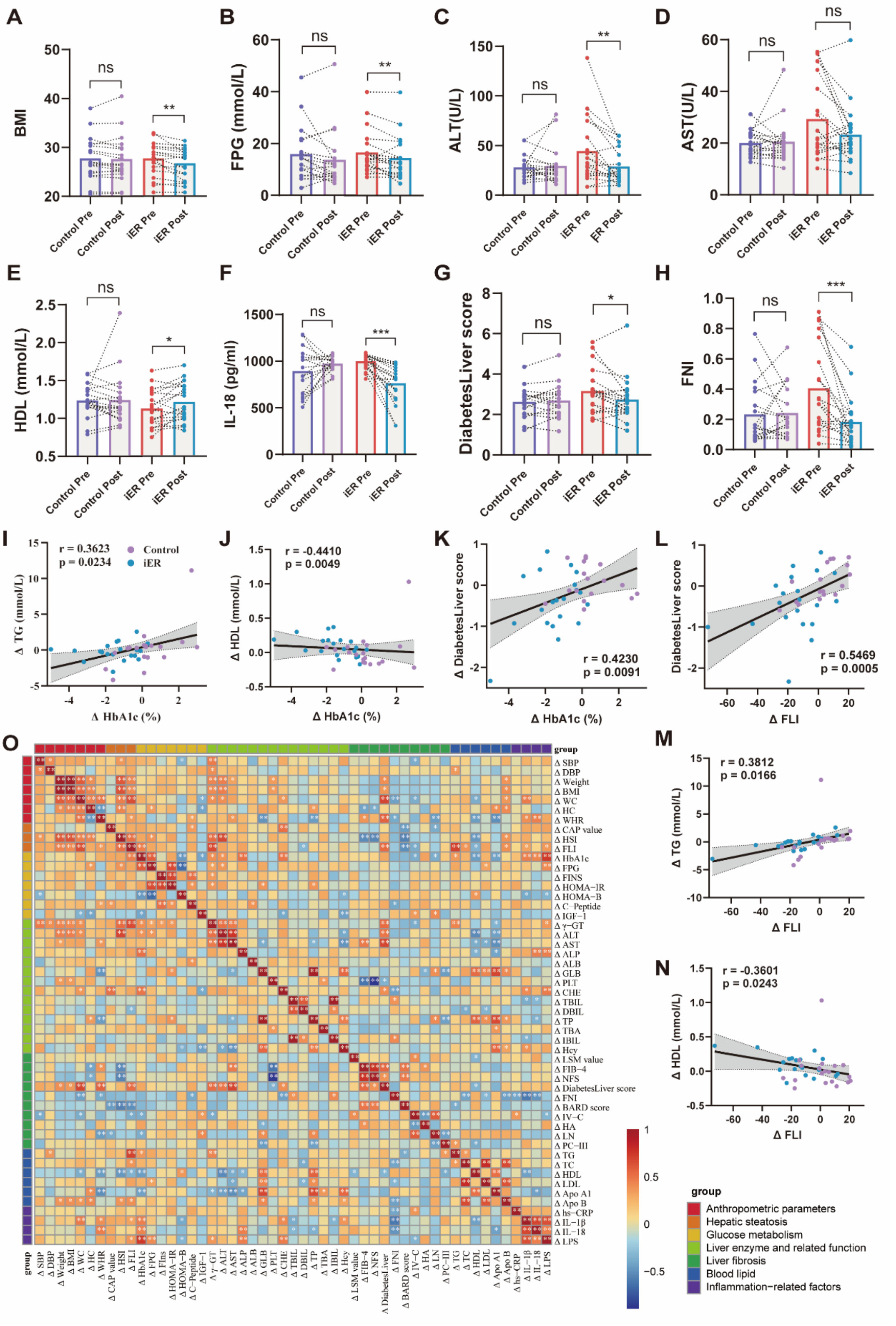


**Figure S2. iER exhibits robust metabolic protective effects on in patients with MASLD and T2DM in a pilot randomized controlled clinical trial, related to Figure 1. (A-H)** Changes in BMI (A), FPG (B), ALT (C), AST (D), HDL (E), IL-18 (F), DiabetesLiver score (G) and FNI (H) levels at baseline (Pre) and at the end of the study (Post) in both the iER group and the control group. The number of samples for each clinical characteristic is shown in **Table S3**. Bars show mean, thin lines show paired data. p-values were calculated using two-tailed Wilcoxon signed-rank tests for non-normally distributed paired data and paired t tests for normally distributed data. *$p < 0.05$, **$p < 0.01$, ***$p < 0.001$, ns, no significance. **(I-K)** Scatter plots of the correlations between changes of HbA1c and changes of serum TG (I), serum HDL (J), and DiabetesLiver score, respectively. **(L-N)** Correlations between changes of FLI and changes of DiabetesLiver score (L), serum TG (M), and serum HDL (N), respectively. **(O)** The heatmap of Spearman's correlation coefficients between different clinical characteristics. Red indicates positive correlations and blue indicates negative. The correlation analysis were performed in all completed participants ($n = 39$). Spearman's correlations were used for non-normally distributed data and Pearson's correlations were used for normally distributed data. r, correlation coefficient, and $p < 0.05$ was considered statistically significant. Source Data are provided as a Source Data file.

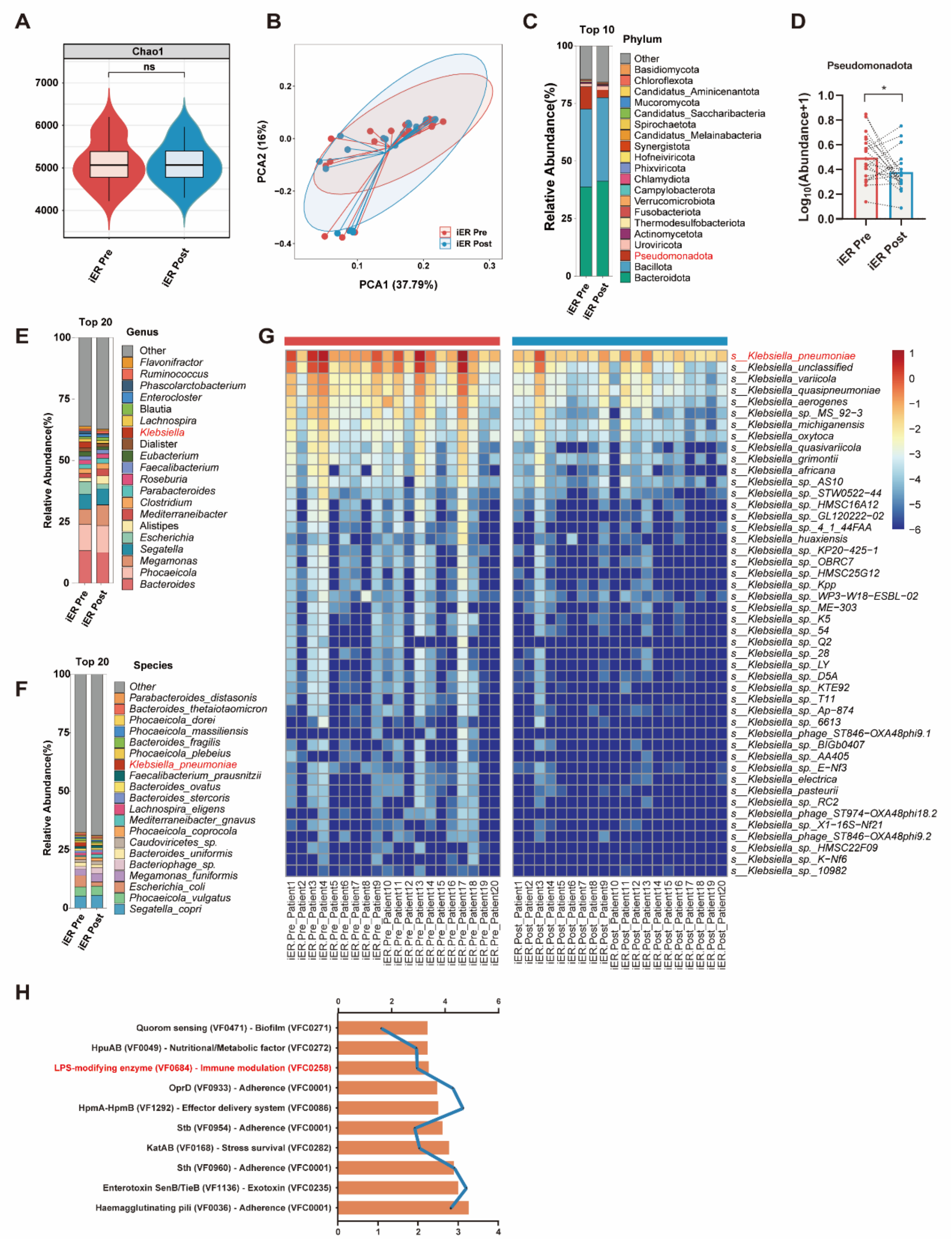


**Figure S3. Comparison of gut microbial composition before and after iER intervention in patients with MASLD and T2DM based on shotgun metagenomic sequencing, related to Figure 2. (A)** Changes in alpha diversity were assessed by Chao index. **(B)** Samples were clustered using Principal Coordinates Analyses (PCA) based on Bray-Curtis dissimilarities. **(C-F)** Bacterial taxonomic profiling at the phylum, genus and species level, respectively and the relative abundance of phylum Pseudomonadota in baseline and the end of iER intervention. **(G)** Changes in the relative abundance of *Klebsiella* at the species level were analyzed among all differential species before and after the iER intervention. **(H)** Virulence-associated genes involved with LPS modifying enzyme (VF0684) were identified by screening genome sequences against the VFDB 2023. Bars show mean, thin lines show paired data. The p-values were calculated using two-tailed Wilcoxon signed-rank tests for paired comparisons. $*p < 0.05$, $**p < 0.01$, $***p < 0.001$, ns, no significance. Source Data are provided as a Source Data file.

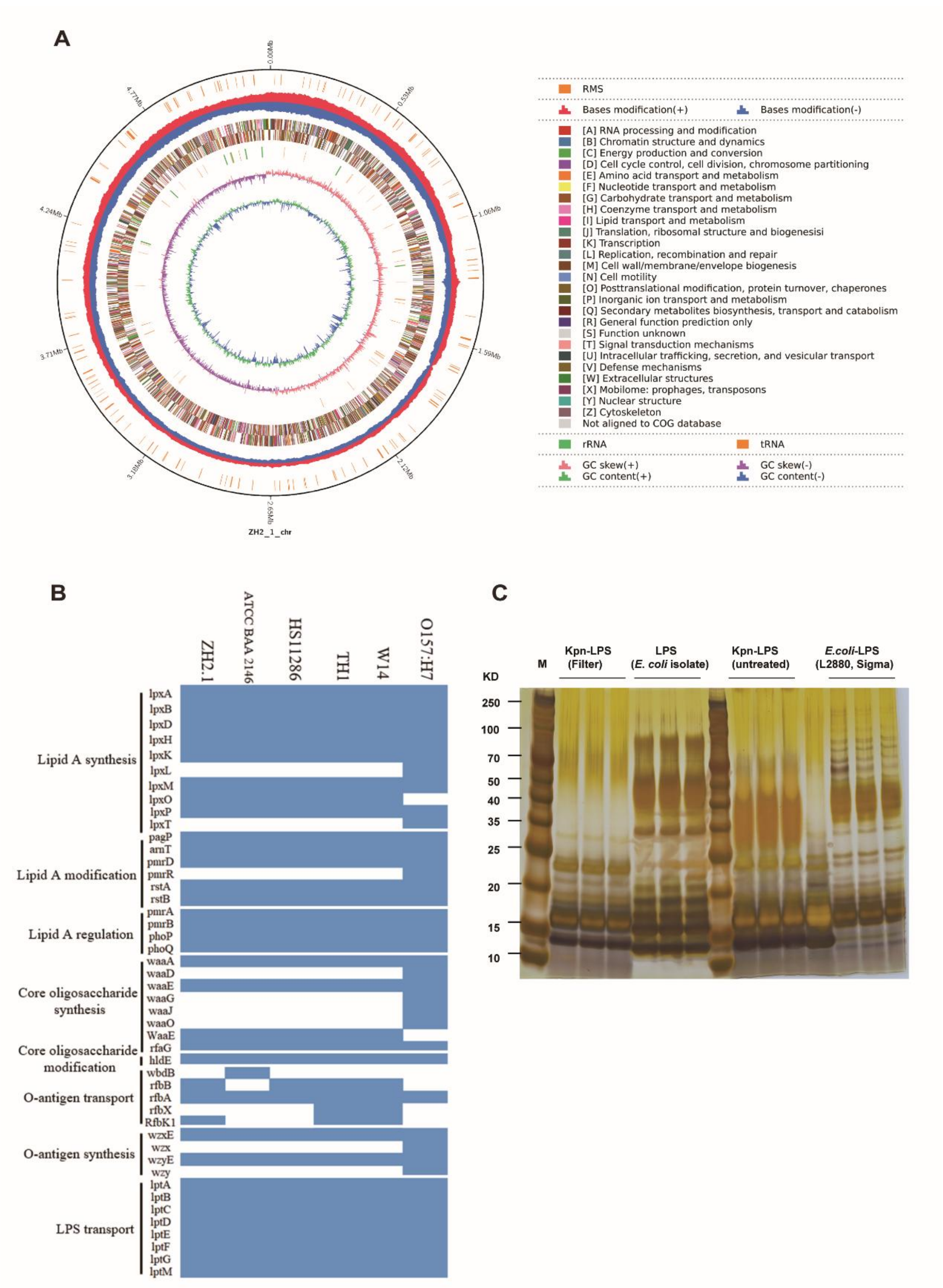


**Figure S4. Genome sequence characteristics and functional annotation of *K. pneumoniae* ZH2.1 strain, related to Figure 3. (A)** Circular genomic map of the *K. pneumoniae* clinical strain ZH2.1 chromosome. The circles from outside to inside represent the distribution of the Cluster of Orthologous Groups (COG) functional categories, genome location coordinates, restriction modification enzymes, base modification sites located in the positive chain, base modification sites located in the negative chain, coding genes in the positive strand, coding genes in the negative strand, rRNA and tRNA distribution, genomic GC skew value, and genomic GC content. **(B)** Sequence comparison among six strains (*K. pneumoniae* ATCC BAA-2146, HS11286, TH1, W14 and *E. coli* O157:H7) based on the 46 genes related to lipopolysaccharide (LPS) synthesis and modification. The blue blocks represent the presence of the genes listed on the left, while blank blocks indicate their absence of the related genes. **(C)** Silver-stained Tricine-SDS-PAGE gel depicting the LPS profiles from strains *K. pneumoniae* and *E. coli*.

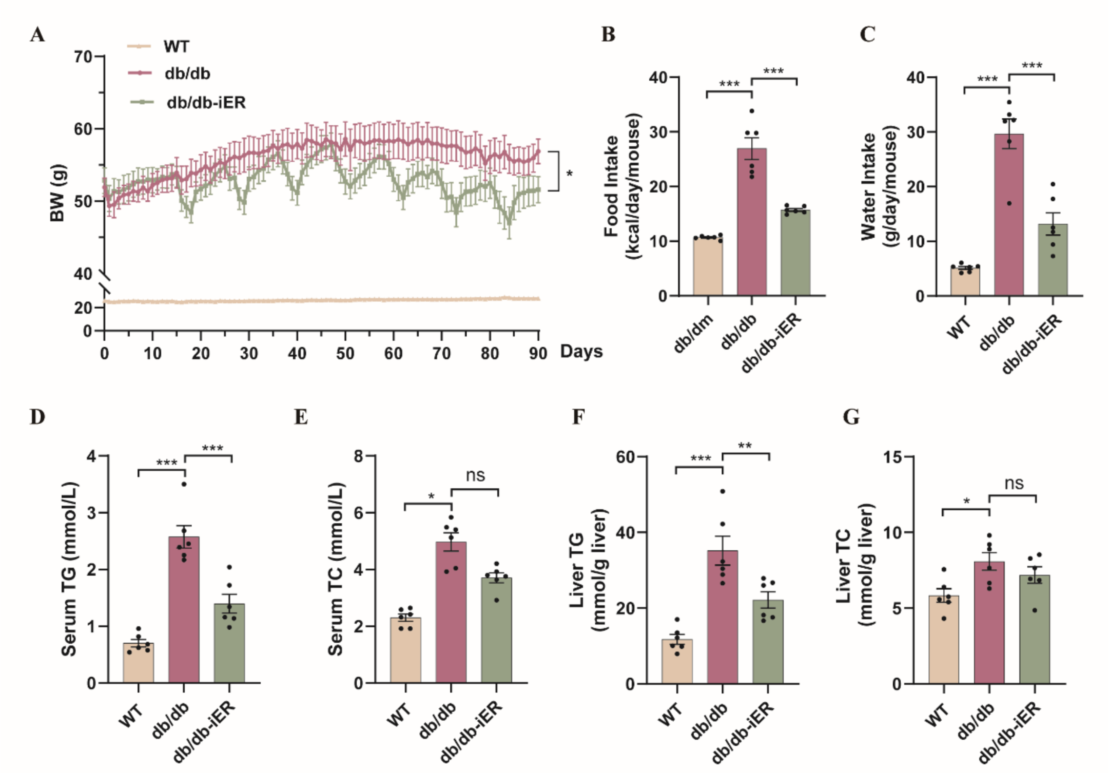


**Figure S5. Metabolic studies in db/db mice, related to Figure 4. (A)** Body weight. **(B)** Food intake. **(C)** Water intake. **(D)** Serum TG. **(E)** Serum TC. **(F)** Liver TG. **(G)** Liver TC. Data are presented as the mean ± SEM. Statistical significance was determined by one-way ANOVA with Tukey tests for multiple comparison test. *$p < 0.05$, **$p < 0.01$, ***$p < 0.001$. Source Data are provided as a Source Data file.

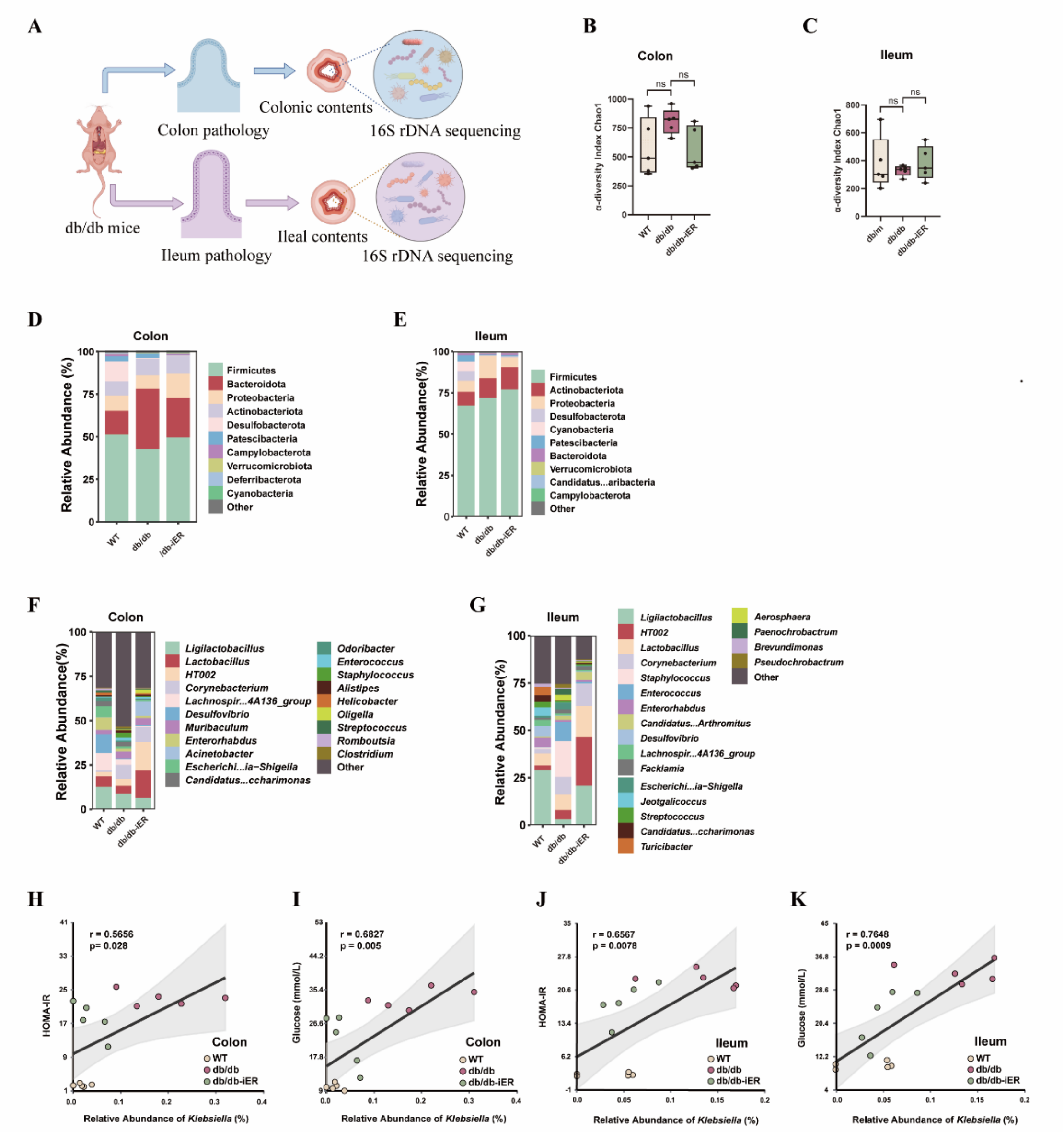


**Figure S6. Alteration of gut microbiome composition in the colon and ileum of mice in response to iER intervention, related to Figure 5. (A)** The experimental workflow of showing colon and ileum sections pathology and bacterial 16S rDNA microbiome analysis from colon and ileum contents in db/db mice. **(B-C)** Alpha diversity of the colon and ileum contents of mice was assessed by Chao1 based on 16S rDNA sequencing. **(D-E)** Bacterial taxonomic profiling at the phylum level (top 10) in colon and ileum contents. **(F-G)** Bacterial taxonomic profiling at the genus level (top 20) in colon and ileum contents. **(H-K)** Spearman's correlations between the relative abundance of the genus Klebsiella in the colon contents and HOMA-IR or fasting blood glucose, and between that in the ileum contents and HOMA-IR or fasting blood glucose, in mice. r, Spearman's correlation coefficient, and $p < 0.05$ was considered statistically significant. Data are shown as mean ± SEM. The p-values were calculated using Kruskal-Wallis test with post hoc Dunn's multiple comparison test (B-C). *$p < 0.05$, **$p < 0.01$, ***$p < 0.001$. Source Data are provided as a Source Data file.

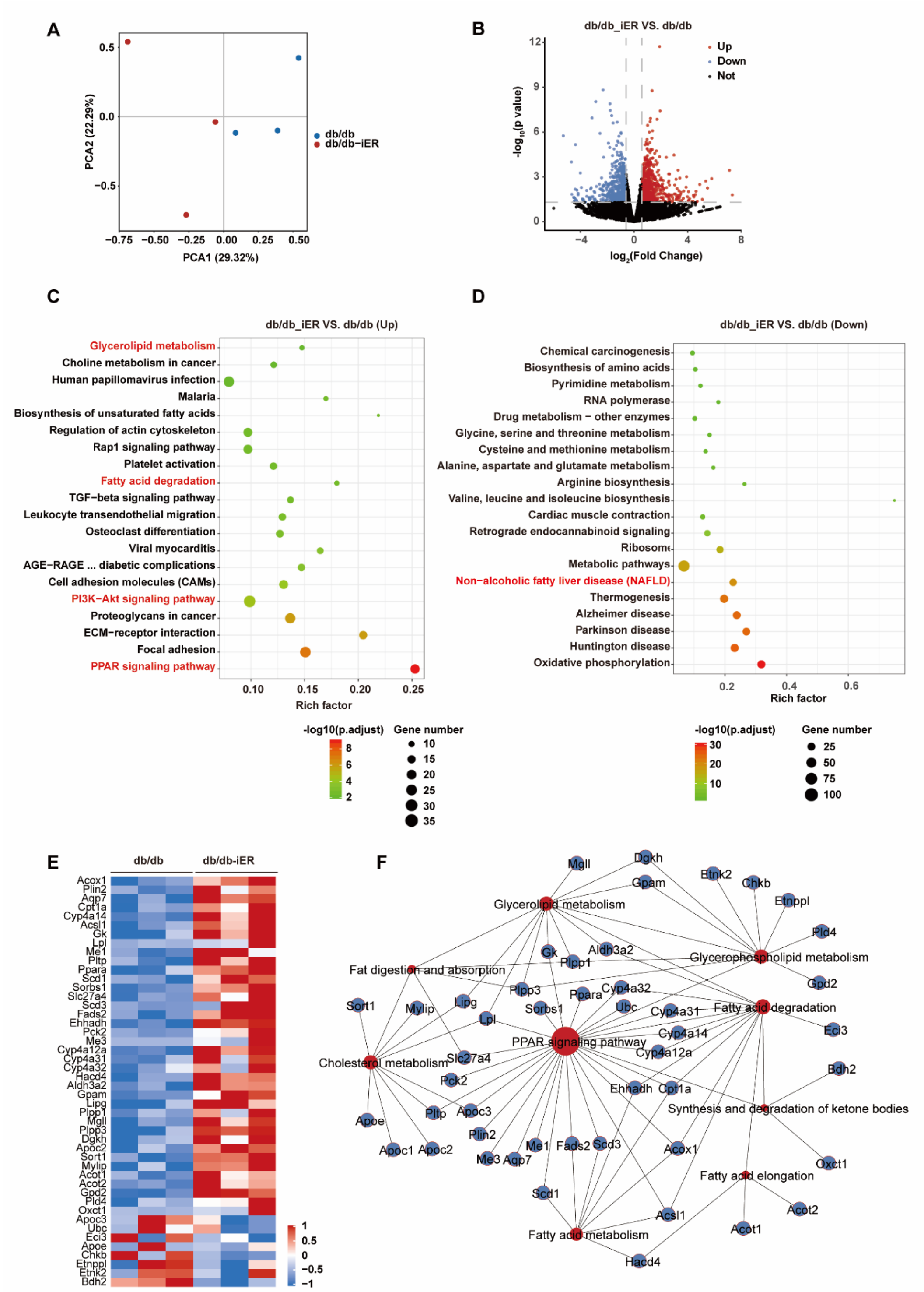


**Figure S7. Transcriptome sequencing analysis reveals alterations in hepatic lipid metabolism by iER intervention in db/db mice, related to Figure 6. (A)** Principal component analysis (PCA) of RNA-Seq data for db/db and db/db_iER groups. **(B)** Volcano plot showing the distribution of p-values ($-\log_{10}$ p value) and fold changes (log $_2$ fold change) in liver transcriptome. **(C-D)** Top 20 most significantly enriched KEGG pathway for up-regulated and down-regulate DEGs, respectively. **(E-F)** Heatmap and KEGG pathway enrichment analysis of DEGs related to hepatic lipid metabolism. Source data are provided as a Source Data file.

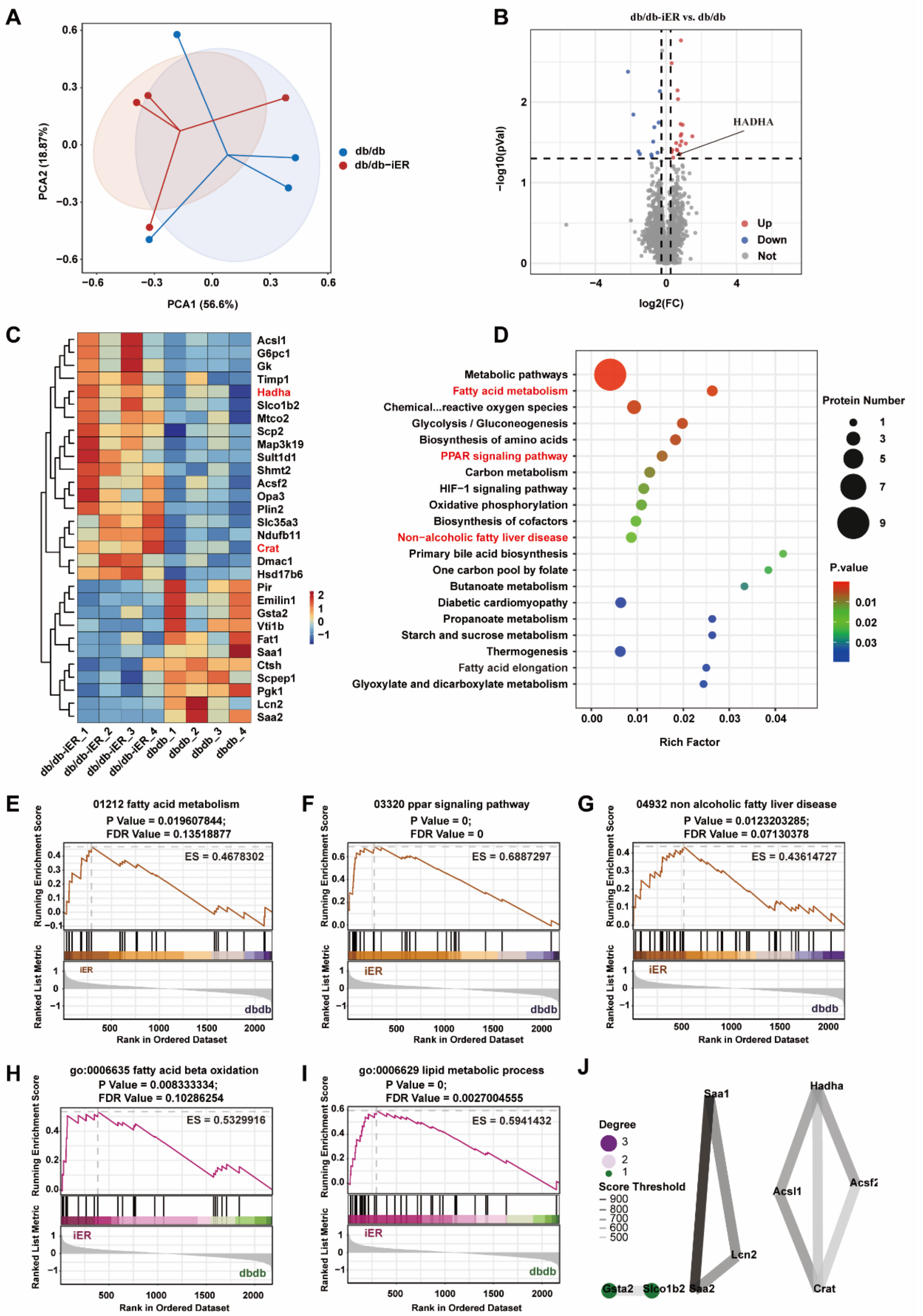


**Figure S8. Liver proteome demonstrates that iER intervention upregulates lipid metabolism-related proteins in db/db mice, related to Figure 6. (A)** Principal component analysis (PCA) of proteomics data for db/db and db/db_iER groups. **(B)** Volcano plot showing log $_2$ fold change versus -log$_{10}$ p value proteomics changes between db/db groups and db/db_iER groups. **(C)** Heatmap of DEPs. **(D)** Top 20 most significantly enriched KEGG pathway. **(E-G)** Gene Set Enrichment Analysis (GSEA)

analysis of fatty acid metabolism, PPAR signaling pathway and non-alcoholic fatty liver disease using KEGG dataset. **(H-I)** GSEA analysis of fatty acid β-oxidation and lipid metabolic process using GO dataset. **(J)** Identification of hub proteins derived from protein-protein interaction (PPI) networks of differential expression acetylated proteins. Source Data are provided as a Source Data file.

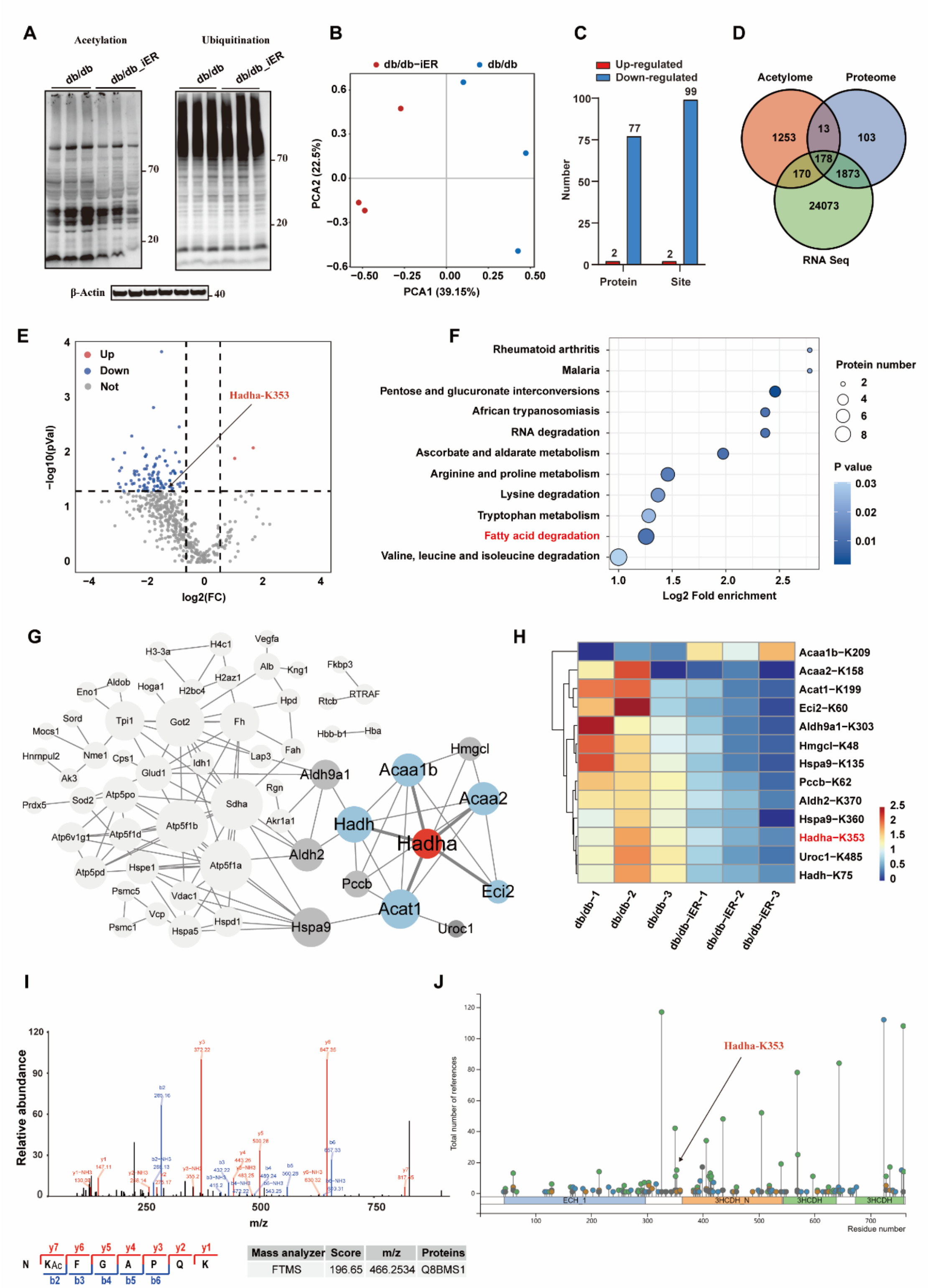


**Figure S9. Liver protein acetylome reveals alteration of fatty acid metabolism by iER intervention in db/db mice, related to Figure 6. (A)** Western blots shows acetylation and ubiquitination modification levels of mouse liver whole-cell lysate using pan-acetylation and ubiquitination antibodies (n = 3 per group). **(B)** Principal component analysis (PCA) of protein acetylome data for db/db and db/db_iER groups. **(C)** Identified differential acetylated sites and proteins were counted. **(D)** Venn diagram showing the overlap of identified all genes, proteins and acetylated proteins from mouse liver multi-omic data. **(E)** Volcano plot showing $\log_2$ fold change versus $-\log_{10}$ p value acetylated proteins changes between db/db groups and db/db_iER groups. **(F)** Significantly enriched KEGG pathways from differentially expressed acetylated proteins. **(G)** Identification of hub acetylated proteins derived from protein-protein interaction networks of differential expression acetylated proteins. **(H)** Heatmap showing direct and indirect interacting proteins of Hadha. **(I)** MS analysis identified HADHA-derived peptides containing acetylated Lys 353 from mouse livers. **(J)** PhosphoSitePlus lollipop plot of mouse Hadha, showing the number of references for the post-translational modified sites throughout the Hadha aminoacidic sequence and Hadha-K353 can be modified by both acetylation and ubiquitination. Source Data are provided as a Source Data file.

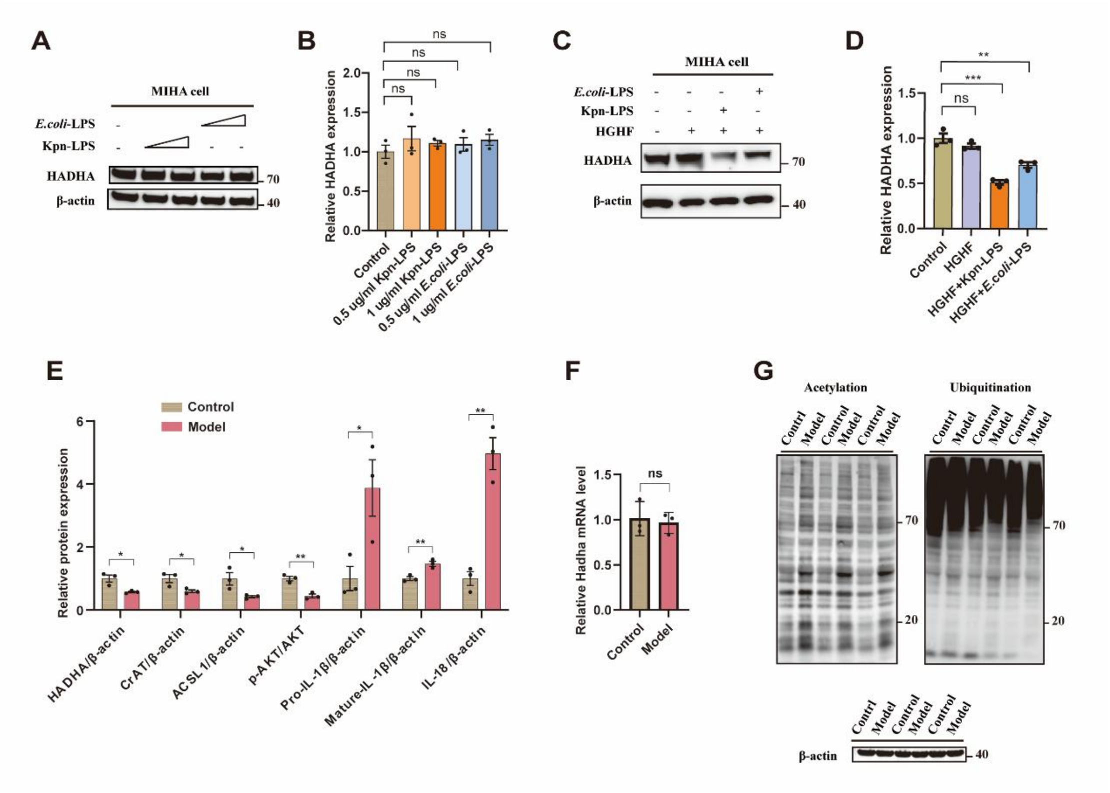


**Figure S10. Effects of LPS combine with HGHF treatment on HADHA expression, pan-acetylated/ubiquitination modifications, insulin resistance and inflammation in MIHA cells, related to Figure 7. (A-B)** Western blotting analysis of HADHA expression in *K. pneumoniae* derived LPS or *E. coli* derived LPS treated MIHA cells. **(C-D)** Western blotting analysis of HADHA expression in HGHF combined with *K. pneumoniae* or *E. coli* derived LPS treated MIHA cells. **(E)** Quantification of Western blotting for control group and model group(HGHF + Kpn-LPS). **(F)** The expression of HADHA at the mRNA level in MIHA cells upon HGHF combined with LPS treatment. **(G)** Detection of pan-acetylated and pan-ubiquitination modifications in HGHF plus Kpn-LPS induced MIHA cells by Western blot analysis. Equal loading was verified using β-actin. Source Data are provided as a Source Data file.

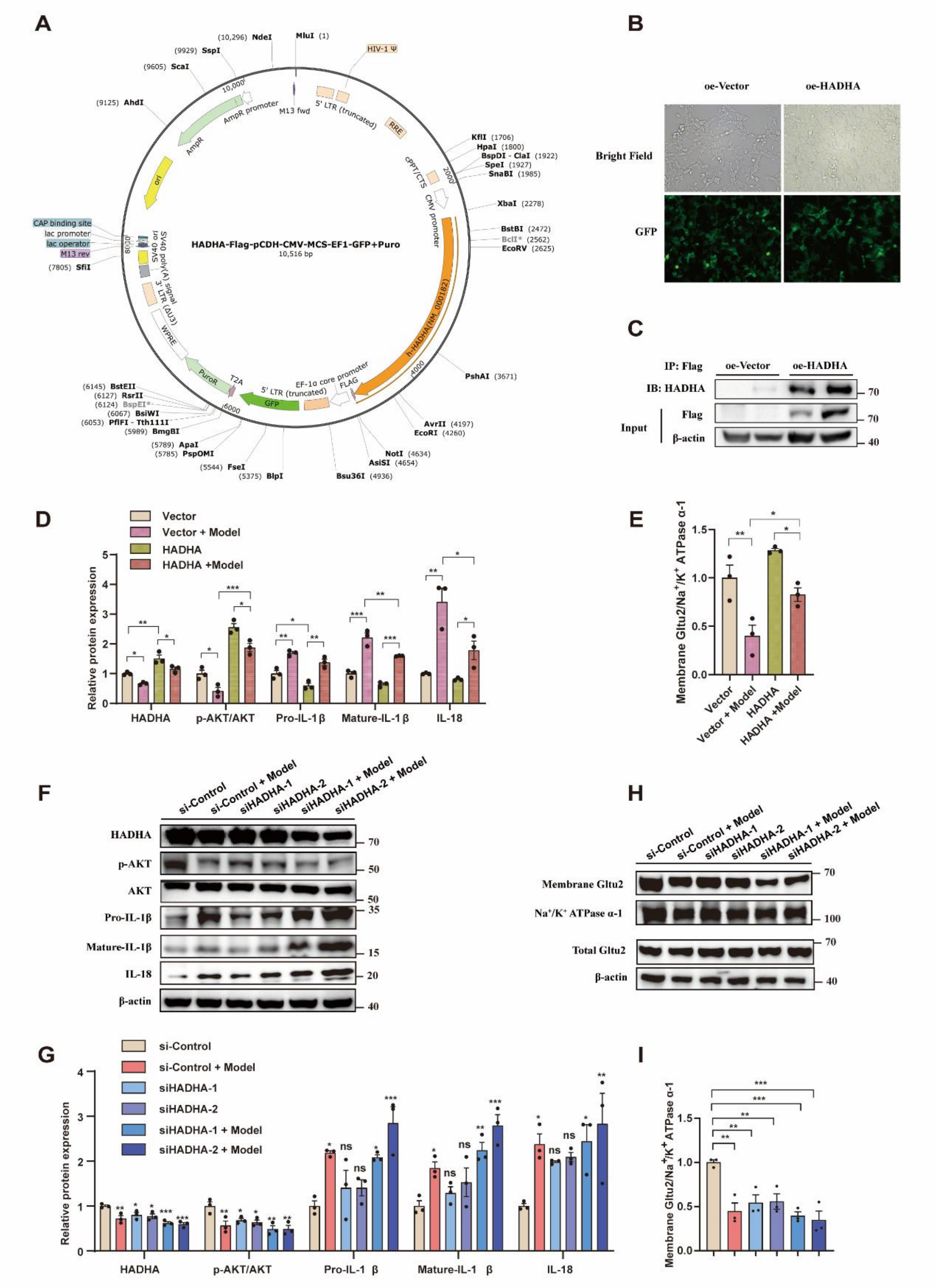


**Figure S11. The effect HADHA overexpression and knockdown on lipid deposition, insulin resistance and inflammation in LPS combine with HGHF-induced MIHA cells, related to Figure 7. (A)** Construction of HADHA-overexpressing lentiviral plasmids encoding HADHA-Flag fusion protein. **(B)** Bright-field and fluorescence images of MIHA cells transfected with a negative control lentivirus (Vector) or a lentivirus overexpressing HADHA (HADHA). **(C)** Expression of the HADHA-Flag fusion protein was verified by western blots. **(D-E)** Quantification of Western blotting for Vector, Vector + Model, HADHA and HADHA+ Model groups. The relative target protein levels were normalized to β-actin for total protein and $Na^{+}$ /$K^{+}$-ATPase for plasma membrane protein. Source Data are provided as a Source Data file. **(F-I)** The expression of p-AKT, AKT, pro-IL-1β, IL-18, HADHA and GLTU2 translocated from cytoplasm to cell membrane in HGHF combined with LPS treated MIHA cells transfected with si-HADHA were examined by Western blotting. Quantification of Western blotting for si-Control, si-Control + Model, siHADHA-1, siHADHA-2, siHADHA-1+ Model and siHADHA-2 + Model groups. β-actin used as a cytosolic loading control. $Na^{+}$/$K^{+}$-ATPase used as a membrane loading control. Asterisks (*) above bars indicate comparisons with controls. Source Data are provided as a Source Data file.

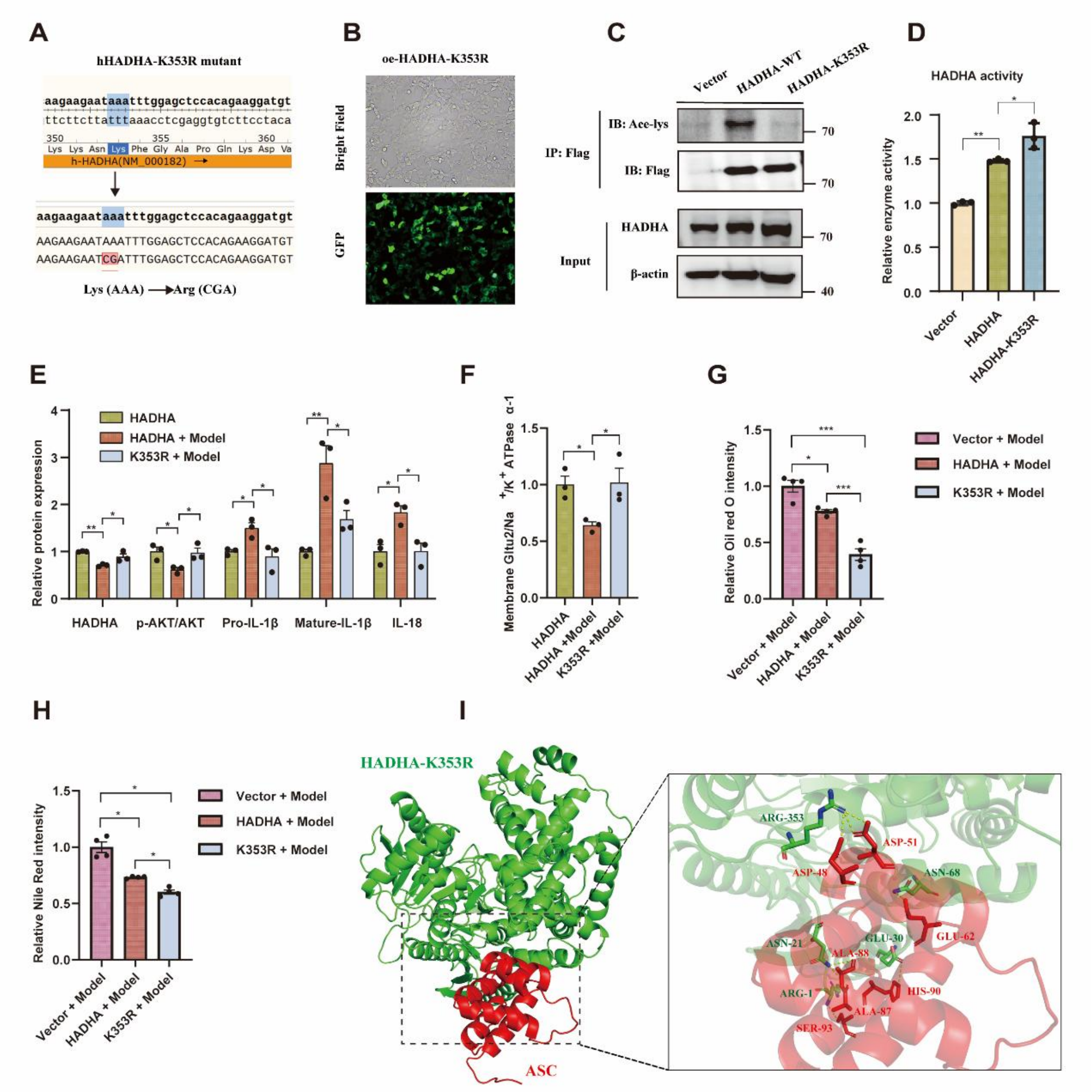


**Figure S12. The effect of HADHA-K353R on lipid deposition, insulin resistance and inflammation in LPS combine with HGHF-induced MIHA cells, related to Figure 7 and Figure 8. (A)** HADHA mutants bearing Lys-to-Arg (K-to-R) substitutions at the acetylated K353 site (HADHA-K353R) were generated to mimic deacetylation overexpression. **(B)** Bright-field and fluorescence images of MIHA cells transfected with a lentivirus overexpressing HADHA-K353R (K353R). **(C)** Immunoprecipitation was performed to verify HADHA deacetylation modifications for HADHA-K353R. **(D)** Comparison of relative enzyme activity in overexpressed HADHA-WT and HADHA-K353R MIHA cells. **(E-F)** Quantification of Western blotting for HADHA, HADHA+ Model and HADHA-K353R + Model groups. **(G)** Oil Red O staining quantitation for Vector + Model, HADHA + Model and K353R + Model groups. **(H)** Nile Red staining quantitation for Vector + Model, HADHA + Model and K353R + Model groups. **(I)** Structure diagram showing the docking model of HADHA-K353R (green)-ASC (red) complex. Source Data are provided as a Source Data file.

### Supplementary Tables

#### Table S1. Key resources in current study.

| REAGENT or RESOURCE | SOURCE | IDENTIFIER |
|---|---|---|
| **Antibodies** | | |
| ACSL1 Rabbit pAb | Abclonal | Cat#A16253； RRID:AB_2768225 |
| IL1β Rabbit pAb | Abclonal | Cat#A1112； RRID:AB_2758416 |
| β-Actin Rabbit mAb | Abclonal | Cat#AC026； RRID:AB_2768234 |
| HADHA Polyclonal antibody | Proteintech | Cat#10758-1-AP； RRID:AB_2115593 |
| CrAT Polyclonal antibody | ProteinTech | Cat#15170-1-AP; RRID:AB_2229978 |
| IL-18 Polyclonal antibody | Proteintech | Cat#10663-1-AP； RRID: AB_2123636 |
| ASC/TMS1 Polyclonal antibody | Proteintech | Cat#10500-1-AP； RRID: AB_2174862 |
| DYKDDDDK tag Monoclonal antibody | Proteintech | Cat#66008-4-Ig； RRID: AB_2918475 |
| Phospho-AKT (Ser473) Monoclonal antibody | Proteintech | Cat#66444-1-lg； RRID:AB_2782958 |
| AKT Polyclonal antibody | Proteintech | Cat#10176-2-AP； RRID:AB_2224574 |
| ATP1A1 Polyclonal antibody | Proteintech | Cat#14418-1-AP; RRID:AB_2227873 |
| Rabbit IgG Isotype Control | Proteintech | Cat#30000-0-AP; RRID:AB_2819035 |
| Mouse IgG1 Isotype Control | Proteintech | Cat#66360-1-Ig; RRID:AB_2827991 |
| Anti-Acetyllysine Mouse mAb | PTM BIO | Cat#PTM-101； RRID:AB_2940830 |
| Anti-Ubiquitin Rabbit mAb | PTM BIO | Cat#PTM-7311; RRID:AB_3677290 |
| Anti-GLUT2 Rabbit mAb | PTM BIO | Cat#PTM-6209 |
| Anti-HSP60 Rabbit mAb | PTM BIO | Cat#PTM-5685 |
| Anti-NFκB p65 Mouse mAb | PTM BIO | Cat#PTM-5591 |
| Goat Anti-Mouse IgG H&L (Alexa Fluor® 488) | Abcam | Cat# ab150113; RRID:AB_2576208 |
| Goat Anti-Rabbit IgG H&L (Alexa Fluor® 555) | Abcam | Cat# ab150078; RRID:AB_2722519 |
| HRP-labeled goat anti-mouse IgG (H+L) antibody | Beyotime | Cat# A0216, RRID:AB_2860575 |
| HRP-labeled Goat Anti-Rabbit IgG (H+L) antibody | Beyotime | Cat# A0208, RRID:AB_2892644 |
| IPKine™ HRP, Goat Anti-Rabbit IgG HCS | Abbkine | Cat# A25222； RRID:AB_2922982 |
| **Biological Samples** | | |
| Human serum | This paper | N/A |
| Human stool sample | This paper | N/A |
| Mouse serum | This paper | N/A |
| Mouse liver tissue | This paper | N/A |
| Mouse colon and ileum tissue | This paper | N/A |
| Mouse colon and ileum contents | This paper | N/A |
| **Chemicals** | | |
| Fetal bovine serum | Vivacell | Cat# C04001 |
| Opti-MEM | Invitrogen | Cat# 31985070 |
| RPMI-1640 | Gibco | Cat# C11875500 |

| | | |
|---|---|---|
| Trypsin–EDTA | Gibco | Cat # 25200056 |
| Penicillin-Streptomycin | Gibco | Cat.# 15140-122 |
| $D^+$ Glucose | Sigma-Aldrich | Cat#G7021 |
| Insulin | Sigma-Aldrich | Cat#I6634 |
| HydroGel | Clear $H_2O$ | Cat# 70-01-5022 |
| Sodium oleate | Sigma-Aldrich | Cat#O7501 |
| Palmitic acid | Sigma-Aldrich | Cat#P0500 |
| DMSO | Sigma-Aldrich | Cat#D8418 |
| LPS | Sigma-Aldrich | Cat#L2880 |
| NAD | Roche | Cat#NAD99-RO |
| Brefeldin A | MCE | Cat#HY-16592 |
| ATP disodium salt | MCE | Cat#HY-B0345A |
| 3-Hydroxypalmitoyl-CoA | Chemsoon | Customized |
| Poly (ethylene glycol) 8000 | Sigma-Aldrich | Cat#89510 |
| Suberoylanilide hydroxamic acid (SAHA) | Sigma-Aldrich | Cat#SML0061 |
| Nicotinamide (NAM) | Sigma-Aldrich | Cat#72340 |
| Oil red O | Sigma-Aldrich | Cat#O0625 |
| BSA (Fatty Acid & IgG Free, BioPremium) | Beyotime | Cat#ST025 |
| Nile Red | Beyotime | Cat#C2051S |
| Lipo8000 | Beyotime | Cat#C0533 |
| PMSF | Beyotime | Cat#ST506 |
| RIPA Lysis Buffer | Beyotime | Cat# P0013B |
| Cell lysis buffer for Western and IP | Beyotime | Cat#P0013 |
| BODIPY 493/503 ( #D3922). | Invitrogen | Cat#D3922 |
| TriZol | Invitrogen | Cat# T9424 |
| Hoechst 33342 | Beyotime | Cat#C1026 |
| Puromycin Dihydrochloride | Beyotime | Cat#ST551 |
| Ampicillin sodium salt | Solarbio | Cat#A8180 |
| MacConkey Inositol Adonitol Carbenicillin | Huankai | |
| Agar | Microbial | Cat#22147 |
| CCK8 | Vazyme | Cat#A311 |
| **Critical Commercial Assays** | | |
| Human IL-1β ELISA Kit | Elabscience | Cat#E-EL-H0149 |
| Human IL-18 ELISA Kit | Elabscience | Cat#E-EL-H0253c |
| Human LPS ELISA Kit | Cusabio | Cat#CSB-E09945h |
| Mouse LPS ELISA Kit | Cusabio | Cat#CSB-E13066m |
| Glucose oxidase–peroxidase assay kit | Nanjing Jiancheng | Cat# A154-1-1 |
| TIANamp Bacteria DNA Kit | TIANGEN | Cat#DP302 |
| LPS extraction kit | iNtRON | Cat#17141 |
| BCA Protein Assay Kit | Beyotime | Cat# P0012S |
| Tissue Mitochondria Isolation Kit | Beyotime | Cat#C3606 |
| Cell Mitochondria Isolation Kit | Beyotime | Cat#C3601 |
| Membrane and Cytosol Protein Extraction Kit | Beyotime | Cat#P0033 |

| | | |
|---|---|---|
| Fast Silver Stain Kit | Beyotime | Cat#P0017S |
| EndoFree Midi Plasmid Kit | TIANGEN | Cat#DP108 |
| Mut Express II Fast Mutagenesis Kit V2 | Vazyme | Cat#C214 |
| Dynabeads™ Protein G Immunoprecipitation Kit | Invitrogen | Cat#10007D |
| Calcium Phosphate Cell Transfection Kit | Beyotime | Cat#C0508 |
| Polybrene | Beyotime | Cat#C0351 |
| Fast Silver Stain Kit | Beyotime | Cat#P0017S |
| Gram-staining kit | Solarbio | Cat#G1060 |
| **Experimental Models: Organisms/Strains** | | |
| Mouse: C57BLKS/JGpt | Nanjing, GemPharmatech | N/A |
| Mouse: BKS.Cg-+Leprdb/+Leprdb/Jcl(db/db) | Nanjing, GemPharmatech | N/A |
| *Klebsiella pneumoniae* | This paper | N/A |
| *Escherichia coli* | This paper | N/A |
| MIHA cells | Hunan Fenghui Biotechnology | N/A |
| HEK293T cells | ATCC | Cat# CRL-1573; RRID: CVCL_0045 |
| **Oligonucleotides** | | |
| Primers for qPCR, See **Table S10** | This paper | N/A |
| pCDH-CMV-MCS-EF1-GFP+Puro HADHA | Hunan Fenghui Biotechnology | N/A |
| pCDH-CMV-MCS-EF1-GFP+Puro vector | Hunan Fenghui Biotechnology | N/A |
| psPAX2 | Hunan Fenghui Biotechnology | N/A |
| pMD2.G | Hunan Fenghui Biotechnology | N/A |
| **Software and Algorithms** | | |
| GraphPad Prism 10.0 | GraphPad Software | https://www.graphpad.com/ |
| ImageJ (v1.42) | National Institutes of Health | https://imagej.nih.gov/ij/ |
| R (v3.6.3) | R Team | https://www.r-project.org |
| Adobe Illustrator | Adobe systems | https://www.adobe.com/ |
| **Other:** | | |
| Human CMNT diet, see **Table S7-S8** | This study | N/A |
| Mouse regular chow diet, see **Table S10** | Hunan Silaike Jinda | N/A |
| Mouse CMNT diet, see **Table S9** | This study | N/A |

**Table S2. Baseline characteristics of the randomized participants, related to Figure 1.**

| Characteristics | Control (n=24) | iER (n=24) | p value |
|---|---|---|---|
| **Anthropometric parameters** | | | |
| Age (years) | 49.96 ± 14.31 | 49.17 ± 12.46 | 0.8389 |
| Male, n (%) | 15 (62.5%) | 17 (70.83%) | 0.5403 |
| Duration of T2DM (years) | 4.5 (1.75-9.25) | 3 (1.75-7) | 0.6690 |
| SBP (mmHg) | 124 (113.5–136.75) | 133.5 (119.5–139.25) | 0.2008 |
| DBP (mmHg) | 81.29 ± 9.23 | 84.33 ± 8.73 | 0.2467 |
| Weight (kg) | 69.6 (64.43–81.13) | 73 (67–83.08) | 0.4704 |
| BMI (kg/$m^2$) | 28.12 ± 4.45 | 27.19 ± 3.49 | 0.4268 |
| WC (cm) | 91.65 ± 9.38 | 91.54 ± 8.16 | 0.9658 |
| HC (cm) | 98.57 ± 8.24 | 98.13 ± 6.35 | 0.8379 |
| WHR | 0.93 ± 0.07 | 0.93 ± 0.06 | 0.9030 |
| **Hepatic steatosis** | | | |
| CAP value | 276.67 ± 36.63 | 295.71 ± 38.39 | 0.0972 |
| HSI | 41.55±5.04 | 40.57 ± 5.38 | 0.5171 |
| FLI | 58.86 ± 28.13 | 57.61 ± 26.26 | 0.8759 |
| **Glucose metabolism** | | | |
| HbA1c (%) | 7.5 (6.4–8.13) | 7.1(6.38–7.85) | 0.7569 |
| FPG (mmol/L) | 7.85 (6.37–9.63) | 7.26(6.47–9.11) | 0.5567 |
| FINS (μIU/ml) | 16 (10–21.9) | 13.87 (11.79–17.33) | 0.7175 |
| HOMA-IR | 4.94 (3.77–8.85) | 5.24 (3.53–6.83) | 0.8900 |
| HOMA-B | 67.84 (33.65–140.41) | 78.58 (47.93–150.85) | 0.5584 |
| C-Peptide (ng/ml) | 2.98 (2.06–3.72) | 2.89 (2.63–3.45) | 0.4617 |
| IGF-1, ng/ml | 122.14 ± 47.9 | 118.5 ± 42.32 | 0.7874 |
| **Liver enzyme and related function** | | | |
| γ-GT (U/L) | 29.05 (22.18–53.95) | 32.25 (26.2–58.05) | 0.4393 |
| ALT (U/L) | 25.75 (18.9–34.93) | 30.8 (18.75–48.63) | 0.4704 |
| AST (U/L) | 19.75 (16.88–24.35) | 20.75 (16.8–34.73) | 0.4394 |
| ALP (U/L) | 68 (57–84.25) | 74 (58.75–85.25) | 0.6440 |
| ALB (g/L) | 43.19 ± 2.35 | 43.63 ± 2.48 | 0.5292 |
| GLB (g/L) | 32.54 ± 6.38 | 28.89 ± 4.8 | 0.0301 |
| PLT(10^9/L) | 227.95 ± 60.02 | 230.58 ± 63.36 | 0.8861 |
| CHE | 9124.1 ± 1339.68 | 9285.75 ± 1151.66 | 0.6655 |
| TBIL (umol/L) | 14.8 (12.03–17.4) | 14.8 (11.68–16.4) | 0.8527 |
| DBIL (umol/L) | 3.7 (2.55–4.15) | 3.7 (2.78–4.45) | 0.7413 |
| TP (g/L) | 75.73 ± 6.07 | 72.53 ± 5.18 | 0.0550 |
| TBA (umol/L) | 3.5 (2.25–5.15) | 4.35 (2.58–5.75) | 0.2397 |
| IBIL (umol/L) | 10.8 (8.65–13) | 11.3 (8.1–12.35) | 0.8431 |
| HCY (umol/L) | 10.73 ± 3 | 11.45 ± 4.66 | 0.5333 |

*Continued on next page*

**Table S2.** ***Continued***

| **Characteristics** | Control (n=24) | iER (n=24) | p value |
|---|---|---|---|
| **Liver fibrosis** | | | |
| LSM value | 6.5 (5.2–7.3) | 5.85 (4.4–7.08) | 0.4126 |
| FIB-4 | 0.92 (0.54–1.16) | 1.03 (0.66–1.3) | 0.5454 |
| NFS | 1.13 ± 1.75 | 1.66 ± 1.18 | 0.2443 |
| DiabetesLiver score | 2.0 ± 0.94 | 3.03 ± 1.11 | 0.6682 |
| FNI | 0.15 (0.09–0.39) | 0.3 (0.13–0.49) | 0.2048 |
| BARD score | 2 (1–3.25) | 2 (2–3) | 0.9064 |
| IV-C (ng/ml) | 40.84 (30.57–51.18) | 24.77 (19.06–43.27) | 0.0453 |
| HA (ng/ml) | 59.04 (47.6–82.6) | 63.06 (44.55–86.11) | 0.9637 |
| LN (ng/ml) | 65.16 (46.78–81.05) | 29.38 (23.6–65.63) | 0.0818 |
| PC-III (ng/ml) | 8.38 (5.39–13.18) | 9.97 (7.27–18.06) | 0.2601 |
| **Blood lipid** | | | |
| TG (mmol/L) | 1.48 (1.03–4.87) | 1.96 (1.49–2.62) | 0.7028 |
| TC (mmol/L) | 4.84 ± 1.15 | 4.62 ± 1.09 | 0.4869 |
| HDL (mmol/L) | 1.23 ± 0.22 | 1.14 ± 0.23 | 0.1381 |
| LDL (mmol/L) | 2.86 ± 0.78 | 2.84 ± 0.79 | 0.9195 |
| Apo A1 (g/L) | 1.4 ± 0.26 | 1.32±0.21 | 0.2317 |
| Apo B (g/L) | 1.03 ± 0.25 | 1.02 ± 0.24 | 0.9583 |
| **Inflammation-related factors** | | | |
| hs-CRP (mg/L) | 2.75(1.42–3.95) | 2.22(1.36–6.02) | 0.9918 |

Values are the mean ± SD (data with normal distribution), median (interquartile range (IQR): 25%-75%) (data with non-normal distribution), or number of patients (percentage of total) (data with proportions). The p-values are for the between-group comparisons by Mann-Whitney U test (data with non-normal distribution), unpaired Student's t test (data with normal distribution) or chi-square tests (data with proportions) for categorical variables, respectively.

Abbreviations: iER, intermittent energy restriction; T2DM, type 2 diabetes mellitus; SBP, systolic blood pressure; DBP, diastolic blood pressure; BMI, body mass index; WC, waist circumference; HC, hip circumference; WHR, waist-to-hip ratio; CAP value, controlled attenuation parameter by transient elastography; HIS, hepatic steatosis index; FLI, fatty liver index; HbA1c, glycated hemoglobin; FPG, fasting plasma glucose; FINS, fasting insulin; HOMA-IR, homeostasis model assessment of insulin resistance; HOMA-B, homeostasis model assessment of β-cell function; IGF-1, insulin-like growth factor 1; γ-GT, γ-glutamyl transpeptidase; ALT, alanine aminotransferase; AST, aspartate aminotransferase; ALP, alkaline phosphatase; ALB, albumin; GLB, globulin; PLT, platelets; CHE, cholinesterase; TBIL, total bilirubin; DBIL, direct bilirubin; IBIL, indirect bilirubin; TP, total protein; TBA, total bile acids; HCY, homocysteine; LSM value, liver stiffness measurement by transient elastography; FIB-4, fibrosis-4 index; NFS, non-alcoholic fatty liver disease fibrosis score; FNI, fibrotic non-alcoholic steatohepatitis index; BARD score, BMI, AST/ALT ration, and Diabetes score; IV-C, Type IV Collagen; HA, hyaluronic acid; LN, laminin; PC-III, Procollagen III; TG,

Triglycerides; TC, Total Cholesterol; HDL, high density lipoprotein; LDL, low density lipoprotein; Apo A1, Apolipoprotein A1; Apo B, Apolipoprotein B; hs-CRP, high-sensitivity C-reactive protein.

**Table S3. Changes in clinical characteristics from pre- to post-control or iER among completed participants, related to Figure 1.**

| Characteristics | Control | | | | | iER | | | | | Δ-p value |
|---|---|---|---|---|---|---|---|---|---|---|---|
| | n* | Pre | Post | Δ | p value | n* | Pre | Post | Δ | p value | |
| **Anthropometric parameters** | | | | | | | | | | | |
| SBP (mmHg) | 19 | 125.21 ± 17.06 | 125 ± 12.83 | -0.21±14.52 | 0.9503 | 20 | 132.35±15.96 | 126.6±16.35 | -5.75±14.11 | 0.0842 | 0.2346 |
| DBP (mmHg) | 19 | 82.68 ± 9.34 | 82.37 ± 10.29 | -0.32 ± 9.67 | 0.8884 | 20 | 84.05 ± 9.51 | 84.15 ± 8.24 | 0.1 ± 10.36 | 0.9660 | 0.8977 |
| Weight (kg) | 19 | 68 (64.1 – 84.85) | 69 (63.8 – 83.3) | -0.5 (-1.95 – 0.45) | 0.3441 | 20 | 77.04 ± 13.12 | 74.23 ± 12.82 | -2.05 (-3.5 – -0.77) | 0.0025 | 0.0327 |
| BMI (kg/m$^2$) | 19 | 27.74 ± 4.41 | 27.59 ± 4.73 | -0.17 (-0.78 – 0.17) | 0.5197 | 20 | 27.7 ± 3.41 | 26.68 ± 3.21 | -0.75 (-1.27 – -0.29) | 0.0029 | 0.0543 |
| WC (cm) | 19 | 90 (85 – 96) | 91 (88 – 94.5) | 1.21 ± 4.53 | 0.1817 | 20 | 92.9 ± 7.11 | 89 ± 7.81 | -3.9 ± 3.86 | 0.0002 | 0.0005 |
| HC (cm) | 19 | 96 (91 – 103) | 94 (90.5 – 99) | -1.58 ± 5.39 | 0.3584 | 20 | 98.7 ± 6.25 | 97.6 ± 6.7 | -1.1 ± 6.07 | 0.4276 | 0.7962 |
| WHR | 19 | 0.93 ± 0.07 | 0.96 ± 0.06 | 0.02 (-0.01 – 0.05) | 0.0532 | 20 | 0.94 ± 0.06 | 0.91 ± 0.05 | -0.03 (-0.05 – 0.01) | 0.0018 | 0.0008 |
| **Hepatic steatosis** | | | | | | | | | | | |
| CAP value | 16 | 272.94 ± 32.07 | 278.12 ± 69.15 | 2.56 ± 58.05 | 0.8622 | 19 | 302.6 ± 37.58 | 265.95 ± 43.27 | -35.79 ± 39.43 | 0.0009 | 0.0269 |
| HSI | 19 | 41.45 ± 5.12 | 41.46 ± 5.7 | 0.01 ± 3.06 | 0.9853 | 20 | 41.66 ± 5.12 | 38.78 ± 4.97 | -2.88 ± 3.05 | 0.0005 | 0.0054 |
| FLI | 19 | 56.81 ± 29.06 | 58.56 ± 26.29 | 3.18 (-6.77 – 10.63) | 0.5846 | 20 | 60.85 (44.38 – 79.18) | 37.25 (27.86 – 77.43) | -13.64 (-21.8 – -3.12) | 0.0012 | 0.0030 |
| **Glucose metabolism** | | | | | | | | | | | |

*Continued on next page*

**Table S3.** ***Continued***

| Characteristics | Control | | | | | iER | | | | | Δ-p value |
|---|---|---|---|---|---|---|---|---|---|---|---|
| | n* | Pre | Post | Δ | p value | n* | Pre | Post | Δ | p value | |
| HbA1c (%) | 19 | 7.6 (6.5 ‑ 8.15) | 7.4 (6.85 ‑ 8.15) | 0.25 ± 1.35 | 0.3978 | 20 | 7.25 (6.48 ‑ 8.48) | 6.1 (5.73 ‑ 6.48) | -1.5 ± 1.33 | 0.0002 | 0.0002 |
| FPG (mmol/L) | 19 | 7.81 (6.45 ‑ 10.29) | 7.83 (7.11 ‑ 9.66) | -0.4 (-1.48 ‑ 0.95) | 0.6435 | 20 | 7.3 (6.7 ‑ 9.11) | 6.41 (5.52 ‑ 7.35) | -1.06 (-3.24 ‑ -0.16) | 0.0036 | 0.1402 |
| FINS (μIU/ml) | 18 | 14.37 (8.14 ‑ 20.09) | 9.83 (7.26 ‑ 13.96) | -1.51 (-5.4 ‑ 1.32) | 0.1770 | 20 | 14.17 (11.76 ‑ 17.33) | 12.67 (8.62 ‑ 15.96) | -1.62 (-4.48 ‑ -0.1) | 0.0068 | 0.6295 |
| HOMA-IR | 18 | 4.51 (3.03 ‑ 7.31) | 3.78 (2.55 ‑ 4.85) | -1.23 ± 2.78 | 0.0979 | 20 | 5.24 (3.6 ‑ 6.87) | 3.1 (2.58 ‑ 4.96) | -1.57 ± 1.49 | 0.0004 | 0.6467 |
| HOMA-B | 18 | 65.17 (30.83 ‑ 109.46) | 36.77 (20.08 ‑ 88.47) | 24.76 ± 126.44 | 0.4860 | 20 | 78.58 (47.93 ‑ 133.91) | 83.75 (66.03 ‑ 131.31) | 10.59 ± 43.67 | 0.1305 | 0.1111 |
| C-Peptide (ng/ml) | 18 | 2.67 ± 1.22 | 2.74 ± 0.89 | 0.07 ± 1.05 | 0.7923 | 20 | 2.89 (2.56 ‑ 3.61) | 2.52 (1.93 ‑ 3.4) | -0.5 ± 0.72 | 0.0031 | 0.0609 |
| IGF-1 (ng/ml) | 16 | 126.5 (75.78 ‑ 149) | 81.39 (62.22 ‑ 105.69) | -35.48 (-55.04 ‑ -3) | 0.0162 | 20 | 109.41 ± 37.22 | 94.78 ±39.71 | -11.48 (-38.47 ‑ 2.91) | 0.1556 | 0.2721 |
| **Liver enzyme and related function** | | | | | | | | | | | |
| γ-GT (U/L) | 19 | 29.3 (21.35 ‑ 54.3) | 33 (22.7 ‑ 50.41) | 1.5 (-2.2 ‑ 6.65) | 0.2514 | 20 | 35.6 (26.4 ‑ 73.45) | 24.6 (18.13 ‑ 35.85) | -9.25 (-13.35 ‑ -1.73) | 0.0057 | 0.0054 |
| ALT (U/L) | 19 | 25.1 (18.9 ‑ 32.8) | 24.6 (18.65 ‑ 29.1) | -0.8 (-4.6 ‑ 4.5) | 0.8248 | 20 | 34.75 (21.6 ‑ 62.43) | 20.35 (17.1 ‑ 42.65) | -11.5 (-23.28 ‑ -0.28) | 0.0043 | 0.0204 |
| AST (U/L) | 19 | 19.2 (16.4 ‑ 23.3) | 19.1 (16.4 ‑ 21.4) | -0.6 (-3.1 ‑ 1.5) | 0.7022 | 20 | 21.75 (18.03 ‑ 42.93) | 20.3 (17.75 ‑ 26.35) | -1.45 (-11.25 ‑ 3.3) | 0.1126 | 0.2793 |
| ALP (U/L) | 17 | 67.53 ± 20.53 | 65.41 ± 22.4 | -2.12 ± 9.97 | 0.3940 | 20 | 74 (62 ‑ 82.5) | 55.5 (49 ‑ 71.5) | -11.5 ± 15.62 | 0.0074 | 0.0400 |
| ALB (g/L) | 19 | 43.47 ± 2.13 | 44.14 ± 2.68 | 0.67 ± 1.54 | 0.0724 | 20 | 43.54 ± 2.33 | 44.07 ± 1.92 | 0.53 ± 1.67 | 0.1709 | 0.7815 |

*Continued on next page*

**Table S3.** ***Continued***

| Characteristics | Control | | | | | iER | | | | | Δ-p value |
|---|---|---|---|---|---|---|---|---|---|---|---|
| | n* | Pre | Post | Δ | p value | n* | Pre | Post | Δ | p value | |
| GLB (g/L) | 19 | 31.31 ± 6.3 | 27.41 ± 3.89 | -3.89 ± 5.74 | 0.0084 | 20 | 27.35 (25.18 – 28.93) | 26.35 (24.6 – 27.03) | -1.71 ± 4.07 | 0.1559 | 0.1759 |
| PLT($10^{\wedge 9}$/L) | 17 | 235.41 ± 57.94 | 237.82 ± 51.43 | 2.41 ± 23.75 | 0.6810 | 20 | 235.15 ± 67.86 | 225.3 ± 73.39 | -9.85 ± 39.65 | 0.2804 | 0.2548 |
| CHE ( U/L) | 16 | 8938 ± 1201.42 | 9429.81 ± 1399.81 | 491.81 ± 1273.65 | 0.1433 | 20 | 9387.85 ± 1004.25 | 8422.4 ± 646.6 | -965.45 ± 1015.66 | 0.0004 | 0.0005 |
| TBIL (umol/L) | 19 | 15.16 ± 3.85 | 14.45 ± 3.84 | -1 (-2.8 – 1.65) | 0.3119 | 20 | 13.9 (11.18 – 16.4) | 15 (13.1 – 18.35) | 0.75 (-1.15 – 3.4) | 0.1913 | 0.1061 |
| DBIL (umol/L) | 19 | 3.67 ± 1.28 | 3.79 ± 1.21 | 0.4 (-0.35 – 0.7) | 0.6092 | 20 | 3.55 (2.68 – 4.4) | 4.1 (3.33 – 4.85) | 0.6 (-0.15 – 1.3) | 0.0673 | 0.3116 |
| TP (g/L) | 19 | 74.77 ± 6.11 | 71.55 ± 4.37 | -3.7 (-7.45 – 1.75) | 0.0249 | 20 | 71.29 ± 4.66 | 70.12 ± 4.3 | -0.1 (-2.33 – 1.55) | 0.2910 | 0.2011 |
| TBA (umol/L) | 19 | 3.2 (2 – 4.2) | 3.2 (2.5 – 3.85) | -0.4 (-0.85 – 0.7) | 0.7112 | 20 | 4.35 (2.58 – 6.15) | 3.85 (2.23 – 5.03) | -0.45 (-1.83 – 0.53) | 0.1913 | 0.5456 |
| IBIL (umol/L) | 17 | 11.35 ± 2.96 | 10.44 ± 2.52 | -1.4 (-2.9 – 0.6) | 0.1448 | 20 | 10.25 (8.03 – 12.35) | 11.4 (8.3 – 13.78) | -0.15 (-1.33 – 2.83) | 0.4665 | 0.0907 |
| Hcy (umol/L) | 18 | 10.85±3.28 | 10.19 ± 3.29 | -0.66 ± 4.32 | 0.5256 | 20 | 10.5 (6.83 – 13.63) | 8.95 (6.95 – 11.58) | -1.89 ± 4.06 | 0.0464 | 0.3724 |
| **Liver fibrosis** | | | | | | | | | | | |
| LSM value | 17 | 6.33 ± 1.5 | 5.89 ± 1.74 | -0.9 (-2.16 – 0.68) | 0.1576 | 19 | 5.95 (4.48 – 7.9) | 6 (4.55 – 7.25) | -0.3 (-1.75 – 1.05) | 0.3165 | 0.8946 |
| FIB-4 | 17 | 0.8 ± 0.32 | 0.82 ± 0.37 | -0.01 (-0.1 – 0.11) | 0.7090 | 20 | 0.93 (0.63 – 1.22) | 1.15 (0.69 – 1.36) | 0.06 (-0.1 – 0.15) | 0.4222 | 0.5938 |
| NFS | 17 | -1.34 ± 0.9 | -1.37 ± 1.04 | -0.03 ± 0.38 | 0.7504 | 20 | -1.29 ± 1.31 | -1.16 ± 1.31 | 0.13 ± 0.6 | 0.3513 | 0.3550 |
| DiabetesLiver score | 17 | 2.62 ± 0.77 | 2.69 ± 0.85 | 0.07 ± 0.43 | 0.4956 | 20 | 3.05 (2.41 – 3.36) | 2.6 (2.14 – 3.03) | -0.42 ± 0.73 | 0.0217 | 0.0189 |

*Continued on next page*

**Table S3.** ***Continued***

| Characteristics | Control | | | | | iER | | | | | Δ-p value |
|---|---|---|---|---|---|---|---|---|---|---|---|
| | n* | Pre | Post | Δ | p value | n* | Pre | Post | Δ | p value | |
| FNI | 19 | 0.14 (0.09 – 0.34) | 0.18 (0.11 – 0.35) | 0.01 ± 0.19 | 0.6726 | 20 | 0.32 (0.17 – 0.62) | 0.15 (0.06 – 0.24) | -0.22 ± 0.24 | 0.0006 | 0.0021 |
| BARD score | 19 | 2 (1 – 3) | 2 (1 – 3) | 0 (-0.5 – 0) | 0.7063 | 20 | 2 (1.75 – 3) | 3 (2 – 3.25) | 0 (0 – 2) | 0.0141 | 0.0376 |
| IV-C (ng/ml) | 16 | 35.88 (28.31 – 43.84) | 18.73 (16.53 – 21.72) | -16.08 (-22.62 – -6.71) | 0.0041 | 20 | 23.67 (19.06 – 43.27) | 20.59 (15.29 – 29.78) | -3.28 (-9.78 – 0.11) | 0.0160 | 0.0721 |
| HA (ng/ml) | 16 | 65.17 (47.22 – 85.82) | 67.04 (60.52 – 85.68) | 7.9 (-8.23 – 16.64) | 0.2243 | 20 | 67.05 (46.73 – 93.54) | 70.99 (44.68 – 93.01) | 1.85 (-6.68 – 14.24) | 0.4222 | 0.6217 |
| LN (ng/ml) | 16 | 65.64 (28.81 – 80.41) | 23.06 (21.9 – 27.37) | -32.98 (-55.79 – -6.9) | 0.0041 | 20 | 25.76 (23.01 – 38.8) | 25.48 (20.84 – 29.97) | -3.03 (-11.99 – 2.38) | 0.0458 | 0.0136 |
| PC-III (ng/ml) | 16 | 7.72 (5.17 – 12.68) | 11.43 (8.05 – 17.9) | 2.72±7.85 | 0.2052 | 20 | 12.31 (8.83 – 19.58) | 13.3 (8.42 – 19.35) | 1.78±10.98 | 0.6407 | 0.7751 |
| **Blood lipid** | | | | | | | | | | | |
| TG (mmol/L) | 19 | 1.49 (1.08 – 4.69) | 1.84 (1.47 – 3.07) | 0.32 (-0.97 – 0.58) | 0.8880 | 20 | 1.96 (1.55 – 2.74) | 1.39 (1.08 – 2.61) | -0.15 (-0.78 – 0.23) | 0.3317 | 0.5091 |
| TC (mmol/L) | 19 | 4.97 (4.21 – 5.79) | 5.16 (4.11 – 5.69) | 0.12 (-0.19 – 0.41) | 0.2953 | 20 | 4.7 ± 1.14 | 4.66 ± 1.02 | 0 (-0.34 – 0.27) | 0.8252 | 0.3465 |
| HDL (mmol/L) | 19 | 1.23 (1.15 – 1.38) | 1.14 (1.03 – 1.31) | -0.03(-0.15 – 0.08) | 0.2857 | 20 | 1.13 ± 0.24 | 1.22 ± 0.25 | 0.06 (0.01 – 0.18) | 0.0135 | 0.0212 |
| LDL (mmol/L) | 19 | 2.92 ± 0.79 | 3.11 ± 1.02 | 0.19 ± 0.82 | 0.3201 | 20 | 2.86 ± 0.83 | 2.79 ± 0.72 | -0.08 ± 0.62 | 0.5833 | 0.2524 |
| Apo A1 (g/L) | 16 | 1.42 ± 0.27 | 1.31 ± 0.15 | -0.11 ± 0.2 | 0.0376 | 20 | 1.28 ± 0.2 | 1.34 ± 0.21 | 0.06 ± 0.16 | 0.1337 | 0.0077 |
| Apo B (g/L) | 18 | 1.04 ± 0.25 | 1.01 ± 0.25 | -0.03 ± 0.22 | 0.5579 | 20 | 1.01 ± 0.25 | 0.92 ± 0.21 | -0.09 ± 0.22 | 0.0823 | 0.4235 |

*Continued on next page*

**Table S3.** ***Continued***

| Characteristics | Control | | | | | iER | | | | | Δ-p value |
|---|---|---|---|---|---|---|---|---|---|---|---|
| | n* | Pre | Post | Δ | p value | n* | Pre | Post | Δ | p value | |
| **Inflammation-related factors** | | | | | | | | | | | |
| hs-CRP (mg/L) | 19 | 2.05 (1.24 – 3.17) | 1.56 (1.01 – 2.44) | -0.12 (-1.33 – 0.14) | 0.1589 | 20 | 2.14 (0.98 – 6.77) | 1.06 (0.76 – 4.25) | -0.98 (-3 – -0.38) | 0.0006 | 0.0946 |
| IL-1β (pg/ml) | 19 | 15.25 (14.18 – 18.58) | 16.04 (14.72 – 29.36) | 0.27 (-0.01 – 1.30) | 0.0105 | 20 | 17.64 (15.66 – 41.12) | 15.03 (14.33 – 17.42) | -2.03 (-19.94 – -0.41) | <0.0001 | <0.0001 |
| IL-18 (pg/ml) | 19 | 931.53 (649.76 – 1041.70) | 993.29 (928.44 – 1017.60) | 57.06 (-59.91 – 231.32) | 0.1688 | 20 | 1016.50 (953.00 – 1060.80) | 809.32 (665.94 – 936.31) | -198.53 (-322.35 – -111.03) | <0.0001 | <0.0001 |
| LPS (pg/ml) | 19 | 128.17 (59.65 – 146.47) | 119.12 (73.98 – 139.37) | 10.77±64.85 | 0.2352 - | 20 | 160.91±65.82 | 125.08±63.51 | -35.83±22.42 | <0.0001 | 0.0071 - |

All the data are presented as the mean ± SD (data with normal distribution) or median (interquartile range (IQR): 25%-75%) (data with non-normal distribution). *Number of participants with data available at pre and post- control or iER for each outcome. The outcome variables from pre to post-control or iER are represented by "Δ" in the table. Within-group comparisons for pre-versus post-control or iER were performed by paired t tests (data with normal distribution) or paired Wilcoxon tests (data with non-normal distribution), and between-group comparisons of changes were tested (Δ-p value) by independent sample t tests (data with normal distribution) or Mann-Whitney U tests (data with non-normal distribution).

**Table S4. Comparison of post-intervention outcomes between Control and iER groups using ANCOVA, related to Figure 1.**

| Outcome index | Δ $Control_{adj}$ | Δ $iER_{adj}$ | p value |
|---|---|---|---|
| SBP (mmHg) | -1.96±2.81 | -4.09±2.74 | 0.5945 |
| DBP (mmHg) | -0.72±1.96 | 0.49±1.91 | 0.6616 |
| Body Weight (kg) | -0.23±0.76 | -2.83±0.74 | 0.0197 |
| BMI (kg/m2) | -0.15±0.28 | -1.02±0.27 | 0.0305 |
| Waist Circumference (cm) | 1.13±0.96 | -3.82±0.94 | 0.0008 |
| Hip Circumference (cm) | -1.72±1.22 | -0.96±1.19 | 0.6585 |
| Waist-to-hip ratio | 0.02±0.01 | -0.03±0.01 | 0.0002 |
| CAP value by transient elastography | -1.62±17.82 | -46.6±15.79 | 0.0680 |
| Hepatic Steatosis Index (HSI) | 0±0.69 | -2.86±0.67 | 0.0054 |
| Fatty Liver Index (FLI) | 1.34±3.78 | -14.73±3.68 | 0.0044 |
| HbA1c (%) | 0.2±0.25 | -1.45±0.24 | 0.0000 |
| Fasting plasma glucose (mmol/L) | 0.09±0.51 | -1.95±0.49 | 0.0066 |
| Fasting plasma insulin (μIU/ml) | -2.31±1.16 | -2.11±1.1 | 0.8971 |
| HOMA-IR | -1.15±0.47 | -1.64±0.45 | 0.4522 |
| C-Peptide (ng/ml) | 0.13±0.17 | -0.5±0.17 | 0.0146 |
| Insulin-like Growth Factor 1 (IGF-1, ng/ml) | -10.58±16.46 | -18.61±14.71 | 0.7199 |
| γ-GT (U/L) | 1.56±5.43 | -13.12±5.29 | 0.0645 |
| ALT (U/L) | -3.16±3.33 | -11.38±3.24 | 0.0933 |
| AST (U/L) | -2.04±1.92 | -3.58±1.87 | 0.5815 |
| ALP (U/L) | -2.51±3.26 | -11.16±3 | 0.0606 |

*Continued on next page*

**Table S4.** ***Continued***

| Outcome index | Δ $Control_{adj}$ | Δ $iER_{adj}$ | p value |
|---|---|---|---|
| ALB (g/L) | 0.67±0.36 | 0.54±0.35 | 0.7964 |
| GLB (g/L) | -2.7±0.84 | -2.84±0.81 | 0.9087 |
| A/G | 0.16±0.05 | 0.15±0.05 | 0.8247 |
| Platelet count (10^9/L) | 2.43±7.96 | -9.87±7.34 | 0.2644 |
| CHE | 346.66±241.93 | -849.32±215.87 | 0.0009 |
| TBIL (umol/L) | -0.57±0.82 | 0.27±0.8 | 0.4706 |
| DBIL (umol/L) | 0.13±0.24 | 0.37±0.23 | 0.4720 |
| DBIL/TBIL | 0.02±0.01 | 0.02±0.01 | 0.9702 |
| TP (g/L) | -2.05±0.94 | -2.29±0.91 | 0.8559 |
| TBA (umol/L) | -0.49±0.35 | -0.37±0.35 | 0.8212 |
| IBIL (umol/L) | -0.81±0.67 | -0.06±0.62 | 0.4201 |
| Homocysteine (Hcy, umol/L) | -0.73±0.75 | -1.83±0.71 | 0.2963 |
| LSM value by transient elastography | -1.21±0.52 | -1.16±0.47 | 0.5305 |
| Fibrosis-4 index (FIB-4) | 0.01±0.11 | 0.12±0.1 | 0.5060 |
| NAFLD fibrosis score (NFS) | 0.04±0.12 | -0.14±0.11 | 0.2868 |
| FNI | 1.34±3.78 | -14.73±3.68 | 0.0044 |
| BARD score | -0.14±0.21 | 0.63±0.21 | 0.0130 |
| DiabetesLiver score | 0.02±0.15 | -0.38±0.13 | 0.0582 |
| IV-C (ng/ml) | -12.71±2.32 | -7.18±2.13 | 0.0910 |
| HA (ng/ml) | 6.5±7.62 | 7.78±6.81 | 0.9007 |
| LN (ng/ml) | -23.63±4.45 | -16.42±3.96 | 0.2448 |
| PC-III (ng/ml) | 1.48±2.42 | 2.77±2.15 | 0.7014 |

*Continued on next page*

**Table S4.** ***Continued***

| **Outcome index** | **Δ $Control_{adj}$** | **Δ $iER_{adj}$** | **p value** |
|---|---|---|---|
| TG (mmol/L) | 0.26±0.54 | -0.21±0.52 | 0.5397 |
| TC (mmol/L) | 0.3±0.2 | -0.05±0.19 | 0.2146 |
| HDL (mmol/L) | 0.01±0.05 | 0.09±0.05 | 0.3183 |
| LDL (mmol/L) | 0.2±0.16 | -0.09±0.15 | 0.2012 |
| Apo A1 (g/L) | -0.15±0.06 | 0.03±0.06 | 0.0396 |
| Apo B (g/L) | -0.08±0.06 | -0.1±0.05 | 0.2340 |
| A1/B | -0.1±0.09 | 0.13±0.08 | 0.0902 |
| hs-CRP (mg/L) | -0.38±0.73 | -1.79±0.71 | 0.1760 |
| IL-1β (pg/ml) | -2.38±2.24 | -5.69±2.24 | 0.3241 |
| IL-18 (pg/ml) | 116.91±42.07 | -161.62±42.07 | 0.0002 |
| LPS (pg/ml) | 5.98±11.06 | -31.28±10.77 | 0.0247 |

Data are presented as baseline-adjusted changes from baseline to post-intervention (Mean ± SE). The adjusted change is calculated as: $Post_{adj}$ - $Pre_{raw}$, where $Post_{adj}$ represents the estimated marginal means derived from the ANCOVA model. ANCOVA P-value: Derived from Analysis of Covariance with the baseline value and grouping included as covariates. The outcome variables from pre to post-control or iER are represented by "Δ" in the table. Abbreviations: $Control_{adj}$: Adjusted value for the Control group; $iER_{adj}$: Adjusted value for the iER group; $Post_{adj}$: Adjusted post-treatment value; $Pre_{raw}$: baseline value

**Table S5. Dietary information between the iER group and the control group, related to Figure 1.**

| | Control | | | | | iER | | | | | |
|---|---|---|---|---|---|---|---|---|---|---|---|
| **Characteristics** | **n*** | **Pre** | **Post** | **Δ** | **p value** | **n*** | **Pre** | **Post** | **Δ** | **p value** | **Δ-p value** |
| Total energy (kcal/d) | 19 | 1952.97 ± 291.99 | 1965.46 (1764.3–2029.95) | -2.69 (-289.15–131.89) | 0.5396 | 20 | 1906.6 ± 288.74 | 1883.69 (1782.09–2005.89) | 47.72 (-82.98–170.9) | 0.9996 | 0.4565 |
| Carbohydrate (g) | 19 | 221.31 ± 47.75 | 235.89 (174.81–257.89) | -3.29 (-31.64–26.34) | 0.7323 | 20 | 228.49 ± 68.81 | 209.45 (178.36–248.21) | -17.64 (-59.66–20.62) | 0.0958 | 0.3185 |
| Protein (g) | 19 | 82.23 (76.52–111.76) | 97.87 (89.27–110.98) | 10.2 (-9.97–25.58) | 0.1650 | 20 | 98.39 ± 24.91 | 92.36 (79.48–114.19) | -7.41 (-20.79–19.41) | 0.6314 | 0.2112 |
| Fat (g) | 19 | 75.83 ± 20.37 | 78.66 (72.1–91.42) | 5.04 (-8.65–18.06) | 0.3348 | 20 | 86.42 (72.06–91.69) | 78.97 (67.43–92.98) | 3.4 ± 32.64 | 0.8373 | 0.8857 |
| Fiber (g) | 19 | 10.15 (8.75–16) | 13 (9.76–18.3) | 0.79 (-0.39–2.68) | 0.1909 | 20 | 12.64 ± 4.49 | 10.31 (8.66–13.79) | -1.65 ± 5.93 | 0.2276 | 0.1285 |

Values are the mean ± SD (data with normal distribution) or median (interquartile range (IQR): 25% -75%) (data with non-normal distribution). *Number of participants with data available at pre and post- control or iER for each outcome. The outcome variables from pre to post-control or iER are represented by "Δ" in the table. Within-group comparisons for pre-versus post-control or iER were performed by paired t tests (data with normal distribution) or paired Wilcoxon tests (data with non-normal distribution), and between-group comparisons of changes were tested (Δ-p value) by independent sample t tests (data with normal distribution) or Mann-Whitney U tests (data with non-normal distribution).

**Table S6. Glucose-lowering medication changes, related to Figure 1.**

| Glucose-lowering medication use | Control | | | | | iER | | | | | Δ-p value |
|---|---|---|---|---|---|---|---|---|---|---|---|
| | n* | Pre | Post | Δ | p value | n* | Pre | Post | Δ | p value | |
| No glucose-lowering agents | 19 | 3 (15.8) | 3 (15.8) | 0 | 1.0000 | 20 | 6 (30) | 16 (80) | 10 | 0.0015 | 0.0002 |
| 1 glucose-lowering agent | 19 | 6（31.6） | 7 (36.8) | 1 | 0.7324 | 20 | 7 (35) | 3 (15) | -4 | 0.1441 | 0.1339 |
| 2 glucose-lowering agent | 19 | 8（42.1） | 8 (42.1) | 0 | 1.0000 | 20 | 5 (25) | 1 (5) | -4 | 0.0765 | 0.0303 |
| ⩾ 3 glucose-lowering agents | 19 | 2（10.5） | 1 (5.3) | -1 | 0.5475 | 20 | 2 (10) | 0 (0) | -2 | 0.1468 | 0.5174 |
| Insulin | 19 | 2 (10) | 1 (5) | -1 | 0.5475 | 20 | 3 (15) | 1 (5) | -2 | 0.2918 | 0.5174 |
| Metformin | 19 | 11 (57.9) | 11 (57.9) | 0 | 1.0000 | 20 | 9 (45) | 3 (15) | -6 | 0.0384 | 0.0063 |
| Acarbose | 19 | 4 (21.1) | 3 (15.8) | -1 | 0.6756 | 20 | 1 (5) | 0 (0) | -1 | 0.3112 | 0.9703 |
| Sulfonylurea | 19 | 4 (21.1) | 3 (15.8) | -1 | 0.6756 | 20 | 1 (5) | 0 (0) | -1 | 0.3112 | 0.9703 |
| DPP4 Inhibitor | 19 | 0 (0) | 0 (0) | 0 | - | 20 | 2 (10) | 0 (0) | -2 | 0.1468 | 0.1363 |
| SGLT2 Inhibitor | 19 | 8 (42.1) | 7 (36.8) | -1 | 0.7400 | 20 | 4 (20) | 0 (0) | -4 | 0.0350 | 0.1339 |
| Thiazolidinediones | 19 | 0 (0) | 0 (0) | 0 | - | 20 | 3 (15) | 1 (5) | -2 | 0.2918 | 0.1363 |

Values are the frequencies n (%) for categorical variables. *Number of participants with data available at pre and post-control or iER for each outcome. Numbers of participants prescribed no, one, two, or three or more antidiabetic medications in the control group and iER group are presented. The outcome variables from pre to post-control or iER are represented by "Δ" in the table. The p values are for the within-group comparisons and between-group comparisons (Δ-p value) by chi-square tests (data with proportions) for categorical variables.

Abbreviations: DPP4, Dipeptidyl Peptidase-4; SGLT2, Sodium-Glucose Cotransporter-2.

**Table S7. Diet Ingredients and daily intake information of human Chinese medical nutrition therapy (CMNT) Diet, related to Figure 1.**

| Diet Ingredients | Daily intake | Note |
|---|---|---|
| **Breakfast** | | |
| Fruit and vegetable gruel | 50g | Fresh pumpkins, Pumpkin seed kernel oil, Maltodextrins, Isomalto-oligosaccharide, Casein, Resistant dextrin, Sodium ascorbate, Potassium citrate, Mono-and diglycerides of Fatty acids esters, Vitamin E, Tea polyphenols and silicon dioxide. |
| **Lunch** | | |
| Solids beverages | 25g | Pumpkin seed kernel oil, Isomalto-oligosaccharide, Casein, Resistant dextrin, Sodium ascorbate, Potassium citrate, Mono-and diglycerides of fatty acids esters, Vitamin E, Tea polyphenols and silicon dioxide. |
| Composite nutritional rice | 60g | **Homologous medicine and food substance:** *Fructus lycii*, G*anoderma lucidum, Folium Mori*, *Poria cocos, Dioscorea opposita Thunb.*(Chinese yam), *Radix Puerariae*, *Cordyceps militaris*, *Momordica grosvenori.*<br>**Wholegrains and others:** Rice, Millet, Corn, Buckwheat, Quinoa, Oat, Spinach powders, Lily root flour, Cucumber powders, Mushroom powder, Wheat dietary fiber, Bitter melon, Pumpkins, Potato, Purple potato, Sweet potato, Mung bean, Konjac flour, Inulin, and edible refined salt. |
| **Dinner** | | |
| Solids beverages | 25g | Pumpkin seed kernel oil, Isomalto-oligosaccharide, casein, Resistant dextrin, Sodium ascorbate, Potassium citrate, Mono-and diglycerides of fatty acids esters, Vitamin E, Tea polyphenols and silicon dioxide. |
| Meal replacement biscuit | 30g | **Homologous medicine and food substance , wholegrains and others:** *Dioscorea opposita Thunb*.(Chinese yam), Wheat flour, MAIKERENJIA, Mix powder (Quinoa, White kidney, Wheat germ, Azuki bean, Black beans, Yellow beans, Liriopes radix, Glutinous rice, Black rice, Maize, Round bract Plantago ovata husk power, Oat, Buckwheat, Hawthorn, Roselle, millet, Brown rice, Chinese jujube, Chinese wolfberry, Pecan nuts, Chia seed, Black sesame, White sesame, Shiitake mushroom, Laminaria hyperborean, Coffee ), Edible vegetable oils, Potato protein, Wheat dietary fiber powder, Resistant dextrin, Maltodextrin, L-arabinose. |

**Table S8. Calorie information of human Chinese medical nutrition therapy (CMNT) Diet, related to Figure 1.**

| Calorie Information | Solids beverages | Fruit and vegetable gruel | Composite nutritional rice | Meal replacement biscuit |
|---|---|---|---|---|
| Energy density (kcal/100g) | 576.24 | 533.22 | 358.75 | 489.96 |
| Protein (g/100g) | 7.20 | 3.40 | 10.50 | 7.10 |
| Protein % | 5.28 | 2.52 | 11.86 | 5.97 |
| Fat (g/100g) | 50.00 | 30.80 | 1.80 | 18.20 |
| Fat % | 84.09 | 52.39 | 4.66 | 35.13 |
| Carbohydrates (g/100g) | 14.50 | 60.80 | 73.90 | 70.00 |
| Carbohydrates % | 10.63 | 45.09 | 83.48 | 58.90 |
| Total fiber (g/100g) | 23.22 | 11.93 | 9.10 | 7.21 |
| Soluble fiber (g/100g) | 0.81 | 1.92 | 7.14 | 4.02 |
| Insoluble fiber (g/100g) | 22.41 | 11.93 | 1.96 | 3.19 |
| Choline (mg/100g) | 9.39 | 11.6 | 87.8 | 89.7 |
| Sodium (mg/100g) | 63.00 | 95.00 | 41.20 | 264.00 |

**Table S9. Mouse CMNT diet calorie information, related to Figure 3**

| **Mouse CMNT Diet** | |
|---|---|
| **Energy** | |
| Energy density (kcal/100g) | 489.50 |
| Protein (g/100g) | 8.21 |
| Protein % | 9.02 |
| Fat (g/100g) | 18.50 |
| Fat % | 46.63 |
| Carbohydrates (g/100g) | 40.37 |
| Carbohydrates % | 44.36 |
| Fiber (g/100g) | 7.68 |
| Sodium (mg/100g) | 92.65 |

**Table S10. Mouse regular chow diet calorie information, related to Figure 3**

| **Mouse regular chow diet** | |
|---|---|
| **Energy** | |
| Energy density (kcal/100g) | 342 |
| Protein % | 23.07 |
| Fat % | 11.85 |
| Carbohydrates % | 65.08 |

**Table S11. Primer sequences for qRT-PCR or PCR, related to Figure 5-Figure 7.**

| Gene/Primer name | Primer sequence (5’→3’) |
|---|---|
| Human β-actin | Forward: GTGCTATCCCTGTACGCCTC<br>Reverse: GGCCATCTCTTGCTCGAAGT |
| Mouse β-actin | Forward: GGTACCACCATGTACCCAGG<br>Reverse: GGTGTAAAACGCAGCTCAGTAA |
| Human HADHA | Forward: TCAAGCAGGGGAAGGTCA<br>Reverse: CTGGAGGATTCGGATGACTT |
| Mouse HADHA | Forward: TGCATTTGCCGCAGCTTTAC<br>Reverse: GTTGGCCCAGATTTCGATTTC |
| siRNA-NC | Forward: UUCUCCGAACGUGUCACGUTT<br>Reverse: ACGUGACACGUUCGGAGAATT |
| Human si-HADHA-1 | Forward: UGGUGACAAGAUUUGUGAATT<br>Reverse: UUCACAAAUCUUGUCACCATT |
| Human si-HADHA-2 | Forward: CCGCCUGGUGACAAGAUUUTT<br>Reverse: AAAUCUUGUCACCAGGCGGTT |
| HADHA_ K353R<br>(Point A) | Forward：GAATcgaTTTGGAGCTCCACAGAAGGATG<br>Reverse：GAGCTCCAAAtcgATTCTTCTTGCACAGGACCTGAC |
| HADHA-353_8370<br>(Point B) | Forward: CCGACAGGACTATAAAGATACCAGGCGTTTCCCC<br>Reverse: CTTTATAGTCCTGTCGGGTTTCGCC |
| 16S rRNA sequencing | 341F: CCTACGGGNGGCWGCAG<br>805R: GACTACHVGGGTATCTAATCC |
